# Dual Metrics of Obesity: Evaluating BMI and Central Obesity as Indicators of Cardiometabolic Risk in Rural India

**DOI:** 10.1101/2024.08.13.24311925

**Authors:** Raghav Prasad, Manogna Sagiraju, Priya Chatterjee, Pravin Sahadevan, Hitesh Pradhan, Jonas S Sundarakumar, SANSCOG Team

## Abstract

**Purpose:** Indians face a higher risk of cardiometabolic diseases (CMDs) at the same BMI compared to Caucasians. This study explored the use of the Dual Metric Obesity Criteria (DMOC), combining central obesity (CO) indices with BMI, to assess sex-specific associations with CMDs in rural Indians.

**Methods:** Baseline cross-sectional data from 3,397 participants aged ≥45 from the Centre for Brain Research-Srinivaspura Aging, Neuro Senescence and COGnition (CBR-SANSCOG) study were analysed. Five obesity indices were examined: BMI, Waist Circumference, Waist-Hip Ratio (WHR), Waist-Height Ratio (WHtR), and Visceral Fat Percentage. Cohen’s Kappa assessed the agreement between BMI and CO indices. Participants were classified based on BMI and CO status: NWNC (Normal Weight No CO), AWNC (Abnormal Weight No CO), NWCO (Normal Weight CO), and AWCO (Abnormal Weight CO). Multinomial logistic regression models evaluated associations between DMOC groups and CMDs. Interaction analyses explored CO and CMD relationships across sex, BMI categories, and age groups. The mediation effect of CO indices on the relationship between BMI and cardiometabolic risk factors was investigated.

**Results:** BMI showed a slight to fair agreement with WHR and moderate agreement with WHtR. Individuals with AWCO had the highest odds for all CMDs (p<0.001). Males with NWCO had a 3.51 (2.14,6.04) times increased odds times for diabetes and 2.04 (1.22,3.37) times odds for dyslipidemia. Individuals < 58 years and males had stronger associations with CMDs.

**Conclusion:** Combining CO indices with BMI effectively identifies high-risk CMD groups in rural Indians, including those with NWCO a previously overlooked group.

## 1. Introduction

Obesity is fast emerging as a major global public health exigency. Even though the burden of obesity has often been associated with high-income countries, recent studies indicate that low and middle-income countries contribute to 70% of global obesity [1]. Mirroring this trend, the prevalence of obesity in India has also been on the rise with the current prevalence of general obesity at 28.6%, and central obesity (CO) at 39.5% in the adult population [2].

The pathogenesis of obesity is complex and reliant on multiple factors: biological, genetic, socioeconomic, psychosocial and environmental [3]. Similarly, the mechanisms through which obesity-mediated diseases manifest are manifold. Obesity is a common denominator for multiple morbidities: diabetes mellitus (DM), hypertension, dyslipidemia, non-alcoholic fatty liver disease, coronary artery disease, obstructive sleep apnoea, hypoventilation syndrome, dementia, neoplasms and even psychological disorders such as depression and anxiety [4]. These diseases, often interconnected, can appear as simple comorbid (≥2 comorbidities) or complex comorbid (≥4 comorbidities) conditions [5].

For any given Body Mass Index (BMI), south Asians, especially Indians have an excessive tendency to accumulate fat – a phenomenon commonly known as thin-fat [6]. Compared to other ethnic groups, Asian Indians tend to have higher waist circumference (WC) and waist-to-hip ratio (WHR) indicating a greater degree of CO as well as visceral obesity and lesser subcutaneous fat deposition [7]. This phenotype, also known as the Asian-Indian phenotype (AIP), is associated with increased levels of plasma insulin, insulin resistance, a higher prevalence of DM, and lower skeletal muscle mass [8]. AIP comprises a metabolically obese group which consists of people with CO within the normal BMI range. Central or android obesity has been associated with a higher risk of developing cardiometabolic diseases (CMDs) compared to gynoid or peripheral obesity [9].

Obesity in the aging population in India is emerging as a significant public health concern. A recently conducted cross-sectional survey reported that one in every four individuals over the age of 60 years were overweight or obese [10]. Venkatrao et al. reported a higher prevalence of obesity in individuals aged 40 and above compared to those under 40 [11]. Additionally, in individuals over 60, the prevalence of CO exceeds that of the general population with higher rates observed in females compared to males [12]. Aging is associated with a decrease in basal metabolic rate and muscle mass, thus leading to an increased body fat percentage.

Obesity, in turn, accelerates cell-senescence by contributing to cellular, genetic and epigenetic changes creating a vicious feedback loop resulting in a decline in health and quality of life [13], [14]. With India’s rapidly growing aging population set to comprise one-fifth of the global population over 60 by 2050 [15], obesity in this group is a critical health concern that must be addressed immediately. This underscores the importance of tailored strategies for managing obesity that consider age- and sex-related factors to effectively improve health outcomes.

Obesity is on the rise in rural India, with the prevalence of general obesity at 21.3% and CO at 33.5% [2]. Although urban obesity rates remain greater, the steep increase in the prevalence of rural obesity over the last decade is alarming [16]. This is especially concerning given the low health literacy in rural areas and limited access to health care. While the government has made efforts to raise awareness and screen for diabetes and hypertension, similar initiatives for obesity have not been taken. With nearly 70% of India’s population residing in rural areas, even a small rise in rural obesity could lead to a substantial increase in the number of individuals with obesity nationwide [17].

While BMI has been traditionally utilized as the principal indicator for assessing obesity status and the risk associated with CMDs, this index does not consider fat distribution, which often varies across populations, resulting in an underestimation of the obesity burden. To remedy this shortcoming, different measures that consider body fat distribution, such as WC, WHR, waist-height ratio (WHtR), and visceral fat percentage (VFP) are commonly used. Whole-body MRI data has established that WC in combination with BMI, addresses the additional variation in fat distribution and this variation is primarily due to the effectiveness of WC measurements in predicting visceral fat [18]. However, there exists a degree of ambiguity arising from the diverse criteria used to define CO, such as the varied guidelines for WC cutoffs. Thus, there is a growing body of evidence that points to the limitations of BMI but there is still uncertainty about which alternative measure or combination of measures is the most effective.

In this study, we aimed to assess the association between various obesity measures (BMI, WC, WHR, WHtR, VFP) and CMDs, including hypertension, diabetes, and dyslipidemia. We evaluated the effectiveness of a composite index, the Dual Metric Obesity Criteria, which combines BMI with any one central obesity measure, in predicting CMD risk in the aging rural Indian population. Our goal was to identify populations at increased risk and elucidate the mechanisms by which central adiposity contributes to CMD development.

## 2. Methods

### 2.1. Study Design and Participants

The study sample was derived from baseline data from the ongoing Centre for Brain Research - Srinivaspura Aging, Neuro Senescence, and COGnition (CBR-SANSCOG) study. The cohort consists of individuals aged 45 and above from the villages of Srinivaspura in Kolar district, Karnataka, India. The detailed study protocol has been published elsewhere [19]. The baseline data was collected up to April 2023, resulting in a total of 6,274 participants. From this group, we included only those who had complete data for all the obesity-related measurements and cardiometabolic markers (CMMs) included in this study, totalling 3,860 participants. We then excluded those with a BMI less than 18.5, resulting in a final cohort of 3,397 participants, as shown in **Figure 1**.

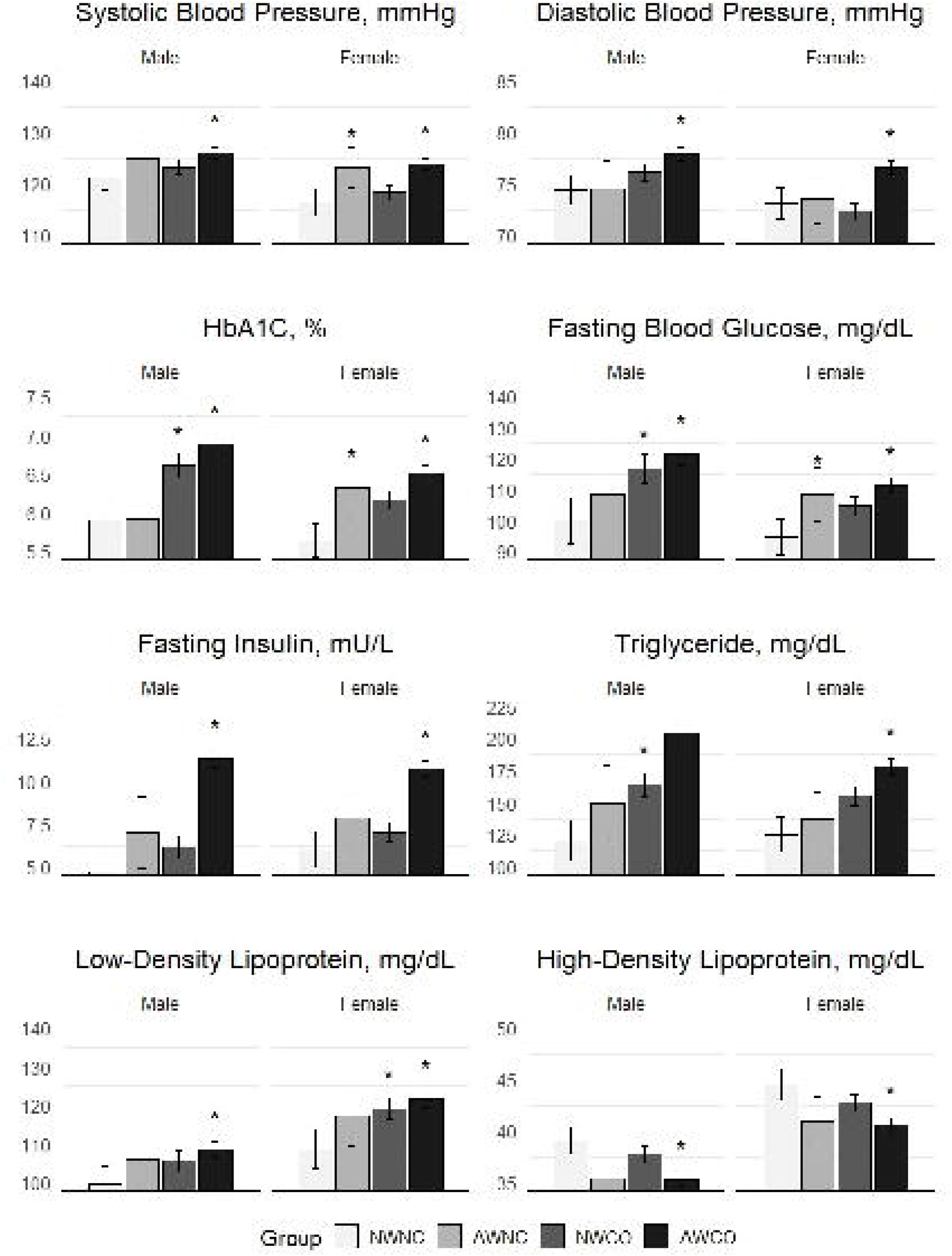

### 2.2. Ethics clearance and informed consent

This cohort study has been approved by the Institutional Ethics Committee of the Centre for Brain Research, Indian Institute of Science (CBR/42/IEC/2022-23). Informed consent was taken from all participants before recruitment into the study.LJLJ

### 2.3. Anthropometric Measures

#### 2.3.1. Body Mass Index (BMI)

Weight in kilograms using a standard electronic scale and height in centimetres using a standard stadiometer was measured in bare feet with light clothing in a standing posture. By dividing the weight by height in metres squared, the BMI was calculated. Utilizing the WHO’s Asia Pacific guidelines for BMI [20] participants with a BMI less than 23 were considered in the normal BMI category and those with a BMI of 23 and above were considered part of the abnormal BMI category (overweight and obese).

#### 2.3.2. Waist Circumference (WC)

Waist circumference was measured using non-stretchable tape at the point above the uppermost lateral border of the right ilium, intersecting with the midaxillary line. Abnormal WC was determined based on three criteria. -

1. International Diabetes Federation (IDF) criteria: Males with a WC ≥ 90 cm and females with a WC ≥ 80 cm [21].
2. World Health Organization 2000 (WHO) criteria: Males with a WC ≥ 94 cm and females with a WC ≥ 80 cm [22].
3. Adult Treatment Panel III (ATP III) criteria: Males with a WC ≥ 102 cm and females with a WC ≥ 88 cm [23].

#### 2.3.3. Waist-Hip Ratio (WHR)

Using a non-stretchable standard measuring tape, the hip circumference was measured horizontally at the largest posterior protuberance of the buttocks while standing. By dividing the waist circumference by the hip circumference, the WHR was calculated. Males with a WHR ≥ 0.9 and females with a WHR ≥ 0.8 met the criteria for CO [24].

#### 2.3.4. Waist-Height Ratio (WHtR)

The WHtR was calculated by dividing the WC by height in centimetres. Males and females with a WHtR ≥ 0.5 met the criteria for CO [25].

#### 2.3.5. Visceral Fat Percentage (VFP)

Visceral obesity was measured using bioelectric impedance (Tanita BC-601, InnerScan V) to get VFP. The cutoff for CO is a VFP ≥ 9% [26].

### 2.4. Dual Metric Obesity Classification (DMOC)

Participants were classified into four groups based on their BMI and CO status:

1. Normal Weight No Central Obesity (NWNC)
2. Abnormal Weight No Central Obesity (AWNC)
3. Normal Weight Central Obesity (NWCO)
4. Abnormal Weight Central Obesity (AWCO)

### 2.5. Cardiometabolic Markers (CMMs)

Systolic blood pressure (SBP) and diastolic blood pressure (DBP) were measured accurately to within 2 mmHg using a Diamond Deluxe mercurial sphygmomanometer in the right arm supine position.

Skilled phlebotomists collected 15 ml of venous blood from fasting participants using vacutainers. Samples were centrifuged onsite at 2000 rpm for 10 minutes immediately after collection. A 4.5 ml serum tube with gel and a 1 ml fluoride tube were sent to a certified partner laboratory, accredited by the National Accreditation Board for Testing and Calibration Laboratories, for biochemical analysis.

Fasting blood glucose (FBG) was measured using the hexokinase method, and Haemoglobin A1c (HbA1c) levels were quantified with a Tosoh Automated Glycohemoglobin Analyzer HLC-723 G8. Fasting insulin levels were determined via electrochemiluminescence immunoassay (ECLIA) on a Cobas e 801 analyser.

The Homeostatic Model Assessment of Insulin Resistance (HOMA-IR) was calculated with the formula:

> HOMA-IR = (fasting glucose × fasting insulin) / 405 [27]

Serum lipid concentrations were measured using various techniques: Total Cholesterol (TC) was assessed through the cholesterol oxidase peroxidase approach, Low-Density Lipoprotein (LDL) was determined using a method involving non-ionic detergent solubilization followed by the action of cholesterol esterase and cholesterol oxidase, Triglycerides (TG) were evaluated by Wahlefeld’s method, and High-Density Lipoprotein (HDL) levels were analyzed using Polyethylene glycol (PEG)-modified cholesteryl esterase and cholesterol oxidase.

The non-HDL cholesterol was calculated using the formula:

> Non-HDL = TC – HDL [28]

The remnant cholesterol (RC) was calculated using the formula:

> RC = TC – (HDL + LDL) [29]

### 2.6 Cardiometabolic Diseases (CMDs)

The following criteria were used to diagnose CMDs:

1. Hypertension: SBP ≥140 mmHg or DBP ≥90 mmHg or self-reported history of hypertension.
2. Diabetes Mellitus: FBG ≥ 126 mg/dL or self-reported history of DM.
3. Dyslipidemia: Presence of self-reported history of dyslipidemia or any one of the following:

a. TC ≥ 200 mg/dL
b. LDL ≥ 160 mg/dL
c. TG ≥ 150 mg/dL
d. HDL < 40 mg/dL (males) and < 50 mg/dL (females)

### 2.7. Covariate assessment

Self-reported questionnaires were used to collect information on age, sex, annual income, years of education, occupation (job skill), tobacco use and alcohol use.

## 3. Statistical analysis

Participants’ demographic characteristics were analysed as follows: categorical variables were described with frequency and percentage, and continuous variables with mean and standard deviation (SD) or median and interquartile range (IQR). Characteristics were compared by sex and BMI status using the Chi-square test for categorical data, and the Mann-Whitney U test or t-test for continuous data. The agreement between CO indices and BMI was evaluated using Cohen’s kappa statistic. Correlations between CO indices and CMMs were calculated using partial correlation, adjusting for age and education, and stratified by sex and BMI status. To analyse the association between DMOC and CMDs, we used multinomial multivariate logistic regression, adjusting for covariates in both sexes. The mean values of CMMs, based on DMOC, were compared using ANCOVA, followed by post hoc Tukey analysis. We analyzed the effects of sex, age, and BMI on the relationship between CO indices and CMMs using multivariable regression with interaction terms, adjusting for confounders. To explore the role of CO in the relationship between BMI and CMMs, we performed an adjusted mediation analysis. This analysis utilized linear regression models for both mediator and outcome variables, conducted using the R ‘mediation’ package. Confidence intervals were estimated through bootstrapping with 1,000 samples to determine the Average Causal Mediation Effects (Indirect) and Average Direct Effects (Direct). Further, the robustness of our findings against potential unmeasured confounding was assessed through sensitivity analyses using the ‘medsens’ function. To assess the diagnostic accuracy of each CO index for CMDs, we calculated the sensitivity and specificity, stratifying the results by BMI status and sex. A p-value of less than 0.05 was considered statistically significant in all analyses. Statistical analyses were performed using Stata version 18.0 and R version 4.3.2.

## 4. Results

### 4.1 Participant Characteristics

The study included 3,397 participants who met the inclusion criteria. Compared to the excluded cohort, these participants were younger (Age: 58.40 vs. 60.20 years, p < 0.001), more educated (Years of education: 4.67 vs. 4.00, p < 0.001), and had a higher percentage of males (49% vs. 47%, p < 0.001).

The baseline characteristics of the study sample are shown in **Table 1**. There was a significant difference in age, education in years, annual income, current tobacco usage, hypertension, and diabetic status between the abnormal and normal BMI groups in both sexes. When including the underweight class, the prevalence of abnormal BMI status was 50.65% for males and 49.37% for females. Within this group, the prevalence of obesity was 33.33% for males and 35.51% for females. The prevalence of CO in the normal BMI group was 33.43% for males and 39.03% for females, while in the abnormal BMI group; it was 65.66% for males and 70.11% for females.

**Table 1:** Baseline Characteristics of participants with respect to BMI status.

| Characteristics | Male n=1,668 |  |  | Female n=1,729 |  |  |
| --- | --- | --- | --- | --- | --- | --- |
|  | Abnormal BMI | Normal BMI | P-value | Abnormal BMI | Normal BMI |  |
|  | n=960 (%) | n=708 (%) |  | n=970 (%) | n=759 (%) |  |
| Age, Mean (SD) | 58.08 (9.31) | 61.57 (9.87) | <0.001 | 56.04 (8.81) | 58.11 (9.59) | <0.001 |
| Education (years), Median (IQR) | 9 (7) | 6 (10) | <0.001 | 1 (7) | 0 (4) | <0.001 |
| Annual Income, less than 50,000 | 607 (63.23) | 507 (71.61) | <0.001 | 767 (79.07) | 636 (83.79) | 0.013 |
| Jobskill level |  |  | 0.1 |  |  | 0.253 |
| Low | 187 (19.48) | 133 (18.79) |  | 351 (36.19) | 265 (34.91) |  |
| Medium | 739 (76.98) | 562 (79.38) |  | 600 (61.86) | 486 (64.03) |  |
| High | 34 (3.54) | 13 (1.84) |  | 19 (1.96) | 8 (1.05) |  |
| Current tobacco user | 227 (23.65) | 263 (37.15) | <0.001 | 258 (26.60) | 311 (40.97) | <0.001 |
| Current alcoholic | 126 (13.13) | 81 (11.44) | 0.302 | 1 (0.10) | 0 (0.00) | 0.376 |
| Hypertension | 412 (42.92) | 225 (31.78) | <0.001 | 357 (36.80) | 179 (23.58) | <0.001 |
| Diabetes | 400 (41.67) | 166 (23.45) | <0.001 | 313 (32.27) | 108 (14.23) | <0.001 |
| Dyslipidemia | 917 (95.52) | 634 (89.55) | <0.001 | 941 (97.01) | 723 (95.26) | 0.057 |
| WC (cm), Mean (SD) | 91.79 (7.16) | 80.27 (5.87) | <0.001 | 86.15 (8.58) | 75.05 (7.60) | <0.001 |
| WHtR, Median (IQR) | 0.55 (0.06) | 0.49 (0.04) | <0.001 | 0.57 (0.08) | 0.49 (0.07) | <0.001 |
| WHR, Median (IQR) | 0.98 (0.07) | 0.94 (0.08) | <0.001 | 0.90 (0.12) | 0.86 (0.14) | <0.001 |
| VFP (%), Median (IQR) | 14 (3) | 10 (4) | <0.001 | 8 (3) | 5 (2) | <0.001 |

*Note. BMI Body Mass Index, WC Waist Circumference, WHtr Waist Height Ratio, WHR Waist Hip Ratio, VFP Visceral Fat Percentage*

### 4.2 Agreement between CO Indices and BMI

Moderate agreement was seen across sexes in WC (IDF, males, K=0.529; females, K=0.497) and WHtR (males, K=0.573; females, K=0.442) and WC in females alone (IDF/WHO, K=0.497). Fair agreement was seen in both sexes in VFP (males, K=0.308; females, K=0.367), WC (WHO, K=0.316) and WHR in males (K=0.243) and WC in females (ATPIII, K=0.331). A slight agreement was seen in WC in males (ATPIII, K=0.072) and WHR in females (K=0.157) as seen in **Table 2**.

**Table 2:** Overlap among participants classified based on BMI with various CO criteria.

| **Central Obesity Criteria** | **Abnormal BMI<sup>a</sup>** | **Normal BMI<sup>a</sup>** | **Cohens Kappa** |
| --- | --- | --- | --- |
| **Male** |  |  |  |
| IDF |  |  | 0.529 |
| Abnormal WC | 589 (61.35) | 37 (5.23) |  |
| Normal WC | 371 (38.65) | 671 (94.77) |  |
| WHO |  |  | 0.316 |
| Abnormal WC | 354 (36.88) | 13 (1.84) |  |
| Normal WC | 606 (63.12) | 695 (98.16) |  |
| ATP III |  |  | 0.072 |
| Abnormal WC | 80 (8.33) | 0 (0.00) |  |
| Normal WC | 880 (91.67) | 708 (100.00) |  |
| WHtR |  |  | 0.573 |
| Abnormal WHtR | 912 (95.00) | 285 (40.25) |  |
| Normal WHtR | 48 (5.00) | 423 (59.75) |  |
| WHR |  |  | 0.243 |
| Abnormal WHR | 911 (94.90) | 515 (72.74) |  |
| Normal WHR | 49 (5.10) | 193 (27.26) |  |
| VO |  |  | 0.308 |
| Abnormal VO | 936 (97.50) | 491 (69.35) |  |
| Normal VO | 24 (2.50) | 217 (30.65) |  |
| **Female** |  |  |  |
| IDF |  |  | 0.497 |
| Abnormal WC | 745 (76.80) | 205 (27.01) |  |
| Normal WC | 225 (23.20) | 554 (72.99) |  |
| WHO |  |  | 0.497 |
| Abnormal WC | 745 (76.80) | 205 (27.01) |  |
| Normal WC | 225 (23.20) | 554 (72.99) |  |
| ATP III |  |  | 0.331 |
| Abnormal WC | 411 (42.37) | 52 (6.85) |  |
| Normal WC | 559 (57.63) | 707 (93.15) |  |
| WHtR |  |  | 0.442 |
| Abnormal WHtR | 869 (89.59) | 357 (47.04) |  |
| Normal WHtR | 101 (10.41) | 402 (52.96) |  |
| WHR |  |  | 0.157 |
| Abnormal WHR | 896 (92.37) | 591 (77.87) |  |
| Normal WHR | 74 (7.63) | 168 (22.13) |  |
| VO |  |  | 0.367 |
| Abnormal VO | 415 (42.78) | 25 (3.29) |  |
| Normal VO | 555 (57.22) | 734 (96.71) |  |

*Note.* WC - Waist Circumference, WHtR - Waist Height Ratio, VO -Visceral Obesity, WHR -Waist Hip Ratio, BMI - Body Mass Index, IDF - International Diabetes Federation, WHO - World Health Organisation, ATP III - Adult Treatment Panel.
*Grades of agreement: 0 indicates no agreement, 0.01–0.20 indicates slight agreement, 0.21–0.40 indicates fair agreement, 0.41–0.60 indicates moderate agreement, 0.61–0.80 indicates substantial agreement, and 0.81–1.00 indicates almost perfect agreement.*
<sup>a</sup> - n (%)

### 4.3 Correlation of CO Indices and CMMs in Normal and Abnormal BMI Groups (Figure 2)

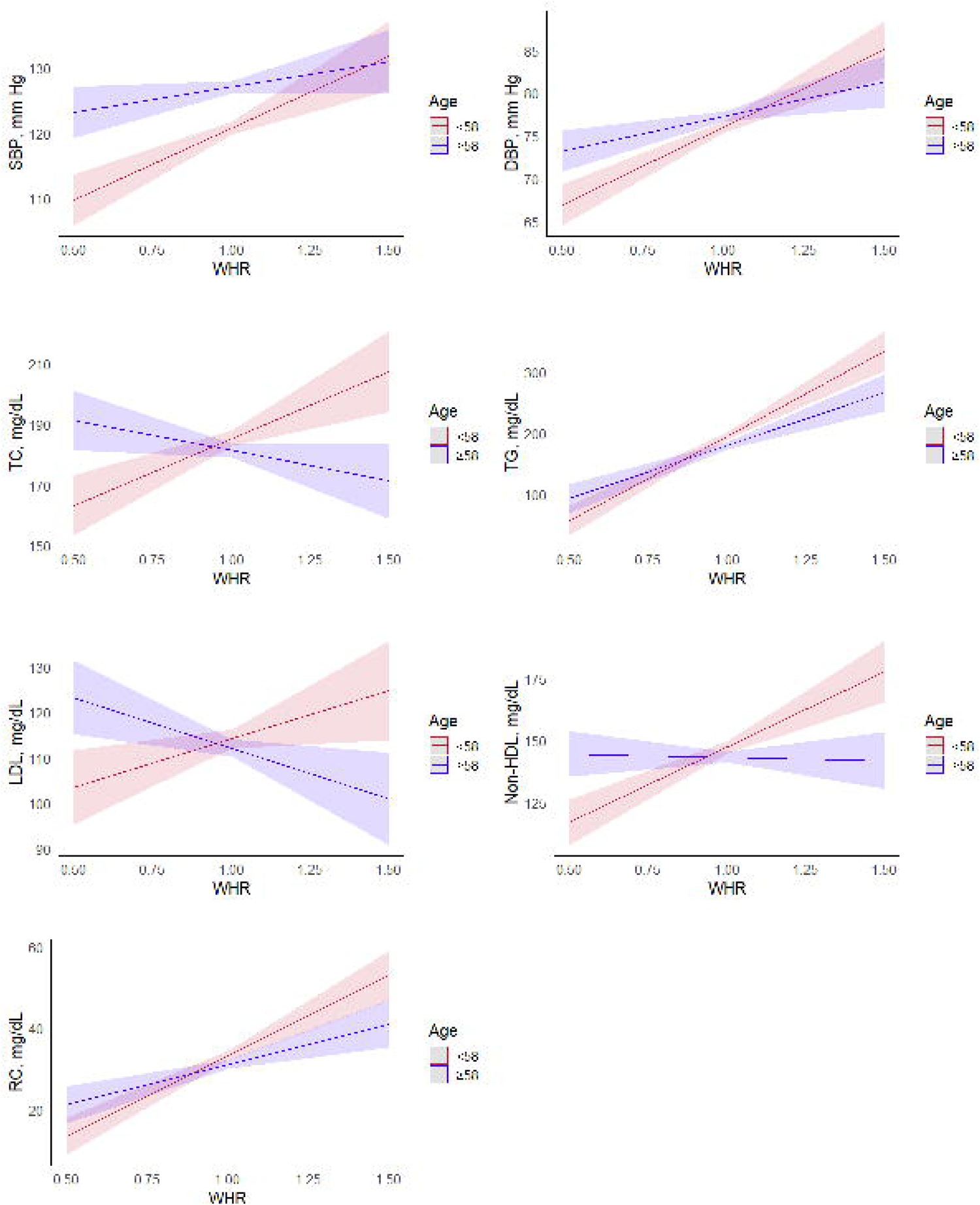

There was an overall positive correlation between various CO indices and CMMs across sexes (**Table S1** and **S2**). The heat map revealed a sex-specific trend: stronger correlations were observed in males with normal BMI and in females with abnormal BMI.

### 4.4 Association of DMOC with CMDs (Figure 3)

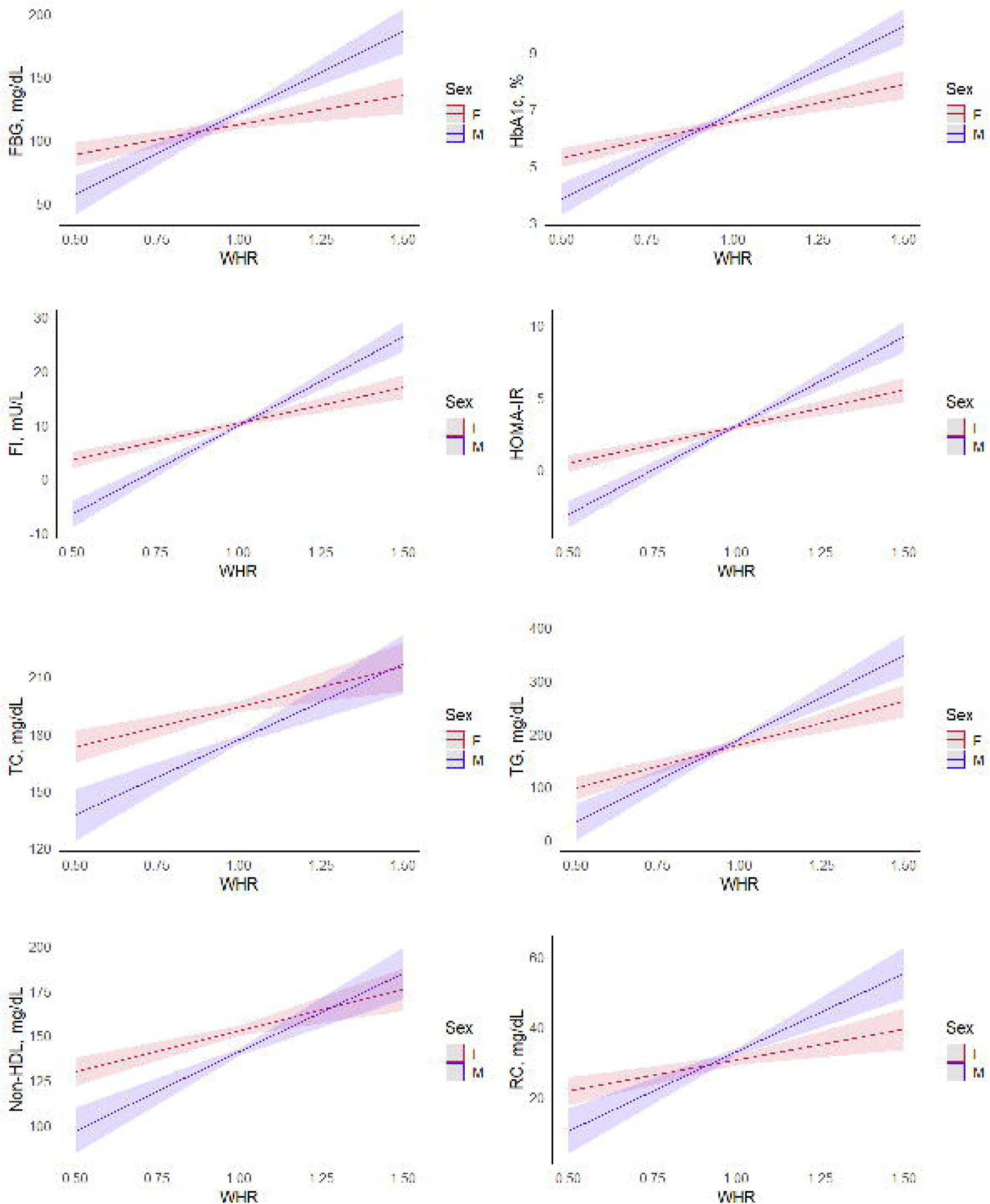

- **AWCO:** This group had increased odds of HTN, DM and Dyslipidemia across both sexes.
- **NWCO:** In this category, males (for all criteria of CO) and females (for all criteria of WC and WHR) had increased odds of DM. Males (for WHtR) had 1.42 times higher odds of HTN. Females (for IDF/WHO criteria of WC) had 1.50 times higher odds of hypertension. Increased odds of dyslipidemia were only seen in males with abnormal WHR and VFP.
- **AWNC:** In this category, males (for all criteria of WC) and females (for all criteria of CO) had increased odds of HTN. Males (for all criteria of WC and VFP) and females (for all criteria of WC and WHR) had increased odds of diabetes. Only males (for all criteria of WC) had increased odds of dyslipidemia.

The individual CO indices are explained in detail in **SS2**. For the lipid parameters, there was a strong association with the AWCO group in both sexes and varying associations for NWCO and AWNC, as seen in **Table S5**.

### 4.5 CMM levels by DMOC groups based on WHR

Individuals with abnormal BMI and WHR consistently had the most adverse risk profile for all CMMs across both sexes. Notably, in the NWCO group, we found significantly higher levels of HbA1c, FBG, and TG in males and LDL in females compared to the NWNC group. Additionally, in the AWNC group, females showed significantly higher levels of SBP, HbA1c and FBG when compared to the NWNC group, whereas no such finding was observed in males (**Figure 4** and **Table S6**). Similar trends were seen for other CO indices with differences seen in the NWCO and AWNC groups when using WC as compared to WHtR or VO as seen in **SS 3**.

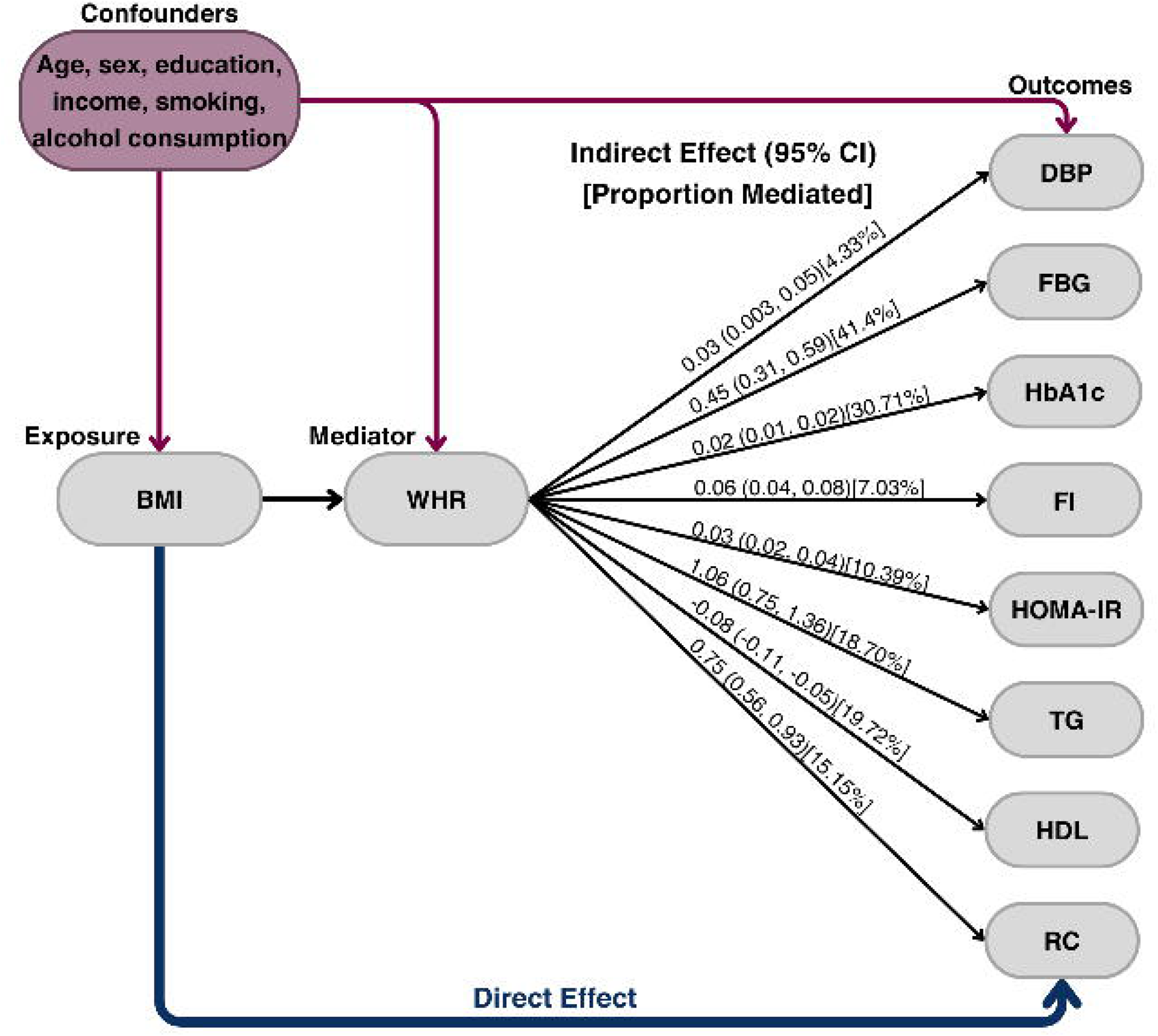

### 4.6 Interaction Effect of Age, Sex and BMI Status

The association between WHR and CMMs demonstrated significant interaction effects with age (**Figure 5** and **Table S12**). Notably, TC, LDL, and Non-HDL cholesterol showed a positive association with WHR in individuals less than 58 years of age, and a negative association in those aged 58 and above. However, this pattern was not observed with RC. A stronger relationship was observed in males compared to females between CMMs and WHR (**Figure 6** and **Table S16**), WHtR, and WC, while the opposite was true for VFP (**SS4.2**). Additionally, a significant interaction effect was found with BMI status, indicating that individuals with abnormal BMI exhibited a stronger relationship between CO indices and CMMs (**SS4.3**).

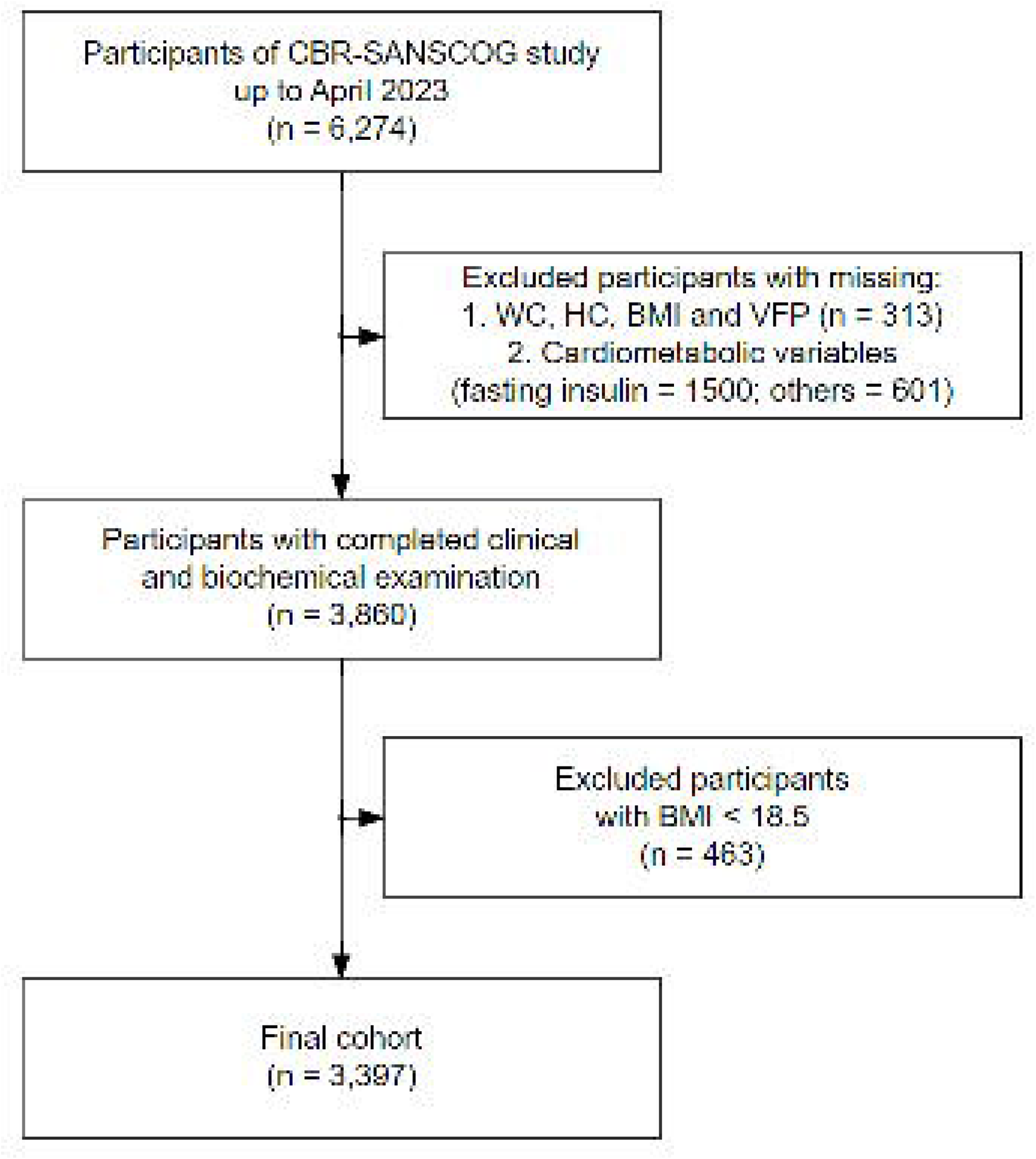

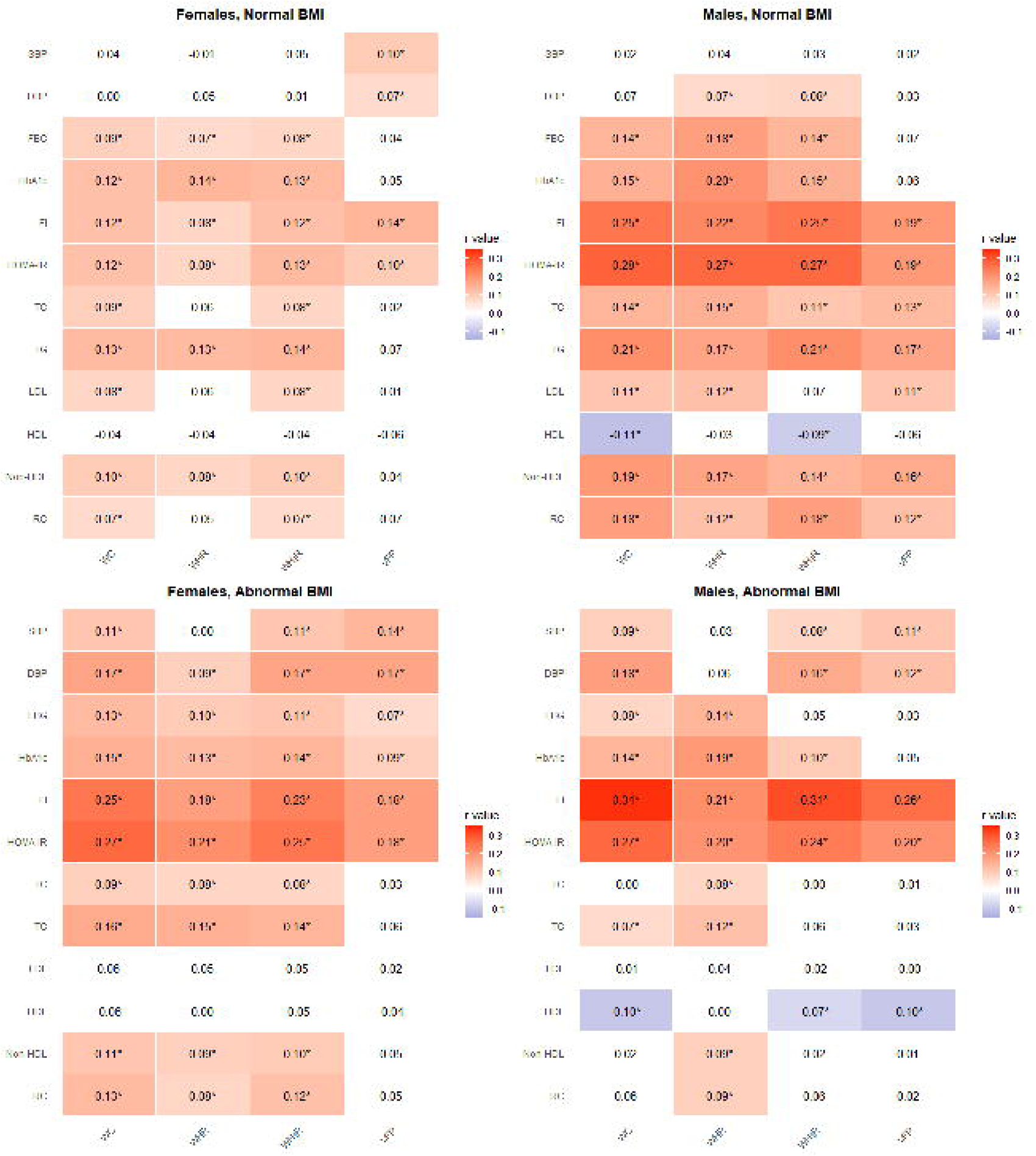

### 4.7 WHR as a Mediator

WHR functioned as a partial mediator in the relationship between BMI and CMMs, particularly those related to diabetes with an average proportional mediation effect of 22% (**Table S24**). It also mediated the associations of DBP (4%), TG (18%), and HDL (20%) as seen in **Figure 7**. **SS5** further illustrates the mediating roles of other CO indices for these associations along with the sensitivity analysis to assess the robustness of the mediation.

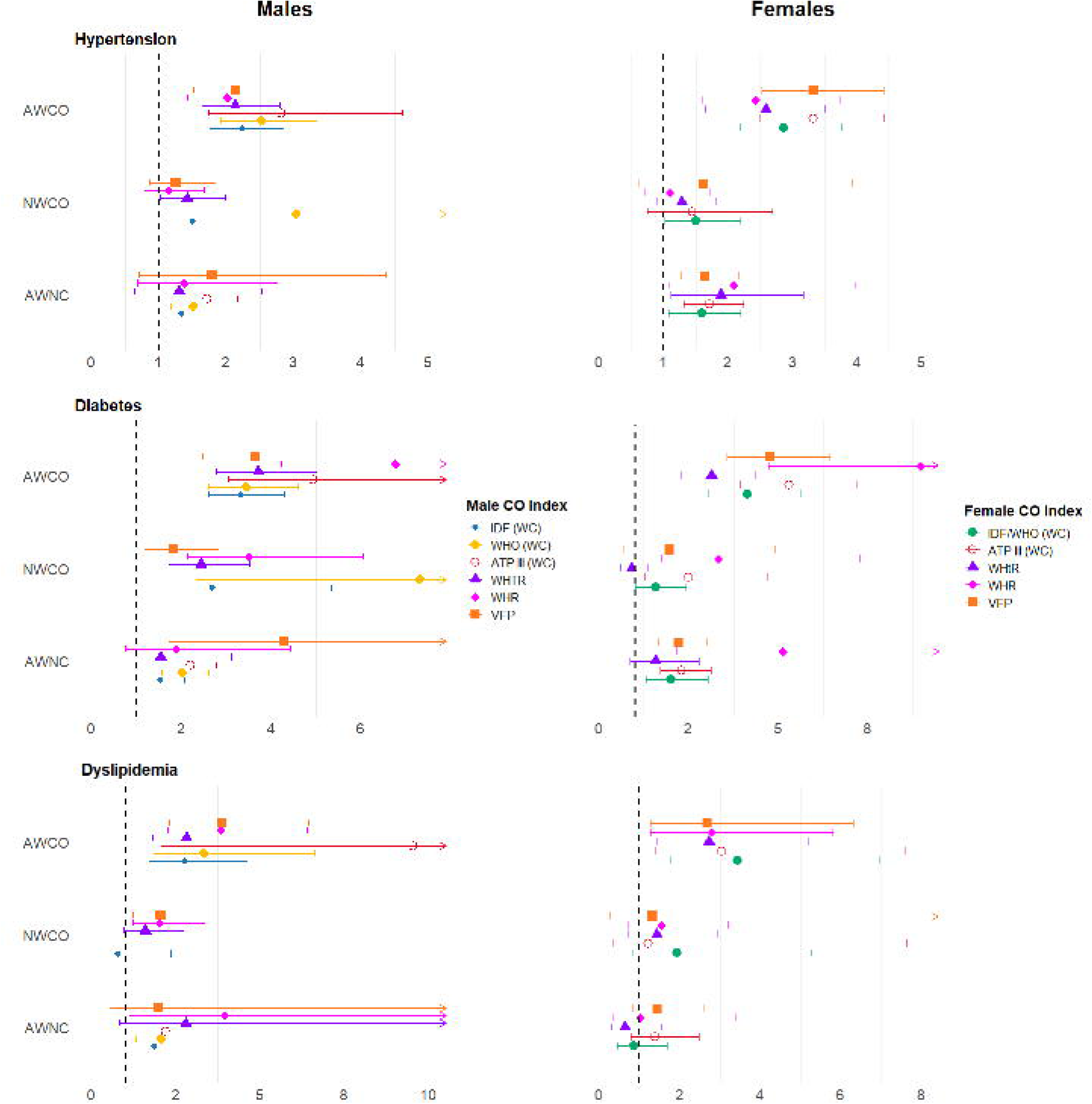

### 4.8 Sensitivity and Specificity of CO Indices based on BMI status

WHR displayed the highest sensitivity across both males and females, regardless of BMI status, as shown in **Table S28**. For females, WHR sensitivity ranged from 78.3% to 94.9% across CMDs, with specificities between 8.2% and 30.6%. For males, sensitivity was 74.1% to 97.8%, with specificities from 4.7% to 39.2%.

## 5. Discussion

While an increased risk of CMDs due to obesity is well-established, our analysis of an aging rural Indian population revealed that the abnormal weight centrally obese (AWCO) group had the highest levels of Cardiometabolic markers (CMMs) and the greatest odds for Cardiometabolic diseases (CMDs). Interestingly, we found that the normal weight centrally obese (NWCO) group showed a higher prevalence of CMDs compared to the normal weight not centrally obese (NWNC) group.

Most CO markers had fair to moderate agreement with BMI across both sexes. Levels of CO indices were predictably higher in the abnormal BMI group for both sexes. There was a 95% overlap in the abnormal BMI group for WHR and WHtR. Interestingly, in the normal BMI group; there were almost 50% of individuals with abnormal CO indices. This group, identified as NWCO, exhibited a significantly higher prevalence of CMDs, with males showing a greater magnitude of association than females. Previous studies have also reported a significant increase in cardiovascular disease risk in NWCO individuals compared to individuals with other types of fat distribution such as the AWNC group [30].

Despite the increasing recognition of NWCO as a significant risk factor for various CMDs, its role in the Indian population remains understudied. Recent years have seen growing interest with a study in the LASI cohort placing its prevalence at 33.9% in females and 17.8% in males [31]. In keeping with this trend, our study showed a higher prevalence of NWCO in females (34.29%) compared to males (29.04%), however, males exhibited a stronger association with CMDs compared to females. This finding may be explained by the protective effect of femoral-gluteal fat distribution in women against CMDs, as suggested by other studies [32].

The AWCO group exhibited the strongest association with CMDs across both sexes, identifying them as a critically high-risk group. Among participants with abnormal BMI, increased WHR seemed to have a stronger association with CMDs compared to the other CO indices. Within the abnormal BMI group, females demonstrated a stronger correlation with CMMs, particularly triglycerides, total cholesterol, Non-HDL, and LDL, possibly indicating different lipid metabolism dynamics between genders. While all groups with either abnormal BMI or CO indices necessitate caution, AWCO comprises a high-risk cohort that warrants intense scrutiny due to its significantly increased association with CMDs compared to groups with abnormal BMI or CO alone. Thus, in resource-poor settings, interventions must be focused on this group.

Across sexes, the magnitude of association with abnormal WHR was especially high for Diabetes. This could be due to a strong link between Diabetes and Obesity. Obesity along with other factors like shared genetic vulnerability, microenvironmental changes impairing insulin signalling and deteriorated β-cell function play an important role in the development and progression of T2DM. However, T2DM can also occur inversely before obesity in some individuals with inherent insulin resistance resulting in increased hepatic glucose production and elevated insulin levels, which are the primary causes of obesity [33]. An important link between obesity, metabolic syndrome and dyslipidemia, seems to be the development of insulin resistance in peripheral tissues leading to the antilipolytic effects of insulin [34].

Among all the CO indices examined in this study, WHR was identified as the most sensitive and consistent indicator for various cardiometabolic parameters, followed by WHtR. Previous findings have been inconsistent: some studies in Indian and other diverse ethnic groups have shown WHR to be superior to BMI alone, [35], [36], [37] while others suggest that WHtR is a more accurate index [38], [39], [40]. While all three WC criteria identified risk for CMDs, the ATPIII criteria which has the highest cutoff for WC failed to capture individuals with CO within the normal BMI group in males.

Different measures of obesity showed varied sensitivity and specificity for different disease conditions. In the abnormal BMI group in males and females, the same measures showed high sensitivity and specificity across all three diseases (DM, HTN, dyslipidaemia)-WHR, WHtR, and VO in males and WHR, WHtR, and WC (IDF and WHO criteria) in females. Similarly, in the normal BMI group, in males, WC (IDF and WHO) consistently had a high sensitivity and specificity. However, in females with normal BMI, the findings were more random. While WC (ATP) and VO had high specificity for each disease, WHR had the highest sensitivity.

In our study, we measured VFP using bio-electric impedance (BIA), which is a well-regarded method for its utility in epidemiological body composition studies [41]. It demonstrated high sensitivity in males and high specificity in females as a predictor of cardiometabolic risk. Visceral adipose tissue (VAT), which envelops internal organs, is anatomically different from subcutaneous adipose tissue (SCAT), located under the skin. VAT is associated with a higher risk of insulin resistance, hypertension, metabolic syndrome, and hepatic steatosis. In contrast, SCAT can potentially enhance insulin sensitivity and lower the risk of T2DM. Although techniques like BIA and abdominal MRI give a more accurate estimation of VFP, their large-scale implementation in a rural setting is impractical due to resource constraints. Therefore, simpler anthropometric measures could be used as a scalable community-level screening tool to estimate the CMD risk.

Although both WHR and WHtR performed well, WHtR would be the more practical choice to employ in the Indian setting as it is less laborious (as it only requires measuring WC along with BMI) and more culturally appropriate. The relevance of CO indices may vary depending on the specific cardiometabolic disease (CMD) being assessed; for instance, VFP showed very high odds for diabetes. Thus, a combination of these measures could be more pertinent for the comprehensive evaluation of CMD risk.

The large sample size of the study allows us to capture the best predictors of cardiometabolic risk in this rural Indian population, particularly for different disease conditions. Further, objective diagnosis, instead of only self-reporting the CMDs allowed us to assess the actual prevalence of these diseases in the study population, especially in a rural setting, where people are not aware of their existing health conditions. The cross-sectional cohort design limits the establishment of causality. Also, the study population being restricted to one region prevents generalizability in a pan-Indian rural context. The study could be enhanced by implementing a longitudinal design to follow patients with obesity over time and observe the development of CMDs.

## 6. Conclusion

In conclusion, we found central obesity to be an important risk factor for cardiometabolic diseases independently and in combination with abnormal BMI, often acting synergistically. This study employed a novel classification system that combined both central and general obesity indices to indicate high-risk groups that were previously overlooked. We argue the merits of incorporating measures such as waist-to-hip ratio and waist-to-height ratio along with BMI for the assessment of high-risk individuals. Moreover, our findings highlight the use of combinations of anthropometric measures depending on their sensitivity and specificity for specific disease conditions. Finally, our study highlights the need for a composite index -instead of BMI alone-taking both BMI and WHR into account to assess the cardiometabolic risk profile. This will reduce the underdiagnosis of obesity, thereby providing a more accurate assessment of its prevalence in India and facilitate the implementation of effective measures to address the obesity epidemic in the country.

## Supporting information

Graphical Abstract

## Data Availability

The data supporting the findings of this study can be made available upon reasonable request to the corresponding author, in accordance with the Centre for Brain Research's data sharing policy and the statutory requirements of the Government of India.

## ABBREVIATION LIST

AIP: Asian-Indian Phenotype, AWCO Abnormal Weight Central Obesity
AWNC: Abnormal Weight No Central Obesity
BMI: Body Mass Index
CMDs: Cardiometabolic Diseases
CMMs: Cardiometabolic Markers
CO: Central Obesity
DBP: Diastolic Blood Pressure
DM: Diabetes Mellitus
DMOC: Dual Metric Obesity Criteria
FBG: Fasting Blood Glucose
HbA1c: Hemoglobin A1c
HDL: High-Density Lipoprotein
HOMA-IR: Homeostatic Model Assessment of Insulin Resistance
HTN: Hypertension
IDF: International Diabetes Federation
LDL: Low-Density Lipoprotein
Non-HDL: Non-High-Density Lipoprotein
NWCO: Normal Weight Central Obesity
NWNC: Normal Weight No Central Obesity
RC: Remnant Cholesterol
SBP: Systolic Blood Pressure
TG: Triglycerides
VFP: Visceral Fat Percentage
WC: Waist Circumference
WHR: Waist-Hip Ratio
WHtR: Waist-Height Ratio

## Acknowledgements

We are grateful to the volunteers who participated in the SANSCOG study. We acknowledge all members of the SANSCOG study team for their valuable contributions in various aspects of the study. We would like to extend our special thanks to Dr. Palash Kumar Malo for his invaluable expertise in statistics.

## Author contributions

**Raghav Prasad**: Conceptualization, Methodology, Formal analysis, Writing – Original Draft, Writing – Review & Editing, Visualization; **Manogna Sagiraju**: Writing – Review & Editing; **Priya Chatterjee**: Writing – Review & Editing; **Pravin Sahadevan**: Conceptualization, Formal analysis; **Hitesh Pradhan**: Data Curation; **Jonas S Sundarakumar**: Supervision.

