## Supplementary Material for "Dual Metrics of Obesity: Evaluating BMI and Central Obesity as Indicators of Cardiometabolic Risk in Rural India"

**Appendix**

**Supplementary Section 1**

### *SS1. Correlation of Central Obesity Indices with Cardiometabolic Markers Stratified by Sex and BMI Status*

There were significant correlations between cardiometabolic markers (CMMs) and central obesity (CO) indices in both sexes as seen in **Figure 2 and** in **Table S1** and **S2**, except for the following:

1. **Normal BMI**
   - **Males**: No correlation between:
     - WC and SBP
     - WHtR and LDL
     - WHR and HDL
   - **Females**: No correlation between:
     - WC and SBP, DBP, or HDL
     - WHtR and DBP or HDL
     - WHR and SBP, DBP, LDL, HDL, and remnant cholesterol
2. **Abnormal BMI**
   - **Males**: No correlation between:
     - WC or WHtR and TC, LDL, TG, Non-HDL cholesterol, and remnant cholesterol
     - WHR and SBP, LDL, and HDL
   - **Females**: No correlation between:
     - WC or WHtR and HDL
     - WHR and SBP, LDL, or HDL

| **Table S1** |  |  |  |  |  |  |  |  |
| --- | --- | --- | --- | --- | --- | --- | --- | --- |
| *TS1. Partial Correlation of central obesity indices and cardiometabolic markers in females and males with normal body mass index.* | | | | | | | | |
| **Cardiometabolic markers** | **Female** | | | | **Male** | | | |
|  | **WC** | **WHtR** | **WHR** | **VFP** | **WC** | **WHtR** | **WHR** | **VFP** |
| SBP | 0.06 | 0.12* | 0.02 | 0.21* | 0.06 | 0.10* | 0.09* | 0.12* |
| DBP | 0.02 | 0.05 | -0.03 | 0.13* | 0.08* | 0.10* | 0.09* | 0.07 |
| FBG | 0.09* | 0.09* | 0.08* | 0.06 | 0.13* | 0.12* | 0.16* | 0.05 |
| HbA1C | 0.13* | 0.14* | 0.14* | 0.08* | 0.16* | 0.15* | 0.20* | 0.08* |
| Fasting Insulin | 0.12* | 0.13* | 0.08* | 0.15* | 0.24* | 0.22* | 0.20* | 0.15* |
| HOMA-IR | 0.12* | 0.14* | 0.09* | 0.12* | 0.27* | 0.24* | 0.25* | 0.15* |
| TC | 0.09* | 0.11* | 0.07* | 0.06 | 0.13* | 0.09* | 0.13* | 0.10* |
| LDL | 0.09* | 0.10* | 0.07 | 0.04 | 0.10* | 0.06 | 0.11* | 0.09* |
| TG | 0.14* | 0.15* | 0.13* | 0.09* | 0.20* | 0.18* | 0.16* | 0.14* |
| HDL | -0.04 | -0.05 | -0.04 | -0.06 | -0.11* | -0.09* | -0.03 | -0.06 |
| Non-HDL | 0.11* | 0.13* | 0.09* | 0.08* | 0.18* | 0.13* | 0.15* | 0.13* |
| Remnant Cholesterol | 0.08* | 0.09* | 0.06 | 0.09* | 0.18* | 0.17* | 0.11* | 0.11* |
| *VFP Visceral Fat Percentage, WHR Waist-Hip Ratio, WHtR Waist-Height Ratio, WC Waist Circumference, HDL High Density Lipoprotein, HOMA-IR Homeostatic Model Assessment for Insulin Resistance, TC Total Cholesterol, LDL Low Density Lipoprotein, FBG Fasting Blood Glucose, SBP Systolic Blood Pressure, DBP Diastolic Blood Pressure, TG Triglyceride.* | | | | | | | | |
| **P-value<0.05* |  |  |  |  |  |  |  |  |

*Adjusted for age and education.*

| **Table S2** |  |  |  |  |  |  |  |  |
| --- | --- | --- | --- | --- | --- | --- | --- | --- |
| *TS2. Partial Correlation of central obesity indices and cardiometabolic markers in females and males with abnormal body mass index.* | | | | | | | | |
| **Cardiometabolic markers** | **Female** | | | | **Male** | | | |
|  | **WC** | **WHtR** | **WHR** | **VFP** | **WC** | **WHtR** | **WHR** | **VFP** |
| SBP | 0.13* | 0.16* | 0.03 | 0.17* | 0.10* | 0.11* | 0.04 | 0.17* |
| DBP | 0.18* | 0.20* | 0.10* | 0.19* | 0.19* | 0.17* | 0.06* | 0.15* |
| FBG | 0.13* | 0.12* | 0.11* | 0.08* | 0.08* | 0.03 | 0.12* | 0.008 |
| HbA1C | 0.16* | 0.15* | 0.14* | 0.10* | 0.14* | 0.10* | 0.18* | 0.06 |
| Fasting Insulin | 0.25* | 0.23* | 0.18* | 0.18* | 0.33* | 0.30* | 0.19* | 0.24* |
| HOMA-IR | 0.27* | 0.25* | 0.21* | 0.18* | 0.27* | 0.22* | 0.18* | 0.17* |
| TC | 0.10* | 0.11* | 0.09* | 0.05 | -0.003 | -0.005 | 0.07* | -0.02 |
| LDL | 0.07* | 0.07* | 0.06 | 0.04 | -0.01 | -0.02 | 0.04 | -0.002 |
| TG | 0.16* | 0.15* | 0.15* | 0.06 | 0.06 | 0.04 | 0.10* | 0.003 |
| HDL | -0.05 | -0.04 | 0.006 | -0.03 | -0.10* | -0.07* | 0.007 | -0.09* |
| Non-HDL | 0.13* | 0.13* | 0.10* | 0.07* | 0.02 | 0.01 | 0.08* | -0.006 |
| Remnant Cholesterol | 0.13* | 0.1* | 0.09* | 0.06 | 0.05 | 0.04 | 0.07* | -0.01 |
| *VFP Visceral Fat Percentage, WHR Waist-Hip Ratio, WHtR Waist-Height Ratio, WC Waist Circumference, HDL High Density Lipoprotein, HOMA-IR Homeostatic Model Assessment for Insulin Resistance, TC Total Cholesterol, LDL Low Density Lipoprotein, FBG Fasting Blood Glucose, SBP Systolic Blood Pressure, DBP Diastolic Blood Pressure, TG Triglyceride.* | | | | | | | | |
| **P-value<0.05* |  |  |  |  |  |  |  |  |

*Adjusted for age and education.*

**Supplementary Section 2**

### *SS2. Association of Dual Metric Obesity Criteria Categories based on various Central Obesity indices with Cardiometabolic Diseases*

The association of hypertension, diabetes and dyslipidemia with DMOC based on WHR, WHtR and VO are the following (**Fig3, Table S3, S4**):

**SS2.1 WHTR**

**AWCO**: Both sexes with abnormal BMI and WHTR had significantly (p<0.001) higher odds of hypertension [Males (OR=2.13, CI 1.64-2.79); Females (OR=2.59, CI 1.64-3.50)], diabetes [Males (OR=3.71, CI 2.77-5.02); Females (OR=3.14, CI 2.29-4.37)] and dyslipidemia [Males (OR=2.82, CI 1.81-4.40); Females (OR=2.73, CI 1.45-5.21)].

**NWCO**: Males with normal weight but abnormal WHTR had higher odds of hypertension (OR=1.42, CI 1.02-1.98, p<0.05) and diabetes (OR=2.44, CI 1.71-3.52, p<0.001). No significant associations were observed in females.

**AWNC**: Females with abnormal weight but normal WHTR had higher odds of hypertension (OR=1.89, CI 1.11-3.17, p<0.05) whereas no associations were seen in males.

**SS2.2 WHR**

**AWCO:** Both sexes with abnormal BMI and WHR had significantly (p<0.001) higher odds of hypertension [Males (OR=2.02, CI 1.42-2.90); Females (OR=2.43, CI 1.61-3.74)], diabetes [Males (OR=6.77, CI 4.22-11.47); Females (OR=8.98, CI 4.74-19.30)] and dyslipidemia [Males (OR=3.83, CI 2.27-6.39); Females (OR=2.79, CI 1.27-5.81)].

**NWCO**: Both sexes with normal BMI and abnormal WHR had higher odds of diabetes [Males (OR=3.51, CI 2.14-6.04); Females (OR=3.33, CI 1.72-7.27)] with p<0.001. Only males were at higher odds of 2.04 for dyslipidemia (CI 1.22-3.37; p<0.01). No significant associations were observed with hypertension in both sexes and with dyslipidemia in females.

**AWNC:** Females with abnormal BMI and normal WHR had higher odds of 5.13 for diabetes (CI 2.16-12.85; p<0.001) and 2.09 for hypertension (CI 1.09-3.98; p<0.05). No significant associations were observed in males for hypertension, diabetes, or dyslipidemia.

**SS2.3 VO**

**AWCO:** Both sexes with abnormal BMI and VO had significantly (p<0.05) higher odds of hypertension [Males (OR=2.13, CI: 1.51,3.05; p<0.001); Females (OR=3.33, CI: 2.52,4.41; p<0.001)]; diabetes [Males (OR=3.65 CI: 2.47,5.55; p<0.001); Females (OR=4.78 CI: 3.56,6.45; p<0.001)] and dyslipidemia [Males (OR=3.88 CI:2.31,6.45; p<0.001); Females (OR=2.68 CI: 1.28,6.32; p<0.05)].

**NWCO:** Males with normal BMI and visceral obesity were at 1.81 times increased odds (CI: 1.18,2.42; p<0.01 for diabetes and 2.05 times increased odds (CI: 1.22,3.42; p<0.01) for dyslipidemia.

**AWNC:** Both sexes with abnormal BI and no visceral obesity were at increased odds for diabetes [Males (OR=4.28 CI: 1.72, 10.53; p<0.01); Females (OR=2.21 CI: 1.65, 2.99; p<0.001)]. Females with AWNC were at 1.64 times increased odds (CI:1.26,2.16; p<0.001) for hypertension.

**SS2.4 WC (IDF criteria)**

**AWCO:** Those with abnormal BMI and waist circumference had higher odds of having Hypertension [Males ( OR=2.23, CI 1.75-2.85); females (OR=2.86, CI=2.18-3.76)]; Diabetes [Males (OR=3.33 CI: 2.60,4.30); Females (OR=4.14 CI:3.07,5.65)] and Dyslipidemia [Males (OR=2.76 CI:1.70,4.63); Females (OR=3.44 CI:1.79,6.97)] in both sexes compared to the NWNC group (p<0.001).

**NWCO:** The group with normal weight but abnormal waist circumference showed increased odds of having Diabetes [Males (OR= 2.70 CI: 1.35, 5.34, p< 0.01); Females (OR=1.57 CI: 1.01, 2.42; p<0.05)] across both sexes. Females with NWCO had 1.50 times increased odds (CI: 1.02.2.19; p<0.05) for hypertension.

**AWNC:** There was a significant association between Hypertension [Males (OR=1.33 CI: 1.01, 1.77); Females (OR=1.59 CI: 1.08, 2.19; p<0.05)] and Diabetes [Males (OR=1.53 CI: 1.14, 2.07); Females (OR=2.01, CI: 1.31, 3.05, p<0.01)] in both sexes. Males with AWNC had 1.86 times increased odds (CI: 1.12, 3.22; p<0.05) for dyslipidemia.

**SS2.5 WC (WHO criteria)**

**AWCO:** Those with abnormal BMI and waist circumference had higher odds of hypertension [Males (OR= 2.53 CI: 1.92,3.35, p< 0.001); Females (OR=2.86 CI:2.18,3.76; p<0.001)], diabetes [Males (OR= 3.45 CI: 2.60,4.59, p< 0.001); Females (OR=4.14 CI:3.07,5.65; p<0.001)] and dyslipidemia [Males (OR= 3.35 CI: 1.84,6.62, p< 0.001); Females (OR=3.44 CI:1.79,6.97; p<0.001)] in both sexes.

**NWCO:** The group with normal BMI and abnormal waist circumference had a higher odds for diabetes [Males (OR= 7.31 CI: 2.32, 27.45, p< 0.01); Females (OR=1.57 CI: 1.01, 2.42; p<0.05)] in both sexes. Females with NWCO had 1.50 times increased odds for hypertension (CI: 1.02, 2.19,	p<0.05).

**AWNC:** The group with abnormal BMI and normal waist circumference had higher odds for hypertension [Males (OR= 1.51 CI: 1.18,1.93, p< 0.001); Females (OR=1.59 CI: 1.08,2.19; p<0.05)] and diabetes [Males (OR= 2.01 CI: 1.56,2.50, p< 0.001); Females (OR=2.01 CI:1.31,3.05; p<0.01)] across sexes and dyslipidemia in males alone (OR=2.07 CI: 1.33,3.28; p<0.01).

**SS2.6 WC (ATPIII)**

**AWCO:** Those with abnormal BMI and waist circumference had higher odds of hypertension [Males (OR= 2.82 CI: 1.74,4.62, p< 0.001); Females (OR=3.32 CI:2.50,4.41; p<0.001)], diabetes [Males (OR= 4.94 CI: 3.04,8.13, p< 0.001); Females (OR=5.30 CI:3.92,7.21; p<0.001)] and dyslipidemia [Males (OR= 9.54 CI: 2.06,16.98, p< 0.05); Females (OR=3.05 CI:1.40,7.61; p<0.01)] in both sexes.

**NWCO:** There were no males in the NWCO group as the large metric of the waist circumference in the ATP III criteria removes the possibility of normal BMI. Females with NWCO had 1.50 times increased odds for diabetes (OR=2.50 CI: 1.27, 4.71, p<0.01).

**AWNC:** The group with abnormal BMI and normal waist circumference had higher odds for hypertension [Males (OR= 1.72 CI: 1.37,2.16, p< 0.001); Females (OR=1.71 CI: 1.31,2.24; p<0.001)] and diabetes [Males (OR= 2.20 CI: 1.75,2.77, p< 0.001); Females (OR=2.30 CI:1.71,3.12; p<0.001)] across sexes and dyslipidemia in males alone (OR=2.20 CI: 1.47,3.33; p<0.001) .

| **Table S3** |  |  |  |  |  |  |
| --- | --- | --- | --- | --- | --- | --- |
| *TS3. Association between Types of Dual Metric Obesity Classification and Cardiometabolic Diseases in males* | | | | | | |
| **Variable** | **Odds Ratio (95% Confidence Interval)** | | | | | |
|  | **WC** | | | **WHTR** | **WHR** | **VFP** |
|  | **IDF** | **WHO** | **ATP III** |  |  |  |
| **Hypertension** |  |  |  |  |  |  |
| **NWNC** | Ref | Ref | Ref | Ref | Ref | Ref |
| **AWNC** | 1.33 (1.01,1.77)* | 1.51 (1.18,1.93)*** | 1.72 (1.37,2.16)*** | 1.30 (0.64,2.52) | 1.38 (0.68,2.76) | 1.79 (0.71,4.37) |
| **NWCO** | 1.50 (0.74,2.98) | 3.04 (0.99,10.33) | a | 1.42 (1.02,1.98)* | 1.14 (0.78,1.67) | 1.25 (0.86,1.84) |
| **AWCO** | 2.23 (1.75,2.85)*** | 2.53 (1.92,3.35)*** | 2.82 (1.74,4.62)*** | 2.13 (1.64,2.79)*** | 2.02 (1.42,2.90)*** | 2.13 (1.51,3.05)*** |
| **Diabetes** |  |  |  |  |  |  |
| **NWNC** | Ref | Ref | Ref | Ref | Ref | Ref |
| **AWNC** | 1.53 (1.14,2.07)** | 2.01 (1.56,2.60)*** | 2.20 (1.75,2.77)*** | 1.54 (0.71,3.11) | 1.88 (0.75,4.42) | 4.28 (1.72,10.53)** |
| **NWCO** | 2.70 (1.35,5.34)** | 7.31 (2.32,27.45)** | a | 2.44 (1.71,3.52)*** | 3.51 (2.14,6.04)*** | 1.81 (1.18,2.82)** |
| **AWCO** | 3.33 (2.60,4.30)*** | 3.45 (2.60,4.59)*** | 4.94 (3.04,8.13)*** | 3.71 (2.77,5.02)*** | 6.77 (4.22,11.47)*** | 3.65 (2.47,5.55)*** |
| **Dyslipidaemia** |  |  |  |  |  |  |
| **NWNC** | Ref | Ref | Ref | Ref | Ref | Ref |
| **AWNC** | 1.86 (1.12,3.22)* | 2.07 (1.33,3.28)** | 2.20 (1.47,3.33)*** | 2.80 (0.82,17.59) | 3.95 (1.12,25.15) | 1.97 (0.53,12.85) |
| **NWCO** | 0.78 (0.31,2.36) | b | a | 1.60 (0.96,2.74) | 2.04 (1.22,3.37)** | 2.05 (1.22,3.42)** |
| **AWCO** | 2.76 (1.70,4.63)*** | 3.35 (1.84,6.62)*** | 9.54 (2.06,16.98)* | 2.82 (1.81,4.40)*** | 3.83 (2.27,6.39)*** | 3.88 (2.31,6.45)*** |
| *Note. IDF International Diabetes Federation, WHO World Health Organisation, NCEP ATP III National Cholesterol Education Program Adult Treatment Panel III, WC Waist Circumference, WHtR Waist-Height Ratio, WHR Waist-Hip Ratio, VO Visceral Obesity, NWNC Normal Weight No Central Obesity, AWNC Abnormal Weight No Central Obesity, NWCO Normal Weight Central Obesity, AWCO Abnormal Weight Central Obesity.* | | | | | | |
| *Adjusted for age, education, income, job skill level, tobacco use, and alcohol consumption.* | | | | | | |
| * p < 0.05, ** p < 0.01, *** p<0.001 | | |  |  |  |  |

| **Table S4** |  |  |  |  |  |
| --- | --- | --- | --- | --- | --- |
| *TS4. Association between Types of Dual Metric Obesity Classification and Cardiometabolic Diseases in females* | | | |  |  |
| **Variable** | **Odds Ratio (95% Confidence Interval)** | | | | |
|  | **WC** | | **WHtR** | **WHR** | **VFP** |
|  | **IDF/WHO** | **ATP III** |  |  |  |
| **Hypertension** |  |  |  |  |  |
| **NWNC** | Ref | Ref | Ref | Ref | Ref |
| **AWNC** | 1.59 (1.08,2.19)* | 1.71 (1.31,2.24)*** | 1.89 (1.11,3.17)* | 2.09 (1.09,3.98)* | 1.64 (1.26,2.16)*** |
| **NWCO** | 1.50 (1.02,2.19)* | 1.44 (0.75,2.69) | 1.28 (0.90,1.82) | 1.10 (0.71,1.72) | 1.62 (0.62,3.92) |
| **AWCO** | 2.86 (2.18,3.76)*** | 3.32 (2.50,4.41)*** | 2.59 (1.64,3.50)*** | 2.43 (1.61,3.74)*** | 3.33 (2.52,4.41)*** |
| **Diabetes** |  |  |  |  |  |
| **NWNC** | Ref | Ref | Ref | Ref | Ref |
| **AWNC** | 2.01 (1.31,3.05)** | 2.30 (1.71,3.12)*** | 1.57 (0.86,2.78) | 5.13 (2.16,12.85)*** | 2.21 (1.65,2.99)*** |
| **NWCO** | 1.57 (1.01,2.42)* | 2.50 (1.27,4.71)** | 0.90 (0.59,1.37) | 3.33 (1.72,7.27)*** | 1.96 (0.69,4.90) |
| **AWCO** | 4.14 (3.07,5.65)*** | 5.30 (3.92,7.21)*** | 3.14 (2.29,4.37)*** | 8.98 (4.74,19.30)*** | 4.78 (3.56,6.45)*** |
| **Dyslipidaemia** |  |  |  |  |  |
| **NWNC** | Ref | Ref | Ref | Ref | Ref |
| **AWNC** | 0.85 (0.45,1.68) | 1.38 (0.79,2.48) | 0.64 (0.29,1.54) | 1.03 (0.35,3.40) | 1.44 (0.82,2.61) |
| **NWCO** | 1.93 (0.84,5.26) | 1.21 (0.35,7.63) | 1.43 (0.72,2.94) | 1.56 (0.72,3.20) | 1.32 (0.26,24.04) |
| **AWCO** | 3.44 (1.79,6.97)*** | 3.05 (1.40,7.61)** | 2.73 (1.45,5.21)** | 2.79 (1.27,5.81)** | 2.68 (1.28,6.32)* |
| *Note. IDF International Diabetes Federation, WHO World Health Organisation, NCEP ATP III National Cholesterol Education Program Adult Treatment Panel III, WC Waist Circumference, WHtR Waist-Height Ratio, WHR Waist-Hip Ratio, VO Visceral Obesity, NWNC Normal Weight No Central Obesity, AWNC Abnormal Weight No Central Obesity, NWCO Normal Weight Central Obesity, AWCO Abnormal Weight Central Obesity.* | | | | | |
| *Adjusted for age, education, income, job skill level, tobacco use, and alcohol consumption.* | | | | | |
| * p < 0.05, ** p < 0.01, *** p<0.001 | |  |  |  |  |

| **Table S5** | |  |  |  |  |  |
| --- | --- | --- | --- | --- | --- | --- |
| *TS5. Association between Types of Dual Metric Obesity Classification and Types of Dyslipidemia stratified by sex.* | | | | | | |
| **Variable** | **Odds Ratio (95% Confidence Interval)** | | | | | |
|  | **WC** | | | **WHTR** | **WHR** | **VFP** |
|  | **IDF** | **WHO** | **NCEP ATP III** |  |  |  |
| **Males** | | | | | | |
| **Elevated TC** |  |  |  |  |  |  |
| **NWNC** | Ref | Ref | Ref | Ref | Ref | Ref |
| **AWNC** | 1.39 (1.02,1.89)* | 1.42 (1.09,1.86)* | 1.38 (1.08,1.76)** | 1.74 (0.84,2.30) | 1.12 (0.47,2.47) | 2.94 (1.15,7.22)* |
| **NWCO** | 1.91 (0.90,3.88) | 2.77 (0.82,8.51) | a | 1.59 (1.09,2.30)* | 1.42 (0.93,2.23) | 1.37 (0.91,2.10) |
| **AWCO** | 1.43 (1.09,1.88)* | 1.35 (0.99,1.83) | 1.20 (0.68,2.06) | 1.65 (1.23,2.23)*** | 1.81 (1.21,2.77)** | 1.67 (1.14,2.48)** |
| **Elevated TG** |  |  |  |  |  |  |
| **NWNC** | Ref | Ref | Ref | Ref | Ref | Ref |
| **AWNC** | 2.43 (1.86,3.18)*** | 2.63 (2.08,3.32)*** | 2.75 (2.22,3.42)*** | 1.84 (0.98,3.40) | 3.20 (1.62,6.29)*** | 2.70 (1.13,6.48)* |
| **NWCO** | 1.79 (0.90,3.54) | 3.10 (1.01,10.48) | a | 2.20 (1.60,3.04)*** | 3.14 (2.13,4.70)*** | 1.83 (1.29,2.62)*** |
| **AWCO** | 3.22 (2.53,4.10)*** | 3.31 (2.51,4.37)*** | 3.29 (2.03,5.45)*** | 4.05 (3.14,5.24)*** | 6.91 (4.77,10.24)*** | 4.33 (3.12,6.07)*** |
| **Elevated LDL** |  |  |  |  |  |  |
| **NWNC** | Ref | Ref | Ref | Ref | Ref | Ref |
| **AWNC** | 1.06 (0.77,1.45) | 1.14 (0.87,1.49) | 1.14 (0.89,1.46) | 1.84 (0.92,3.50) | 1.15 (0.48,2.55) | 3.65 (1.46,8.91)** |
| **NWCO** | 2.13 (1.02,4.27)* | 2.61 (0.78,8.00) | a | 1.32 (0.92,1.91) | 1.52 (0.99,2.37) | 1.53 (1.01,2.35)* |
| **AWCO** | 1.29 (0.98,1.70) | 1.23 (0.90,1.68) | 1.17 (0.66,2.00) | 1.26 (0.94,1.70) | 1.59 (1.06,2.43)* | 1.51 (1.03,2.26)* |
| **Decreased HDL** |  |  |  |  |  |  |
| **NWNC** | Ref | Ref | Ref | Ref | Ref | Ref |
| **AWNC** | 1.27 (0.97,1.68) | 1.54 (1.21,1.97)*** | 1.69 (1.36,2.12)*** | 0.68 (0.37,1.25) | 1.35 (0.71,2.64) | 0.93 (0.39,2.22) |
| **NWCO** | 1.54 (0.76,3.33) | 2.09 (0.63,9.41) | a | 1.42 (1.03,1.95)* | 1.43 (1.01,2.01)* | 1.66 (1.18,2.32)** |
| **AWCO** | 2.31 (1.79,2.99)*** | 2.34 (1.74,3.18)*** | 2.93 (1.67,5.47)*** | 2.17(1.68,2.79)*** | 2.35 (1.69,3.26)*** | 2.57 (1.87,3.52)*** |
| **Females** | | | | | | |
| **Elevated TC** |  |  |  |  |  |  |
| **NWNC** | Ref | Ref | Ref | Ref | Ref | Ref |
| **AWNC** | 1.26 (0.91,1.75) | 1.26 (0.91,1.75) | 1.34 (1.06,1.70)* | 1.37 (0.86,2.16) | 1.67 (0.93,3.00) | 1.22 (0.96,1.55) |
| **NWCO** | 1.5 (0.91,1.75) | 1.05 (0.75,1.47) | 1.28 (0.71,2.28) | 1.23 (0.91,1.66) | 1.49 (1.02,2.19)* | 0.61 (0.22,1.47) |
| **AWCO** | 1.47 (1.16,1.86)** | 1.47 (1.16,1.86)** | 1.54 (1.19,1.99)*** | 1.56 (1.21,2.02)*** | 1.94 (1.35,2.83)*** | 1.61 (1.25,2.07)*** |
| **Elevated TG** |  |  |  |  |  |  |
| **NWNC** | Ref | Ref | Ref | Ref | Ref | Ref |
| **AWNC** | 1.31 (0.95,1.81) | 1.31 (0.95,1.81) | 1.54 (1.22,1.94)*** | 1.25 (0.79,1.97) | 1.62 (0.88,2.94) | 1.70 (1.35,2.14)*** |
| **NWCO** | 1.11 (0.80,1.55) | 1.11 (0.80,1.55) | 0.96 (0.53,1.71) | 1.23 (0.91,1.65) | 2.00 (1.37,2.97)*** | 0.83 (0.33,1.90) |
| **AWCO** | 2.39 (1.89,3.02)*** | 2.39 (1.89,3.02)*** | 2.92 (2.26,3.79)*** | 2.37 (1.84,3.05)*** | 3.73 (2.57,5.50)*** | 2.49 (1.954,3.22)*** |
| **Elevated LDL** |  |  |  |  |  |  |
| **NWNC** | Ref | Ref | Ref | Ref | Ref | Ref |
| **AWNC** | 1.34 (0.97,1.86) | 1.34 (0.97,1.86) | 1.31 (1.03,1.66)* | 1.38 (0.87,2.19) | 1.74 (0.97,3.12) | 1.17 (0.92,1.49) |
| **NWCO** | 1.13 (0.81,1.59) | 1.14 (0.81,1.59) | 1.35 (0.75,2.39) | 1.32 (0.98,1.80) | 1.40 (0.96,2.07) | 0.38 (0.11,1.01) |
| **AWCO** | 1.34 (1.06,1.70)* | 1.34 (1.06,1.70)* | 1.34 (1.04,1.74)* | 1.49 (1.16,1.94)** | 1.69 (1.17,2.47)** | 1.39 (1.08,1.79)* |
| **Decreased HDL** |  |  |  |  |  |  |
| **NWNC** | Ref | Ref | Ref | Ref | Ref | Ref |
| **AWNC** | 1.28 (0.88,1.89) | 1.28 (0.88,1.89) | 1.57 (1.18,2.10)** | 0.88 (0.54,1.49) | 1.84 (0.95,2.09) | 1.58 (1.19,2.11)** |
| **NWCO** | 1.20 (0.82,1.78) | 1.20 (0.82,1.78) | 0.79 (0.43,1.52) | 0.92 (0.66,1.29) | 1.42 (0.95,2.09) | 1.10 (0.45,3.08) |
| **AWCO** | 2.15 (1.61,2.88)*** | 2.15 (1.61,2.88)*** | 2.08 (1.50,2.94)*** | 1.90 (1.40,2.57)*** | 2.39 (1.61,3.51)*** | 2.15 (1.55,3.02)*** |
| *Note. IDF International Diabetes Federation, WHO World Health Organisation, NCEP ATP III National Cholesterol Education Program Adult Treatment Panel III, WC Waist Circumference, WHtR Waist-Height Ratio, WHR Waist-Hip Ratio, VO Visceral Obesity, NWNC Normal Weight No Central Obesity, AWNC Abnormal Weight No Central Obesity, NWCO Normal Weight Central Obesity, AWCO Abnormal Weight Central Obesity, TC Total Cholesterol, LDL Low Density Lipoprotein, TG Triglyceride, HDL High Density Lipoprotein* | | | | | | |
| *Adjusted for age, education, income, job skill level, tobacco use, and alcohol consumption.* | | | | | | |
| * p < 0.05, ** p < 0.01, *** p<0.001 | | |  |  |  |  |

Supplementary Section 3

### *SS3. Cardiometabolic Marker Levels Based on Dual Metric Obesity Criteria Groups*

| **Table S6** |  |  |  |  |  |  |
| --- | --- | --- | --- | --- | --- | --- |
| *TS6. Comparison of means of cardiometabolic markers between dual metric obesity criteria groups based on waist-hip ratio (WHR).* | | | | | | |
| **Cardiometabolic Marker** | **NWNC^a^** | **AWNC^a^** | **NWCO^a^** | **AWCO^a^** | **p-value^b^** | **Post Hoc Analysis^c^** |
| **Females** | | | | | | |
| **SBP** | 116.49 [113.94, 119.04] | 123.24 [119.40, 127.09] | 118.34 [116.98, 119.70] | 123.93 [122.82, 125.03] | <0.001 | NWCO-AWCO*, NWNC-AWCO*, NWNC-AWNC* |
| **DBP** | 73.13 [71.60, 74.67] | 73.51 [71.20, 75.82] | 72.33 [71.51, 73.15] | 76.65 [75.99, 77.31] | <0.001 | NWCO-AWCO*, NWNC-AWCO* |
| **FBG** | 94.57 [88.77, 100.37] | 108.32 [99.58, 117.06] | 104.59 [101.49, 107.68] | 111.27 [108.76, 113.79] | <0.001 | NWCO-AWCO*, NWNC-AWCO*, NWNC-AWNC* |
| **HbA1C** | 5.64 [5.42, 5.86] | 6.32 [5.99, 6.65] | 6.16 [6.05, 6.28] | 6.51 [6.42, 6.61] | <0.001 | NWCO-AWCO*, NWNC-AWCO*, NWNC-AWNC* |
| **Fasting Insulin** | 6.03 [5.01, 7.06] | 7.85 [6.30, 9.39] | 7.03 [6.49, 7.58] | 10.73 [10.28, 11.17] | <0.001 | AWNC-AWCO*, NWCO-AWCO*, NWNC-AWCO* |
| **TG** | 125.26 [111.59, 138.93] | 137.53 [116.94, 158.13] | 155.24 [147.96, 162.53] | 177.84 [171.92, 183.76] | <0.001 | AWNC-AWCO*, NWCO-AWCO*, NWNC-AWCO* |
| **LDL** | 108.69 [103.54, 113.83] | 117.21 [109.46, 124.97] | 119.08 [116.34, 121.82] | 121.54 [119.31, 123.77] | <0.001 | NWNC-AWCO*, NWNC-NWCO* |
| **HDL** | 44.54 [43.00, 46.07] | 40.99 [38.67, 43.31] | 42.77 [41.95, 43.59] | 40.62 [39.95, 41.28] | <0.001 | NWCO-AWCO*, NWNC-AWCO* |
| **Males** | | | | | | |
| **SBP** | 121.24 [118.95, 123.53] | 124.90 [120.35, 129.44] | 123.30 [121.90, 124.71] | 126.03 [124.97, 127.08] | <0.001 | NWCO-AWCO*, NWNC-AWCO* |
| **DBP** | 74.46 [73.07, 75.85] | 74.49 [71.73, 77.25] | 76.13 [75.28, 76.98] | 77.89 [77.25, 78.53] | <0.001 | NWCO-AWCO*, NWNC-AWCO* |
| **FBG** | 99.97 [92.46, 107.48] | 108.48 [93.58, 123.38] | 116.67 [112.07, 121.27] | 121.33 [117.88, 124.79] | <0.001 | NWNC-AWCO*, NWNC-NWCO* |
| **HbA1C** | 5.90 [5.64, 6.16] | 5.92 [5.40, 6.43] | 6.61 [6.46, 6.77] | 6.89 [6.77, 7.01] | <0.001 | AWNC-AWCO*, NWCO-AWCO*, NWNC-AWCO*, NWNC-NWCO* |
| **Fasting Insulin** | 4.73 [3.67, 5.78] | 7.02 [4.93, 9.11] | 6.15 [5.50, 6.79] | 11.30 [10.81, 11.78] | <0.001 | AWNC-AWCO*, NWCO-AWCO*, NWNC-AWCO* |
| **TG** | 120.63 [105.34, 135.93] | 148.97 [118.62, 179.32] | 163.82 [154.46, 173.18] | 204.17 [197.13, 211.21] | <0.001 | AWNC-AWCO*, NWCO-AWCO*, NWNC-AWCO*, NWNC-NWCO* |
| **LDL** | 99.69 [95.20, 104.18] | 105.96 [97.05, 114.87] | 105.67 [102.92, 108.42] | 108.52 [106.46, 110.59] | 0.005 | NWNC-AWCO* |
| **HDL** | 39.10 [37.77, 40.43] | 35.34 [32.70, 37.98] | 37.77 [36.96, 38.59] | 35.24 [34.63, 35.85] | <0.001 | NWCO-AWCO*, NWNC-AWCO* |
| *Note. SBP Systolic Blood Pressure, DBP Diastolic Blood Pressure, FBG Fasting Blood Glucose, HbA1C Glycated Hemoglobin, TG Triglyceride, LDL Low Density Lipoprotein, HDL High Density Lipoprotein.* | | | | | | |
| *^a^ - mean [95% CI]* |  |  |  |  |  |  |
| *^b^ - ANCOVA adjusted for age, education, income, job skill level, tobacco use, and alcohol consumption.* | | | |  |  |  |
| *^c^ - post hoc Tukey analysis* | |  |  |  |  |  |
| ** P<0.05* |  |  |  |  |  |  |

| **Table S7** |  |  |  |  |  |  |
| --- | --- | --- | --- | --- | --- | --- |
| *TS7. Comparison of means of cardiometabolic markers between dual metric obesity criteria groups based on Waist-Height ratio (WHtR).* | | | | | | |
| **Cardiometabolic Marker** | **NWNC^a^** | **AWNC^a^** | **NWCO^a^** | **AWCO^a^** | **p-value^b^** | **Post Hoc Analysis^c^** |
| **Females** | | | | | | |
| **SBP** | 116.84 [115.19, 118.48] | 120.30 [117.02, 123.58] | 119.16 [117.41, 120.91] | 124.29 [123.17, 125.41] | <0.001 | NWCO-AWCO*, NWNC-AWCO*, NWNC-AWNC* |
| **DBP** | 72.17 [71.19, 73.16] | 72.57 [70.60, 74.55] | 72.88 [71.84, 73.93] | 76.86 [76.18, 77.53] | <0.001 | AWNC-AWCO*, NWCO-AWCO*, NWNC-AWCO* |
| **FBG** | 102.46 [98.70, 106.21] | 104.59 [97.10, 112.09] | 102.27 [98.28, 106.26] | 111.80 [109.24, 114.35] | <0.001 | NWCO-AWCO*, NWNC-AWCO* |
| **HbA1C** | 6.01 [5.87, 6.15] | 6.21 [5.93, 6.49] | 6.09 [5.94, 6.24] | 6.53 [6.44, 6.63] | <0.001 | NWCO-AWCO*, NWNC-AWCO* |
| **Fasting Insulin** | 6.41 [5.75, 7.08] | 7.83 [6.51, 9.15] | 7.26 [6.56, 7.96] | 10.82 [10.37, 11.27] | <0.001 | AWNC-AWCO*, NWCO-AWCO*, NWNC-AWCO* |
| **TG** | 140.86 [132.01, 149.70] | 138.38 [120.73, 156.02] | 157.33 [147.94, 166.72] | 178.99 [172.98, 185.01] | <0.001 | AWNC-AWCO*, NWCO-AWCO*, NWNC-AWCO* |
| **LDL** | 114.39 [111.05, 117.72] | 119.22 [112.56, 125.87] | 119.47 [115.93, 123.01] | 121.44 [119.18, 123.71] | 0.008 | NWNC-AWCO* |
| **HDL** | 43.54 [42.55, 44.53] | 43.00 [41.02, 44.98] | 42.74 [41.69, 43.79] | 40.37 [39.70, 41.05] | <0.001 | NWCO-AWCO*, NWNC-AWCO* |
| **Males** | | | | | | |
| **SBP** | 121.54 [119.99, 123.08] | 123.13 [118.54, 127.71] | 124.53 [122.65, 126.41] | 126.12 [125.07, 127.17] | <0.001 | NWNC-AWCO* |
| **DBP** | 75.02 [74.08, 75.95] | 73.33 [70.55, 76.12] | 76.65 [75.51, 77.79] | 77.95 [77.31, 78.59] | <0.001 | AWNC-AWCO*, NWNC-AWCO* |
| **FBG** | 107.36 [102.29, 112.44] | 102.43 [87.36, 117.49] | 119.18 [113.00, 125.36] | 121.64 [118.18, 125.09] | <0.001 | NWNC-AWCO*, NWNC-NWCO* |
| **HbA1C** | 6.25 [6.07, 6.42] | 6.03 [5.51, 6.56] | 6.68 [6.46, 6.89] | 6.88 [6.76, 7.00] | <0.001 | AWNC-AWCO*, NWNC-AWCO*, NWNC-NWCO* |
| **Fasting Insulin** | 4.92 [4.21, 5.63] | 7.50 [5.39, 9.61] | 7.01 [6.15, 7.88] | 11.27 [10.78, 11.75] | <0.001 | AWNC-AWCO*, NWCO-AWCO*, NWNC-AWCO*, NWNC-NWCO* |
| **TG** | 139.28 [128.90, 149.65] | 165.85 [135.05, 196.64] | 171.01 [158.37, 183.65] | 203.22 [196.16, 210.29] | <0.001 | NWCO-AWCO*, NWNC-AWCO*, NWNC-NWCO* |
| **LDL** | 102.61 [99.58, 105.64] | 118.86 [109.87, 127.86] | 106.15 [102.46, 109.85] | 107.84 [105.78, 109.90] | 0.002 | NWNC-AWCO* |
| **HDL** | 38.68 [37.78, 39.58] | 38.19 [35.52, 40.85] | 37.33 [36.23, 38.42] | 35.09 [34.48, 35.70] | <0.001 | NWCO-AWCO*, NWNC-AWCO* |
| *Note. SBP Systolic Blood Pressure, DBP Diastolic Blood Pressure, FBG Fasting Blood Glucose, HbA1C Glycated Hemoglobin, TG Triglyceride, LDL Low Density Lipoprotein, HDL High Density Lipoprotein.* | | | | | | |
| *^a^ - mean [95% CI]* |  |  |  |  |  |  |
| *^b^ - ANCOVA adjusted for age, education, income, job skill level, tobacco use, and alcohol consumption.* | | | |  |  |  |
| *^c^ - post hoc Tukey analysis* | |  |  |  |  |  |
| ** P<0.05* |  |  |  |  |  |  |

| **Table S8** |  |  |  |  |  |  |
| --- | --- | --- | --- | --- | --- | --- |
| *TS8. Comparison of means of cardiometabolic markers between dual metric obesity criteria groups based on visceral fat percentage (VFP).* | | | | | | |
| **Cardiometabolic Marker** | **NWNC^a^** | **AWNC^a^** | **NWCO^a^** | **AWCO^a^** | **p-value^b^** | **Post Hoc Analysis^c^** |
| **Females** | | | | | | |
| **SBP** | 117.82 [116.61, 119.03] | 121.42 [120.03, 122.82] | 121.20 [114.64, 127.76] | 127.16 [125.55, 128.77] | <0.001 | AWNC-AWCO*, NWNC-AWCO*, NWNC-AWNC* |
| **DBP** | 72.47 [71.74, 73.20] | 74.99 [74.15, 75.83] | 73.60 [69.64, 77.56] | 78.31 [77.34, 79.28] | <0.001 | AWNC-AWCO*, NWNC-AWCO*, NWNC-AWNC* |
| **FBG** | 102.48 [99.70, 105.26] | 108.82 [105.63, 112.02] | 99.12 [84.07, 114.18] | 114.02 [110.33, 117.72] | <0.001 | NWNC-AWCO*, NWNC-AWNC* |
| **HbA1C** | 6.05 [5.95, 6.16] | 6.36 [6.24, 6.48] | 5.92 [5.36, 6.48] | 6.68 [6.54, 6.82] | <0.001 | AWNC-AWCO*, NWNC-AWCO*, NWNC-AWNC* |
| **Fasting Insulin** | 6.73 [6.24, 7.22] | 9.36 [8.80, 9.92] | 9.25 [6.61, 11.89] | 12.04 [11.39, 12.69] | <0.001 | AWNC-AWCO*, NWNC-AWCO*, NWNC-AWNC* |
| **TG** | 148.99 [142.40, 155.57] | 169.49 [161.92, 177.06] | 137.45 [101.78, 173.12] | 181.81 [173.06, 190.57] | <0.001 | NWNC-AWCO*, NWNC-AWNC* |
| **LDL** | 117.39 [114.93, 119.85] | 119.05 [116.22, 121.88] | 98.80 [85.46, 112.14] | 124.10 [120.82, 127.37] | <0.001 | NWCO-AWCO*, NWNC-AWCO*, NWCO-AWNC*, NWNC-NWCO* |
| **HDL** | 43.23 [42.50, 43.97] | 41.03 [40.18, 41.87] | 41.18 [37.19, 45.16] | 40.14 [39.16, 41.12] | <0.001 | NWNC-AWCO*, NWNC-AWNC* |
| **Males** | | | | | | |
| **SBP** | 119.86 [117.71, 122.02] | 127.50 [121.02, 133.98] | 124.01 [122.58, 125.45] | 125.93 [124.89, 126.97] | <0.001 | NWNC-AWCO*, NWNC-NWCO* |
| **DBP** | 74.48 [73.17, 75.79] | 76.67 [72.72, 80.61] | 76.20 [75.33, 77.07] | 77.75 [77.12, 78.38] | <0.001 | NWCO-AWCO*, NWNC-AWCO* |
| **FBG** | 109.02 [101.91, 116.14] | 127.48 [106.09, 148.86] | 113.49 [108.76, 118.22] | 120.50 [117.08, 123.93] | 0.008 | NWNC-AWCO* |
| **HbA1C** | 6.29 [6.04, 6.53] | 7.29 [6.54, 8.03] | 6.48 [6.32, 6.64] | 6.82 [6.71, 6.94] | <0.001 | NWCO-AWCO*, NWNC-AWCO* |
| **Fasting Insulin** | 4.68 [3.69, 5.68] | 8.61 [5.61, 11.61] | 6.23 [5.57, 6.90] | 11.14 [10.66, 11.62] | <0.001 | NWCO-AWCO*, NWNC-AWCO* |
| **TG** | 137.59 [123.05, 152.12] | 168.81 [125.10, 212.52] | 158.44 [148.78, 168.10] | 202.19 [195.19, 209.19] | <0.001 | NWCO-AWCO*, NWNC-AWCO* |
| **LDL** | 100.24 [96.01, 104.47] | 115.65 [102.92, 128.38] | 105.72 [102.90, 108.53] | 108.20 [106.17, 110.24] | 0.004 | NWNC-AWCO* |
| **HDL** | 39.75 [38.50, 41.00] | 39.02 [35.26, 42.78] | 37.42 [36.59, 38.25] | 35.15 [34.55, 35.75] | <0.001 | NWCO-AWCO*, NWNC-AWCO*, NWNC-NWCO* |
| *Note. SBP Systolic Blood Pressure, DBP Diastolic Blood Pressure, FBG Fasting Blood Glucose, HbA1C Glycated Haemoglobin, TG Triglyceride, LDL Low Density Lipoprotein, HDL High Density Lipoprotein.* | | | | | | |
| *^a^ - mean [95% CI]* |  |  |  |  |  |  |
| *^b^ - ANCOVA adjusted for age, education, income, job skill level, tobacco use, and alcohol consumption.* | | | |  |  |  |
| *^c^ - post hoc Tukey analysis* | |  |  |  |  |  |
| ** P<0.05* |  |  |  |  |  |  |

| **Table S9** |  |  |  |  |  |  |
| --- | --- | --- | --- | --- | --- | --- |
| *TS9. Comparison of means of cardiometabolic markers between dual metric obesity criteria groups based on waist circumference using the Internation Diabetes Federation (IDF) criteria.* | | | | | | |
| **Cardiometabolic Marker** | **NWNC^a^** | **AWNC^a^** | **NWCO^a^** | **AWCO^a^** | **p-value^b^** | **Post Hoc Analysis^c^** |
| **Females** | | | | | | |
| **SBP** | 117.29 [115.89, 118.69] | 120.76 [118.56, 122.95] | 119.66 [117.36, 121.96] | 124.82 [123.61, 126.03] | <0.001 | AWNC-AWCO*, NWCO-AWCO*, NWNC-AWCO*, NWNC-AWNC* |
| **DBP** | 72.37 [71.53, 73.21] | 73.78 [72.46, 75.10] | 72.88 [71.49, 74.26] | 77.21 [76.48, 77.93] | <0.001 | AWNC-AWCO*, NWCO-AWCO*, NWNC-AWCO* |
| **FBG** | 100.91 [97.71, 104.10] | 105.73 [100.71, 110.74] | 106.32 [101.07, 111.58] | 112.66 [109.90, 115.41] | <0.001 | NWNC-AWCO* |
| **HbA1C** | 5.97 [5.85, 6.09] | 6.25 [6.06, 6.44] | 6.26 [6.07, 6.46] | 6.58 [6.47, 6.68] | <0.001 | AWNC-AWCO*, NWCO-AWCO*, NWNC-AWCO* |
| **Fasting Insulin** | 6.55 [5.99, 7.10] | 7.72 [6.84, 8.60] | 7.53 [6.61, 8.45] | 11.35 [10.87, 11.83] | <0.001 | AWNC-AWCO*, NWCO-AWCO*, NWNC-AWCO* |
| **TG** | 144.41 [136.88, 151.94] | 149.43 [137.62, 161.24] | 159.95 [147.57, 172.33] | 182.41 [175.92, 188.91] | <0.001 | AWNC-AWCO*, NWCO-AWCO*, NWNC-AWCO* |
| **LDL** | 116.01 [113.17, 118.85] | 119.05 [114.59, 123.51] | 118.86 [114.19, 123.53] | 121.86 [119.41, 124.31] | 0.025 | NWNC-AWCO* |
| **HDL** | 43.35 [42.50, 44.19] | 42.05 [40.73, 43.38] | 42.68 [41.28, 44.07] | 40.22 [39.49, 40.95] | <0.001 | NWCO-AWCO*, NWNC-AWCO* |
| **Males** | | | | | | |
| **SBP** | 122.55 [121.32, 123.78] | 124.61 [122.96, 126.26] | 126.22 [120.99, 131.44] | 126.83 [125.52, 128.14] | <0.001 | NWNC-AWCO* |
| **DBP** | 75.63 [74.89, 76.37] | 75.98 [74.98, 76.98] | 76.49 [73.32, 79.65] | 78.81 [78.02, 79.61] | <0.001 | AWNC-AWCO*, NWNC-AWCO* |
| **FBG** | 111.65 [107.61, 115.69] | 115.92 [110.49, 121.35] | 120.64 [103.44, 137.85] | 123.67 [119.36, 127.98] | 0.001 | NWNC-AWCO* |
| **HbA1C** | 6.40 [6.26, 6.54] | 6.56 [6.37, 6.75] | 6.88 [6.28, 7.48] | 7.01 [6.86, 7.16] | <0.001 | AWNC-AWCO*, NWNC-AWCO* |
| **Fasting Insulin** | 5.54 [4.99, 6.09] | 8.14 [7.40, 8.88] | 9.66 [7.32, 12.01] | 12.93 [12.34, 13.52] | <0.001 | AWNC-AWCO*, NWCO-AWCO*, NWNC-AWCO*, NWNC-NWCO* |
| **TG** | 151.23 [142.97, 159.50] | 189.58 [178.46, 200.70] | 166.84 [131.64, 202.04] | 208.77 [199.95, 217.59] | <0.001 | AWNC-AWCO*, NWNC-AWCO*, NWNC-AWNC* |
| **LDL** | 103.69 [101.28, 106.10] | 108.27 [105.03, 111.51] | 110.33 [100.07, 120.60] | 108.47 [105.89, 111.04] | 0.028 | NWNC-AWCO* |
| **HDL** | 38.15 [37.44, 38.86] | 36.68 [35.73, 37.64] | 37.89 [34.86, 40.91] | 34.34 [33.58, 35.10] | <0.001 | AWNC-AWCO*, NWNC-AWCO* |
| *Note. SBP Systolic Blood Pressure, DBP Diastolic Blood Pressure, FBG Fasting Blood Glucose, HbA1C Glycated Haemoglobin, TG Triglyceride, LDL Low Density Lipoprotein, HDL High Density Lipoprotein.* | | | | | | |
| *^a^ - mean [95% CI]* |  |  |  |  |  |  |
| *^b^ - ANCOVA adjusted for age, education, income, job skill level, tobacco use, and alcohol consumption.* | | | |  |  |  |
| *^c^ - post hoc Tukey analysis* | |  |  |  |  |  |
| ** P<0.05* |  |  |  |  |  |  |

| **Table S10** |  |  |  |  |  |  |
| --- | --- | --- | --- | --- | --- | --- |
| *TS10. Comparison of means of cardiometabolic markers between dual metric obesity criteria groups based on waist circumference using the World Health Organization (WHO) criteria.* | | | | | | |
| **Cardiometabolic Marker** | **NWNC^a^** | **AWNC^a^** | **NWCO^a^** | **AWCO^a^** | **p-value^b^** | **Post Hoc Analysis^c^** |
| **Females** | | | | | | |
| **SBP** | 117.29 [115.89, 118.69] | 120.76 [118.56, 122.95] | 119.66 [117.36, 121.96] | 124.82 [123.61, 126.03] | <0.001 | AWNC-AWCO*, NWCO-AWCO*, NWNC-AWCO*, NWNC-AWNC* |
| **DBP** | 72.37 [71.53, 73.21] | 73.78 [72.46, 75.10] | 72.88 [71.49, 74.26] | 77.21 [76.48, 77.93] | <0.001 | AWNC-AWCO*, NWCO-AWCO*, NWNC-AWCO* |
| **FBG** | 100.91 [97.71, 104.10] | 105.73 [100.71, 110.74] | 106.32 [101.07, 111.58] | 112.66 [109.90, 115.41] | <0.001 | NWNC-AWCO* |
| **HbA1C** | 5.97 [5.85, 6.09] | 6.25 [6.06, 6.44] | 6.26 [6.07, 6.46] | 6.58 [6.47, 6.68] | <0.001 | AWNC-AWCO*, NWCO-AWCO*, NWNC-AWCO* |
| **Fasting Insulin** | 6.55 [5.99, 7.10] | 7.72 [6.84, 8.60] | 7.53 [6.61, 8.45] | 11.35 [10.87, 11.83] | <0.001 | AWNC-AWCO*, NWCO-AWCO*, NWNC-AWCO* |
| **TG** | 144.41 [136.88, 151.94] | 149.43 [137.62, 161.24] | 159.95 [147.57, 172.33] | 182.41 [175.92, 188.91] | <0.001 | AWNC-AWCO*, NWCO-AWCO*, NWNC-AWCO* |
| **LDL** | 116.01 [113.17, 118.85] | 119.05 [114.59, 123.51] | 118.86 [114.19, 123.53] | 121.86 [119.41, 124.31] | 0.025 | NWNC-AWCO* |
| **HDL** | 43.35 [42.50, 44.19] | 42.05 [40.73, 43.38] | 42.68 [41.28, 44.07] | 40.22 [39.49, 40.95] | <0.001 | NWCO-AWCO*, NWNC-AWCO* |
| **Males** | | | | | | |
| **SBP** | 122.56 [121.36, 123.77] | 124.98 [123.69, 126.27] | 132.31 [123.50, 141.11] | 127.66 [125.97, 129.34] | <0.001 | NWNC-AWCO*, NWNC-AWNC* |
| **DBP** | 75.62 [74.89, 76.35] | 76.63 [75.85, 77.42] | 78.46 [73.12, 83.80] | 79.58 [78.56, 80.60] | <0.001 | AWNC-AWCO*, NWNC-AWCO* |
| **FBG** | 112.12 [108.14, 116.09] | 119.39 [115.14, 123.65] | 112.18 [83.12, 141.25] | 122.88 [117.31, 128.45] | 0.01 | NWNC-AWCO* |
| **HbA1C** | 6.41 [6.28, 6.55] | 6.72 [6.57, 6.87] | 6.78 [5.77, 7.79] | 7.03 [6.84, 7.23] | <0.001 | NWNC-AWCO*, NWNC-AWNC* |
| **Fasting Insulin** | 5.71 [5.17, 6.25] | 9.23 [8.65, 9.81] | 8.41 [4.45, 12.36] | 14.24 [13.48, 15.00] | <0.001 | AWNC-AWCO*, NWCO-AWCO*, NWNC-AWCO*, NWNC-AWNC* |
| **TG** | 151.59 [143.46, 159.73] | 198.39 [189.68, 207.11] | 176.50 [117.01, 235.99] | 206.42 [195.02, 217.82] | <0.001 | NWNC-AWCO*, NWNC-AWNC* |
| **LDL** | 103.90 [101.53, 106.27] | 108.37 [105.83, 110.91] | 111.37 [94.05, 128.69] | 108.43 [105.11, 111.75] | 0.05 | None |
| **HDL** | 38.18 [37.48, 38.88] | 35.76 [35.01, 36.51] | 35.74 [30.62, 40.86] | 34.37 [33.39, 35.35] | <0.001 | NWNC-AWCO*, NWNC-AWNC* |
| *Note. SBP Systolic Blood Pressure, DBP Diastolic Blood Pressure, FBG Fasting Blood Glucose, HbA1C Glycated Hemoglobin, TG Triglyceride, LDL Low Density Lipoprotein, HDL High Density Lipoprotein.* | | | | | | |
| *^a^ - mean [95% CI]* |  |  |  |  |  |  |
| *^b^ - ANCOVA adjusted for age, education, income, job skill level, tobacco use, and alcohol consumption.* | | | |  |  |  |
| *^c^ - post hoc Tukey analysis* | |  |  |  |  |  |
| ** P<0.05* |  |  |  |  |  |  |

| **Table S11** |  |  |  |  |  |  |
| --- | --- | --- | --- | --- | --- | --- |
| *TS11. Comparison of means of cardiometabolic markers between dual metric obesity criteria groups based on waist circumference using the National Cholesterol Education Program Adult Treatment Panel III (NCEP ATP III) criteria.* | | | | | | |
| **Cardiometabolic Marker** | **NWNC^a^** | **AWNC^a^** | **NWCO^a^** | **AWCO^a^** | **p-value^b^** | **Post Hoc Analysis^c^** |
| **Females** | | | | | | |
| **SBP** | 117.71 [116.47, 118.94] | 122.33 [120.93, 123.72] | 120.96 [116.39, 125.53] | 125.99 [124.36, 127.61] | <0.001 | AWNC-AWCO*, NWNC-AWCO*, NWNC-AWNC* |
| **DBP** | 72.48 [71.74, 73.23] | 75.04 [74.21, 75.88] | 72.88 [70.14, 75.63] | 78.27 [77.29, 79.24] | <0.001 | AWNC-AWCO*, NWCO-AWCO*, NWNC-AWCO*, NWNC-AWNC* |
| **FBG** | 101.12 [98.30, 103.93] | 107.25 [104.09, 110.42] | 119.39 [109.01, 129.77] | 116.21 [112.52, 119.91] | <0.001 | AWNC-AWCO*, NWNC-AWCO*, NWNC-AWNC*, NWNC-NWCO* |
| **HbA1C** | 6.00 [5.90, 6.11] | 6.31 [6.19, 6.43] | 6.64 [6.25, 7.03] | 6.75 [6.62, 6.89] | <0.001 | AWNC-AWCO*, NWNC-AWCO*, NWNC-AWNC*, NWNC-NWCO* |
| **Fasting Insulin** | 6.63 [6.14, 7.12] | 9.11 [8.55, 9.66] | 9.26 [7.44, 11.08] | 12.42 [11.77, 13.06] | <0.001 | AWNC-AWCO*, NWCO-AWCO*, NWNC-AWCO*, NWNC-AWNC*, NWNC-NWCO* |
| **TG** | 146.98 [140.32, 153.65] | 162.79 [155.30, 170.29] | 170.66 [146.08, 195.24] | 191.04 [182.30, 199.79] | <0.001 | AWNC-AWCO*, NWNC-AWCO*, NWNC-AWNC* |
| **LDL** | 116.41 [113.90, 118.93] | 119.54 [116.72, 122.37] | 121.75 [112.47, 131.02] | 123.48 [120.18, 126.78] | 0.009 | NWNC-AWCO* |
| **HDL** | 43.09 [42.34, 43.84] | 41.00 [40.15, 41.84] | 44.19 [41.42, 46.95] | 40.17 [39.19, 41.15] | <0.001 | NWCO-AWCO*, NWNC-AWCO*, NWNC-AWNC* |
| **Males** | | | | | | |
| **SBP** | 122.74 [121.55, 123.94] | 125.64 [124.56, 126.71] | d | 129.63 [126.07, 133.18] | <0.001 | NWNC-AWCO*, NWNC-AWNC* |
| **DBP** | 75.67 [74.95, 76.40] | 77.42 [76.77, 78.07] | d | 81.00 [78.84, 83.16] | <0.001 | AWNC-AWCO*, NWNC-AWCO*, NWNC-AWNC* |
| **FBG** | 112.12 [108.18, 116.06] | 120.06 [116.53, 123.59] | d | 127.46 [115.75, 139.17] | 0.003 | NWNC-AWCO*, NWNC-AWNC* |
| **HbA1C** | 6.42 [6.28, 6.56] | 6.78 [6.66, 6.90] | d | 7.48 [7.07, 7.88] | <0.001 | AWNC-AWCO*, NWNC-AWCO*, NWNC-AWNC* |
| **Fasting Insulin** | 5.76 [5.22, 6.30] | 10.53 [10.04, 11.02] | d | 17.11 [15.49, 18.73] | <0.001 | AWNC-AWCO*, NWNC-AWCO*, NWNC-AWNC* |
| **TG** | 152.05 [143.99, 160.11] | 200.67 [193.44, 207.91] | d | 208.83 [184.84, 232.81] | <0.001 | NWNC-AWCO*, NWNC-AWNC* |
| **LDL** | 104.04 [101.69, 106.38] | 108.40 [106.30, 110.51] | d | 108.26 [101.28, 115.25] | 0.022 | NWNC-AWNC* |
| **HDL** | 38.14 [37.44, 38.83] | 35.41 [34.78, 36.03] | d | 33.47 [31.40, 35.53] | <0.001 | NWNC-AWCO*, NWNC-AWNC* |
| *Note. SBP Systolic Blood Pressure, DBP Diastolic Blood Pressure, FBG Fasting Blood Glucose, HbA1C Glycated Hemoglobin, TG Triglyceride, LDL Low Density Lipoprotein, HDL High Density Lipoprotein.* | | | | | | |
| *^a^ - mean [95% CI]* |  |  |  |  |  |  |
| *^b^ - ANCOVA adjusted for age, education, income, job skill level, tobacco use, and alcohol consumption.* | | | |  |  |  |
| *^c^ - post hoc Tukey analysis* | |  |  |  |  |  |
| *d - no males with normal BMI and abnormal waist circumference according to NCEP ATP III criteria.* | | | | |  |  |
| ** P<0.05* |  |  |  |  |  |  |

**Figure S1**

*FS1. Comparison of means (95% confidence intervals) of cardiometabolic markers according to dual metric obesity criteria categories based on Waist-Height ratio (WHtR) for males and females.*P<0.05 for significant difference of group when compared to normal weight no central obesity group. NWNC Normal Weight No Central Obesity, AWNC Abnormal Weight No Central Obesity, NWCO Normal Weight Central Obesity, AWCO Abnormal Weight Central Obesity*

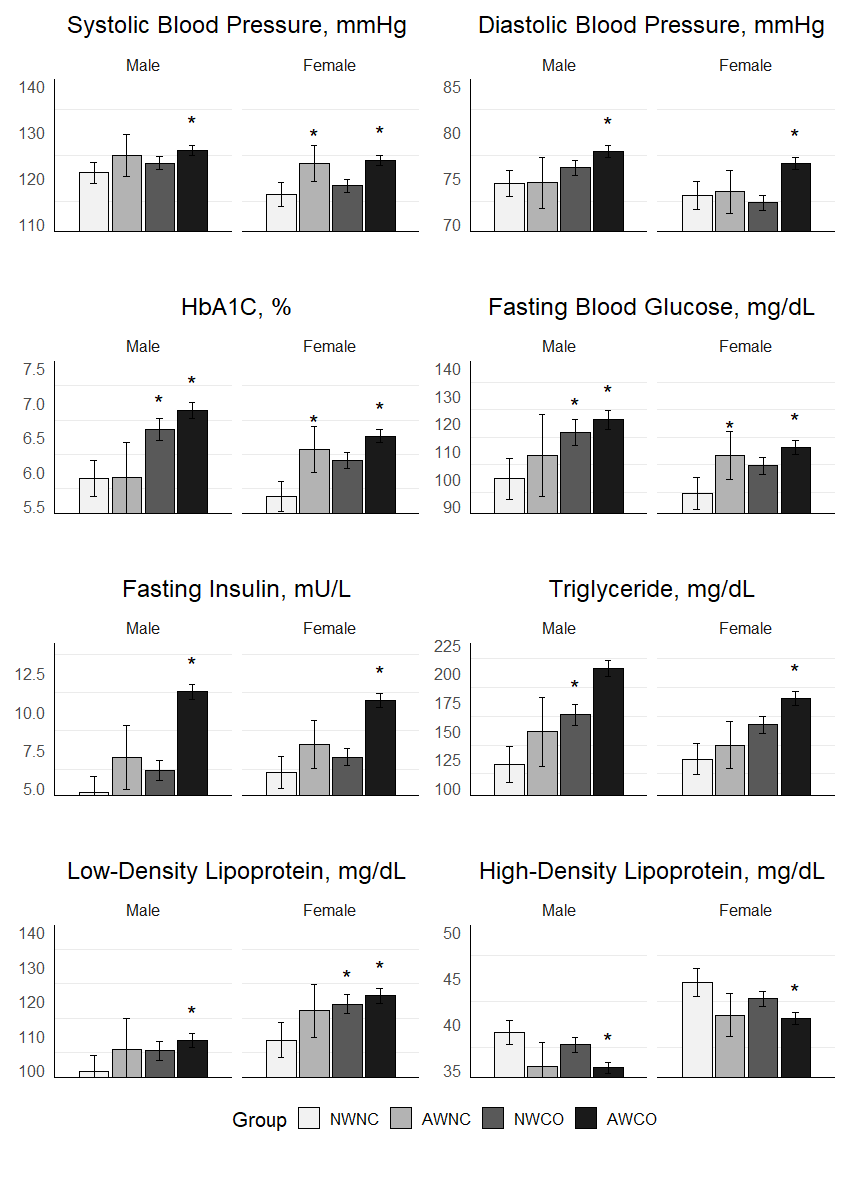

**Figure S2**

*FS2. Comparison of means (95% confidence intervals) of cardiometabolic markers according to dual metric obesity criteria categories based on visceral fat percentage (VFP) for males and females.*P<0.05 for significant difference of group when compared to normal weight no central obesity group. NWNC Normal Weight No Central Obesity, AWNC Abnormal Weight No Central Obesity, NWCO Normal Weight Central Obesity, AWCO Abnormal Weight Central Obesity*
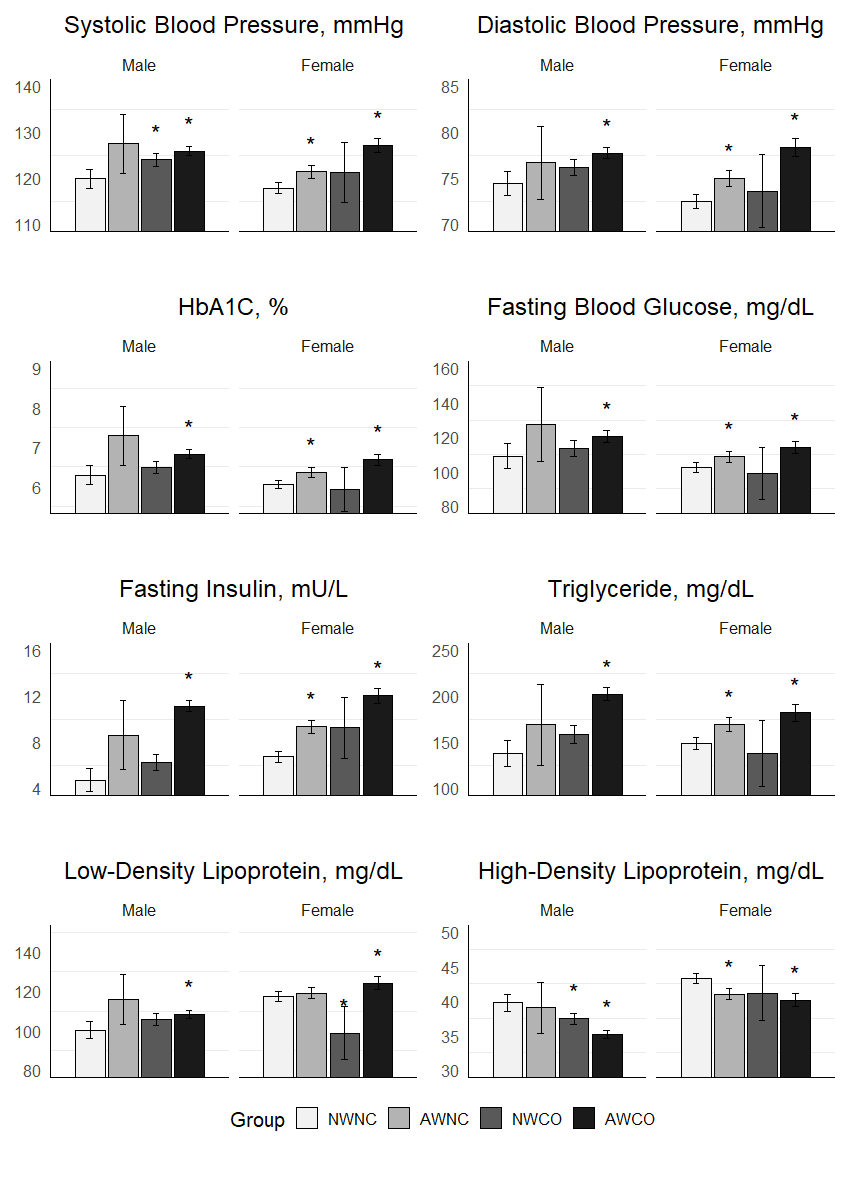

**Figure S3**

*FS3. Comparison of means (95% confidence intervals) of cardiometabolic markers according to dual metric obesity criteria categories based on waist circumference (WC) using International Diabetic Federation (IDF) cutoffs for males and females.*P<0.05 for significant difference of group when compared to normal weight no central obesity group. NWNC Normal Weight No Central Obesity, AWNC Abnormal Weight No Central Obesity, NWCO Normal Weight Central Obesity, AWCO Abnormal Weight Central Obesity*

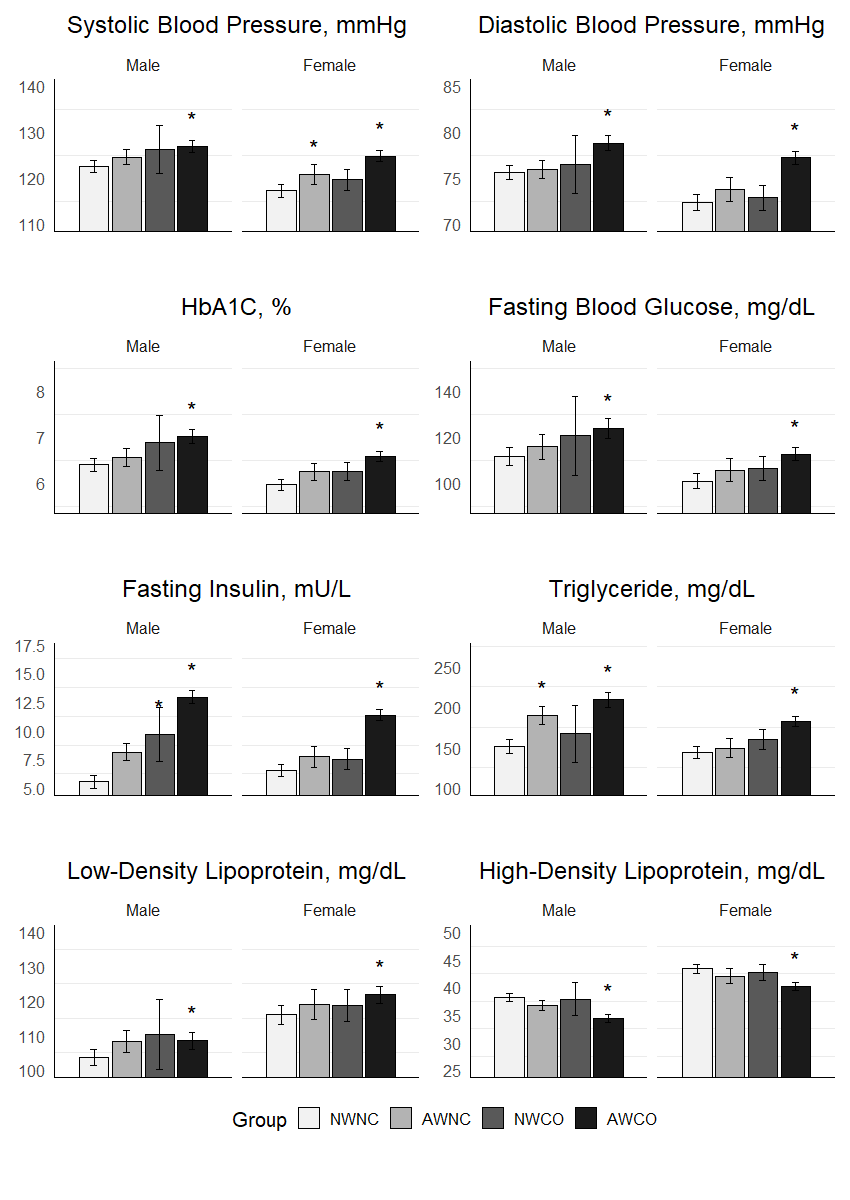

**Figure S4**

*FS4. Comparison of means (95% confidence intervals) of cardiometabolic markers according to dual metric obesity criteria categories based on waist circumference (WC) using World Health Organization (WHO) cutoffs for males and females.*P<0.05 for significant difference of group when compared to normal weight no central obesity group. NWNC Normal Weight No Central Obesity, AWNC Abnormal Weight No Central Obesity, NWCO Normal Weight Central Obesity, AWCO Abnormal Weight Central Obesity*

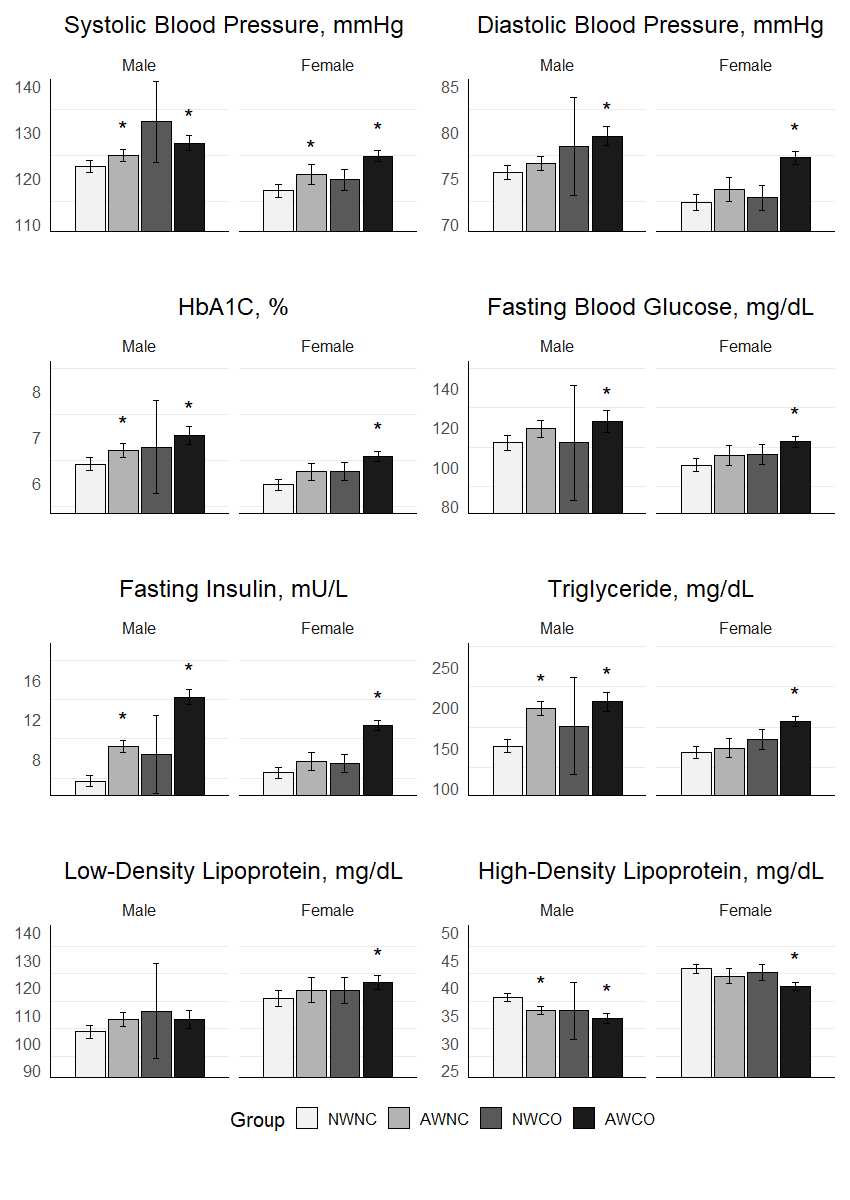

**Figure S5**

*FS5. Comparison of means (95% confidence intervals) of cardiometabolic markers according to dual metric obesity criteria categories based on waist circumference (WC) using Adult Treatment Program III (ATP III) cutoffs for males and females.*P<0.05 for significant difference of group when compared to normal weight no central obesity group. NWNC Normal Weight No Central Obesity, AWNC Abnormal Weight No Central Obesity, NWCO Normal Weight Central Obesity, AWCO Abnormal Weight Central Obesity*

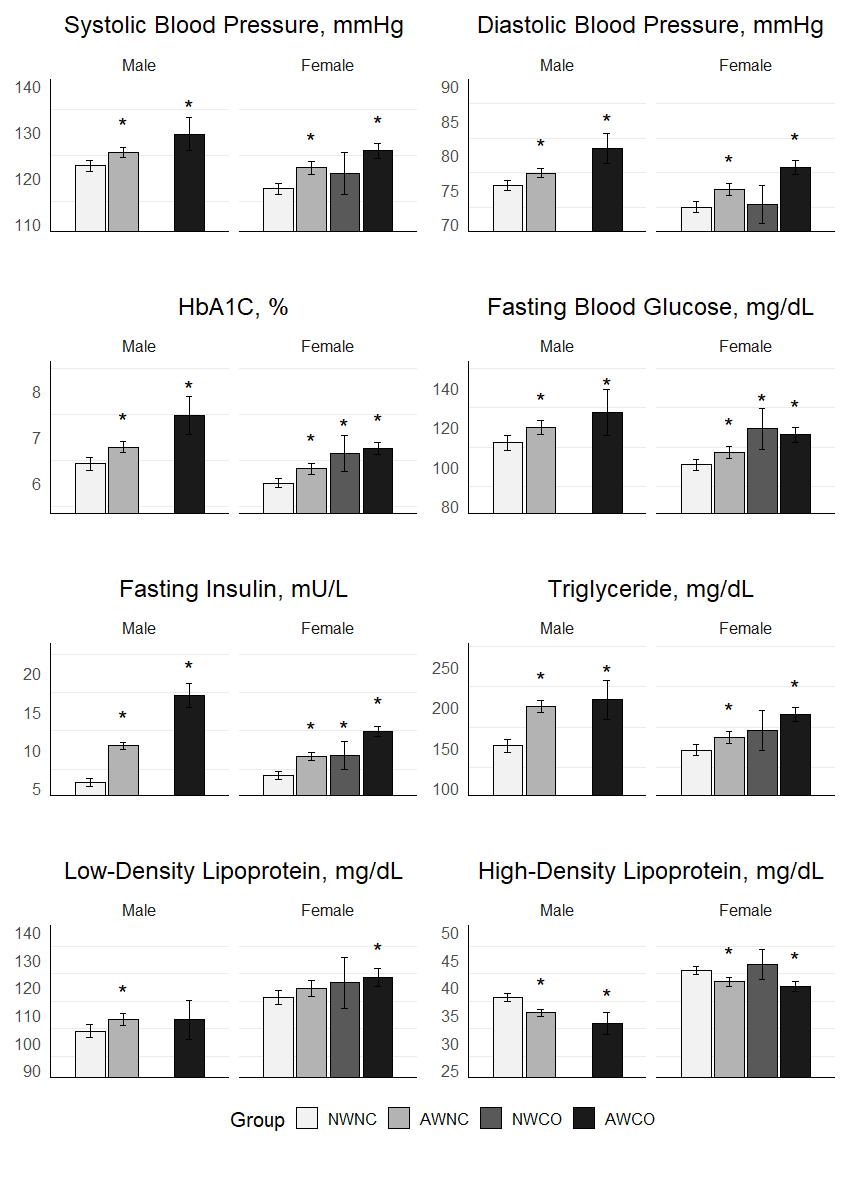

**Supplementary Section 4**

### *SS4. Interaction Effect of age, sex and BMI status on relationship of central obesity markers and cardiometabolic markers.*

#### **SS4.1 Age**

| **Table S12** |  |  |
| --- | --- | --- |
| *TS12. Interaction Effect of Age on the Association of Waist-hip Ratio and Cardiometabolic Markers* | | |
| **Predictor Variables** | **β (95% CI)** | **p-value** |
| **SBP** |  |  |
| Main Effect: WHR | 22.10 (13.00, 31.30) | <0.001 |
| Main Effect:Age 58+ | 20.70 (8.85, 32.50) | 0.001 |
| Interaction: WHR *Age 58+ | **-14.40 (-27.00, -1.74)** | **0.03** |
| **DBP** |  |  |
| Main Effect: WHR | 18.10 (12.50, 23.80) | <0.001 |
| Main Effect:Age 58+ | 11.30 (3.96, 18.50) | 0.003 |
| Interaction: WHR *Age 58+ | **-9.99 (-17.80, -2.19)** | **0.012** |
| **FBG** |  |  |
| Main Effect: WHR | 74.10 (48.20, 100.00) | <0.001 |
| Main Effect:Age 58+ | -22.20 (-55.70, 11.30) | 0.19 |
| Interaction: WHR *Age 58+ | 22.70 (-13.20, 58.50) | 0.22 |
| **HbA1c** |  |  |
| Main Effect: WHR | 3.58 (2.65, 4.50) | <0.001 |
| Main Effect:Age 58+ | -0.56 (-1.75, 0.64) | 0.36 |
| Interaction: WHR *Age 58+ | 0.68 (-0.60, 1.96) | 0.30 |
| **Fasting Insulin** |  |  |
| Main Effect: WHR | 20.20 (16.10, 24.40) | <0.001 |
| Main Effect:Age 58+ | 4.12 (-1.20, 9.43) | 0.1 |
| Interaction: WHR *Age 58+ | -5.65 (-11.30, 0.04) | 0.05 |
| **HOMA-IR** |  |  |
| Main Effect: WHR | 8.11 (6.57, 9.65) | <0.001 |
| Main Effect:Age 58+ | 1.63 (-0.36, 3.63) | 0.11 |
| Interaction: WHR *Age 58+ | -2.15 (-4.28, -0.01) | 0.05 |
| **TC** |  |  |
| Main Effect: WHR | 44.10 (21.20, 67.10) | <0.001 |
| Main Effect:Age 58+ | 60.20 (30.50, 89.90) | <0.001 |
| Interaction: WHR *Age 58+ | **-64.20 (-96.00, -32.40)** | **<0.001** |
| **TG** |  |  |
| Main Effect: WHR | 279.00 (223.00, 336.00) | <0.001 |
| Main Effect:Age 58+ | 89.20 (16.30, 162.00) | 0.017 |
| Interaction: WHR *Age 58+ | **-105.00 (-183.00, -27.40)** | **0.008** |
| **LDL** |  |  |
| Main Effect: WHR | 21.30 (2.49, 40.20) | 0.027 |
| Main Effect:Age 58+ | 41.60 (17.20, 65.90) | 0.001 |
| Interaction: WHR *Age 58+ | **-43.60 (-69.70, -17.50)** | **0.001** |
| **HDL** |  |  |
| Main Effect: WHR | -16.50 (-22.20, -10.80) | <0.001 |
| Main Effect:Age 58+ | 1.15 (-6.20, 8.49) | 0.76 |
| Interaction: WHR *Age 58+ | -1.00 (-8.86, 6.86) | 0.8 |
| **Non-HDL Cholesterol** |  |  |
| Main Effect: WHR | 60.6 (39.3, 82.0) | <0.001 |
| Main Effect: Age ≥58 | 59.1 (31.5, 86.7) | <0.001 |
| Interaction: WHR * Age ≥58 | **-63.2 (-92.7, -33.7)** | **<0.001** |
| **Remnant Cholesterol** |  |  |
| Main Effect: WHR | 39.3 (28.7, 49.9) | <0.001 |
| Main Effect: Age ≥58 | 17.5 (3.75, 31.3) | 0.013 |
| Interaction: WHR * Age ≥58 | **-19.6 (-34.3, -4.86)** | **0.009** |
| *Note. SBP Systolic Blood Pressure, DBP Diastolic Blood Pressure, FBG Fasting Blood Glucose, HbA1c Glycosylated Haemoglobin, HOMA-IR Homeostatic Model Assessment for Insulin Resistance, TC Total Cholesterol, TG Triglyceride, LD Low Density Lipoprotein, HDL High Density Lipoprotein, WHR Wist Hip Ratio* | | |

| **Table S13** |  |  |
| --- | --- | --- |
| *TS13. Interaction Effect of Age on the Association of Waist-Height Ratio and Cardiometabolic Markers* | | |
| **Predictor Variables** | **β (95% CI)** | **p-value** |
| **SBP** |  |  |
| Main Effect: WHtR | 55.2 (42.1, 68.2) | <0.001 |
| Main Effect:Age 58+ | 15.2 (5.38, 25.1) | 0.002 |
| Interaction: WHtR * Age 58+ | -14.9 (-33.3, 3.43) | 0.111 |
| **DBP** |  |  |
| Main Effect: WHtR | 36.8 (28.7, 44.8) | <0.001 |
| Main Effect:Age 58+ | 6.89 (0.813, 13.0) | 0.026 |
| Interaction: WHtR * Age 58+ | -9.21 (-20.6, 2.13) | 0.111 |
| **FBG** |  |  |
| Main Effect: WHtR | 71.3 (33.6, 109) | <0.001 |
| Main Effect:Age 58+ | -20.8 (-49.2, 7.61) | 0.151 |
| Interaction: WHtR * Age 58+ | 38.5 (-14.5, 91.5) | 0.155 |
| **HbA1c** |  |  |
| Main Effect: WHtR | 4.02 (2.67, 5.36) | <0.001 |
| Main Effect:Age 58+ | -0.725 (-1.74, 0.288) | 0.161 |
| Interaction: WHtR * Age 58+ | 1.57 (-0.323, 3.45) | 0.104 |
| **Fasting Insulin** |  |  |
| Main Effect: WHtR | 48.0 (42.3, 53.6) | <0.001 |
| Main Effect:Age 58+ | 2.51 (-1.78, 6.79) | 0.252 |
| Interaction: WHtR * Age 58+ | -6.79 (-14.8, 1.20) | 0.096 |
| **HOMA-IR** |  |  |
| Main Effect: WHtR | 15.8 (13.7, 18.0) | <0.001 |
| Main Effect:Age 58+ | 0.674 (-0.962, 2.31) | 0.419 |
| Interaction: WHtR * Age 58+ | -1.89 (-4.94, 1.16) | 0.224 |
| **TC** |  |  |
| Main Effect: WHtR | 94.4 (61.3, 127) | <0.001 |
| Main Effect:Age 58+ | 7.23 (-17.7, 32.2) | 0.57 |
| Interaction: WHtR * Age 58+ | -13.2 (-59.7, 33.3) | 0.578 |
| **TG** |  |  |
| Main Effect: WHtR | 359 (277, 441) | <0.001 |
| Main Effect:Age 58+ | 16.5 (-45.0, 78.0) | 0.598 |
| Interaction: WHtR * Age 58+ | -44.5 (-159, 70.2) | 0.447 |
| **LDL** |  |  |
| Main Effect: WHtR | 56.6 (29.5, 83.8) | <0.001 |
| Main Effect:Age 58+ | 4.05 (-16.4, 24.5) | 0.698 |
| Interaction: WHtR * Age 58+ | -6.32 (-44.5, 31.9) | 0.746 |
| **HDL** |  |  |
| Main Effect: WHtR | -20.4 (-28.7, -12.1) | <0.001 |
| Main Effect:Age 58+ | -3.24 (-9.47, 2.98) | 0.307 |
| Interaction: WHtR * Age 58+ | 6.18 (-5.43, 17.8) | 0.297 |
| **Non-HDL Cholesterol** |  |  |
| Main Effect: WHtR | 115 (84.2, 145) | <0.001 |
| Main Effect: Age ≥58 | 10.5 (-12.6, 33.5) | 0.373 |
| Interaction: WHtR * Age ≥58 | -19.4 (-62.4, 23.6) | 0.377 |
| **Remnant Cholesterol** |  |  |
| Main Effect: WHtR | 58.1 (42.8, 73.5) | <0.001 |
| Main Effect: Age ≥58 | 6.42 (-5.14, 18.0) | 0.276 |
| Interaction: WHtR * Age ≥58 | -13.1 (-34.6, 8.5) | 0.235 |
| *Note. SBP Systolic Blood Pressure, DBP Diastolic Blood Pressure, FBG Fasting Blood Glucose, HbA1c Glycosylated Haemoglobin, HOMA-IR Homeostatic Model Assessment for Insulin Resistance, TC Total Cholesterol, TG Triglyceride, LD Low Density Lipoprotein, HDL High Density Lipoprotein, WHtR Waist-Height Ratio* | | |

| **Table S14** |  |  |
| --- | --- | --- |
| *TS14. Interaction Effect of Age on the Association of Waist Circumference and Cardiometabolic Markers* | | |
| **Predictor Variables** | **β (95% CI)** | **p-value** |
| **SBP** |  |  |
| Main Effect: WC | 0.40 (0.32, 0.48) | <0.001 |
| Main Effect: Age ≥58 | 23.30 (13.90, 32.70) | <0.001 |
| Interaction: WC * Age ≥58 | **-0.19 (-0.30, -0.08)** | **0.001** |
| **DBP** |  |  |
| Main Effect: WC | 0.27 (0.22, 0.31) | <0.001 |
| Main Effect: Age ≥58 | 9.56 (3.78, 15.34) | 0.001 |
| Interaction: WC * Age ≥58 | **-0.09 (-0.16, -0.02)** | **0.012** |
| **FBG** |  |  |
| Main Effect: WC | 0.69 (0.46, 0.91) | <0.001 |
| Main Effect: Age ≥58 | -15.60 (-42.70, 11.40) | 0.257 |
| Interaction: WC * Age ≥58 | 0.19 (-0.13, 0.51) | 0.25 |
| **HbA1c** |  |  |
| Main Effect: WC | 0.03 (0.02, 0.04) | <0.001 |
| Main Effect: Age ≥58 | -0.54 (-1.50, 0.42) | 0.271 |
| Interaction: WC * Age ≥58 | 0.01 (-0.00, 0.02) | 0.167 |
| **Fasting Insulin** |  |  |
| Main Effect: WC | 0.30 (0.27, 0.34) | <0.001 |
| Main Effect: Age ≥58 | 4.34 (0.24, 8.43) | 0.038 |
| Interaction: WC * Age ≥58 | **-0.06 (-0.11, -0.01)** | **0.012** |
| **HOMA-IR** |  |  |
| Main Effect: WC | 0.11 (0.09, 0.12) | <0.001 |
| Main Effect: Age ≥58 | 1.35 (-0.21, 2.90) | 0.089 |
| Interaction: WC * Age ≥58 | **-0.02 (-0.04, -0.00)** | **0.04** |
| **TC** |  |  |
| Main Effect: WC | 0.33 (0.13, 0.53) | 0.001 |
| Main Effect: Age ≥58 | 16.60 (-7.44, 40.60) | 0.176 |
| Interaction: WC * Age ≥58 | -0.19 (-0.47, 0.09) | 0.19 |
| **TG** |  |  |
| Main Effect: WC | 2.53 (2.04, 3.01) | <0.001 |
| Main Effect: Age ≥58 | 23.70 (-34.80, 82.20) | 0.427 |
| Interaction: WC * Age ≥58 | -0.35 (-1.04, 0.34) | 0.322 |
| **LDL** |  |  |
| Main Effect: WC | 0.15 (-0.02, 0.31) | 0.082 |
| Main Effect: Age ≥58 | 10.60 (-9.10, 30.30) | 0.292 |
| Interaction: WC * Age ≥58 | -0.12 (-0.35, 0.12) | 0.334 |
| **HDL** |  |  |
| Main Effect: WC | -0.22 (-0.27, -0.17) | <0.001 |
| Main Effect: Age ≥58 | -1.91 (-7.78, 3.96) | 0.524 |
| Interaction: WC * Age ≥58 | 0.02 (-0.05, 0.09) | 0.527 |
| **Non-HDL Cholesterol** |  |  |
| Main Effect: WC | 0.55 (0.37, 0.74) | <0.001 |
| Main Effect: Age ≥58 | 18.50 (-3.74, 40.74) | 0.103 |
| Interaction: WC * Age ≥58 | -0.21 (-0.48, 0.05) | 0.113 |
| **Remnant Cholesterol** |  |  |
| Main Effect: WC | 0.41 (0.32, 0.50) | <0.001 |
| Main Effect: Age ≥58 | 7.88 (-3.13, 18.89) | 0.161 |
| Interaction: WC * Age ≥58 | -0.10 (-0.23, 0.03) | 0.143 |
| *Note. SBP Systolic Blood Pressure, DBP Diastolic Blood Pressure, FBG Fasting Blood Glucose, HbA1c Glycosylated Haemoglobin, HOMA-IR Homeostatic Model Assessment for Insulin Resistance, TC Total Cholesterol, TG Triglyceride, LD Low Density Lipoprotein, HDL High Density Lipoprotein, WC Waist Circumference* | | |

**Figure S6**

##### *FS6. Interaction effect of age on the association of waist circumference (WC) and cardiometabolic marker.*

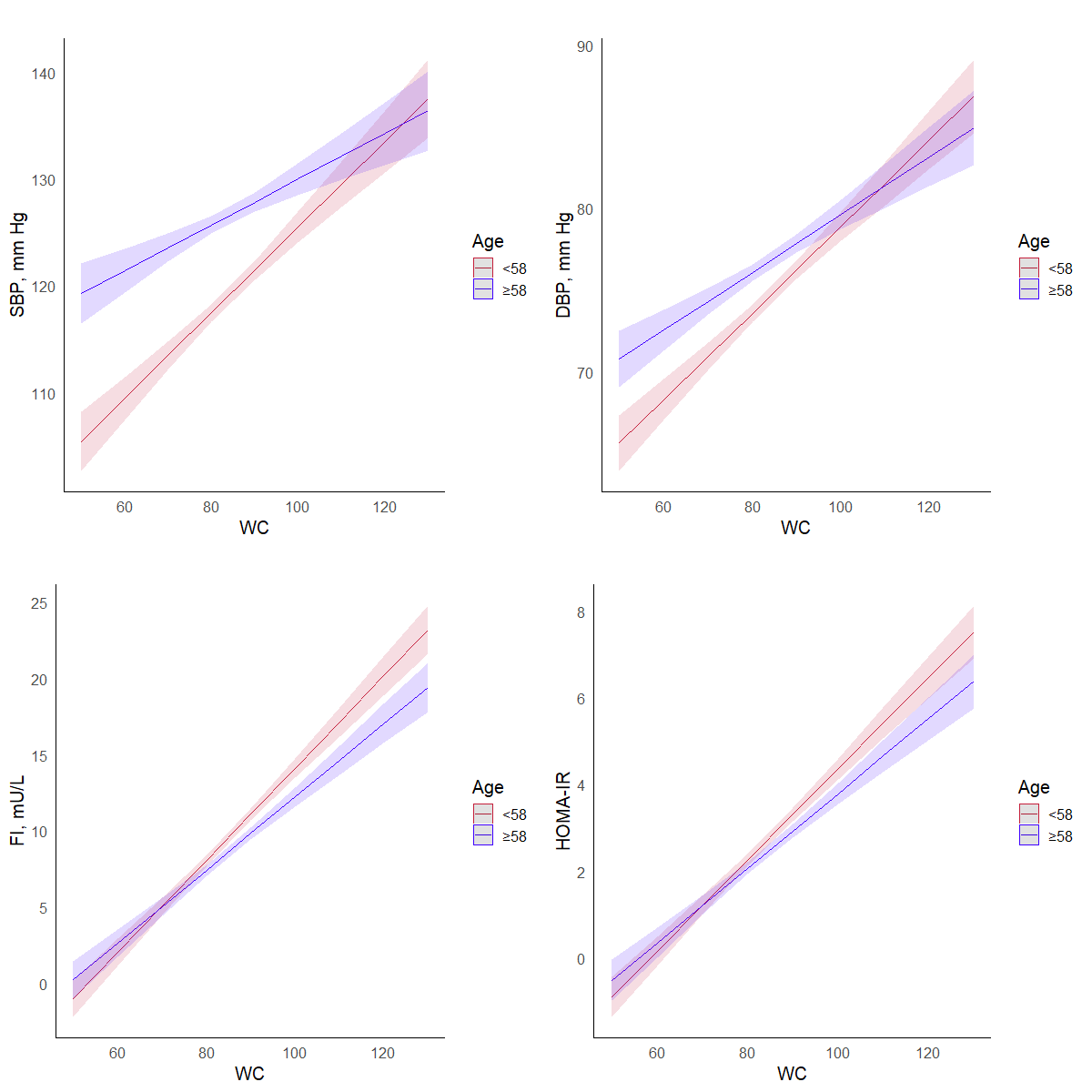

*Note. SBP Systolic Blood Pressure, DBP Diastolic Blood Pressure, FI Fasting Insulin, HOMA-IR Homeostatic Model Assessment for Insulin Resistance, WC Waist Circumference.*

| **Table S15** |  |  |
| --- | --- | --- |
| *TS15. Interaction Effect of Age on the Association of Visceral Fat Percentage and Cardiometabolic Markers* | | |
| **Predictor Variables** | **β (95% CI)** | **p-value** |
| **SBP** |  |  |
| Main Effect: VFP | 1.11 (0.92, 1.31) | <0.001 |
| Main Effect: Age ≥58 | 12.60 (9.77, 15.40) | <0.001 |
| Interaction: VFP * Age ≥58 | **-0.61 (-0.88, -0.35)** | **<0.001** |
| **DBP** |  |  |
| Main Effect: VFP | 0.69 (0.57, 0.81) | <0.001 |
| Main Effect: Age ≥58 | 4.97 (3.24, 6.70) | <0.001 |
| Interaction: VFP * Age ≥58 | **-0.35 (-0.52, -0.19)** | **<0.001** |
| **FBG** |  |  |
| Main Effect: VFP | 1.79 (1.22, 2.36) | <0.001 |
| Main Effect: Age ≥58 | 2.36 (-5.74, 10.46) | 0.568 |
| Interaction: VFP * Age ≥58 | -0.41 (-1.18, 0.36) | 0.295 |
| **HbA1c** |  |  |
| Main Effect: VFP | 0.07 (0.05, 0.10) | <0.001 |
| Main Effect: Age ≥58 | 0.14 (-0.15, 0.43) | 0.343 |
| Interaction: VFP * Age ≥58 | **-0.01 (-0.04, 0.02)** | **0.529** |
| **Fasting Insulin** |  |  |
| Main Effect: VFP | 0.63 (0.54, 0.72) | <0.001 |
| Main Effect: Age ≥58 | 1.25 (-0.01, 2.51) | 0.051 |
| Interaction: VFP * Age ≥58 | **-0.28 (-0.40, -0.16)** | **<0.001** |
| **HOMA-IR** |  |  |
| Main Effect: VFP | 0.23 (0.19, 0.26) | <0.001 |
| Main Effect: Age ≥58 | 0.53 (0.05, 1.00) | 0.029 |
| Interaction: VFP * Age ≥58 | **-0.10 (-0.15, -0.06)** | **<0.001** |
| **TC** |  |  |
| Main Effect: VFP | 0.12 (-0.39, 0.62) | 0.651 |
| Main Effect: Age ≥58 | 12.70 (5.49, 19.80) | <0.001 |
| Interaction: VFP * Age ≥58 | **-1.18 (-1.86, -0.50)** | **<0.001** |
| **TG** |  |  |
| Main Effect: VFP | 6.13 (4.90, 7.37) | <0.001 |
| Main Effect: Age ≥58 | 15.60 (-2.03, 33.10) | 0.083 |
| Interaction: VFP * Age ≥58 | **-2.72 (-4.40, -1.05)** | **0.001** |
| **LDL** |  |  |
| Main Effect: VFP | -0.17 (-0.59, 0.24) | 0.406 |
| Main Effect: Age ≥58 | 8.63 (2.77, 14.50) | 0.004 |
| Interaction: VFP * Age ≥58 | **-0.73 (-1.29, -0.17)** | **0.011** |
| **HDL** |  |  |
| Main Effect: VFP | -0.69 (-0.81, -0.57) | <0.001 |
| Main Effect: Age ≥58 | 0.17 (-1.56, 1.89) | 0.849 |
| Interaction: VFP * Age ≥58 | 0.06 (-0.11, 0.22) | 0.502 |
| **Non-HDL Cholesterol** |  |  |
| Main Effect: VFP | 0.81 (0.34, 1.28) | <0.001 |
| Main Effect: Age ≥58 | 12.50 (5.82, 19.10) | <0.001 |
| Interaction: VFP * Age ≥58 | **-1.24 (-1.87, -0.60)** | **<0.001** |
| **Remnant Cholesterol** |  |  |
| Main Effect: VFP | 0.98 (0.75, 1.21) | <0.001 |
| Main Effect: Age ≥58 | 3.85 (0.55, 7.16) | 0.022 |
| Interaction: VFP * Age ≥58 | **-0.51 (-0.82, -0.20)** | **0.001** |
| *Note. SBP Systolic Blood Pressure, DBP Diastolic Blood Pressure, FBG Fasting Blood Glucose, HbA1c Glycosylated Haemoglobin, HOMA-IR Homeostatic Model Assessment for Insulin Resistance, TC Total Cholesterol, TG Triglyceride, LD Low Density Lipoprotein, HDL High Density Lipoprotein, VFP Visceral Fat Percentage* | | |

**Figure S7**

##### *FS7. Interaction Effect of Age on the Association of Visceral Fat Percentage and Cardiometabolic Markers*

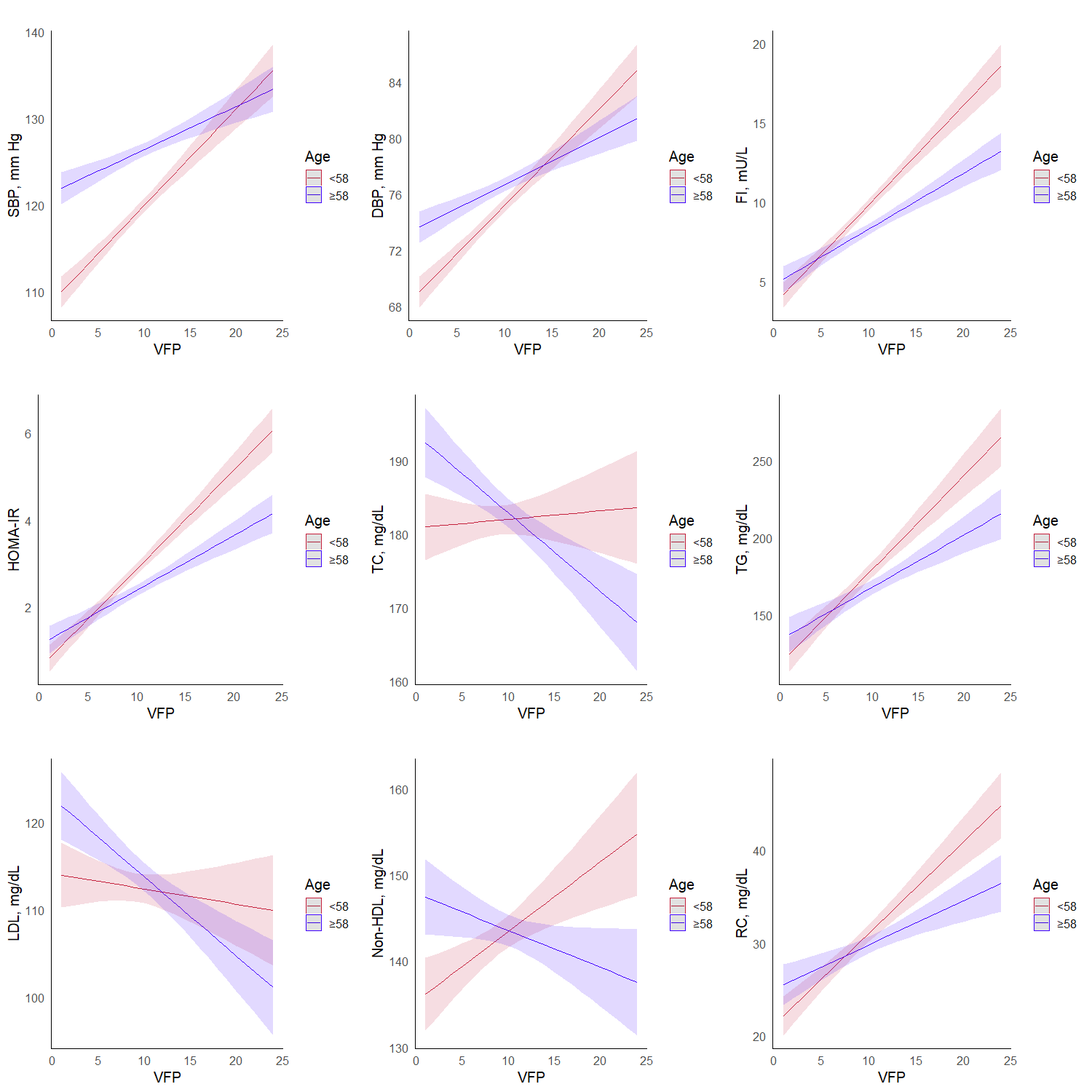

*Note. SBP Systolic Blood Pressure, DBP Diastolic Blood Pressure, FI Fasting Insulin, HOMA-IR Homeostatic Model Assessment for Insulin Resistance, TC Total Cholesterol, TG Triglyceride, LDL Low Density Lipoprotein, VFP Visceral Fat Percentage.*

#### **SS4.2 Sex**

| **Table S16** |  |  |
| --- | --- | --- |
| *TS16. Interaction Effect of Sex on the Association of Waist-Hip Ratio and Cardiometabolic Markers* | | |
| **Predictor Variables** | **β (95% CI)** | **p-value** |
| **SBP** |  |  |
| Main Effect: WHR | 9.50 (0.87, 18.10) | 0.0309 |
| Main Effect: Sex, Male | -6.00 (-20.00, 8.05) | 0.402 |
| Interaction: WHR * Sex, Male | 8.98 (-5.86, 23.80) | 0.236 |
| **DBP** |  |  |
| Main Effect: WHR | 7.81 (2.59, 13.00) | 0.00338 |
| Main Effect: Sex, Male | -6.45 (-15.00, 2.05) | 0.137 |
| Interaction: WHR * Sex, Male | 8.35 (-0.63, 17.30) | 0.0684 |
| **FBG** |  |  |
| Main Effect: WHR | 46.70 (22.90, 70.50) | <0.001 |
| Main Effect: Sex, Male | -73.90 (-113.00, -35.10) | <0.001 |
| Interaction: WHR * Sex, Male | **83.30 (42.30, 124.00)** | **<0.001** |
| **HBA1c** |  |  |
| Main Effect: WHR | 2.56 (1.71, 3.40) | <0.001 |
| Main Effect: Sex, Male | -3.21 (-4.59, -1.83) | <0.001 |
| Interaction: WHR * Sex, Male | **3.51 (2.05, 4.97)** | **<0.001** |
| **Fasting Insulin** |  |  |
| Main Effect: WHR | 13.50 (9.74, 17.30) | <0.001 |
| Main Effect: Sex, Male | -19.70 (-25.90, -13.60) | <0.001 |
| Interaction: WHR * Sex, Male | **19.40 (12.90, 25.90)** | **<0.001** |
| **HOMAIR** |  |  |
| Main Effect: WHR | 5.10 (3.68, 6.52) | <0.001 |
| Main Effect: Sex, Male | -7.12 (-9.43, -4.81) | <0.001 |
| Interaction: WHR * Sex, Male | **7.22 (4.78, 9.66)** | **<0.001** |
| **TC** |  |  |
| Main Effect: WHR | 42.00 (21.30, 62.80) | <0.001 |
| Main Effect: Sex, Male | -54.40 (-88.20, -20.60) | 0.00163 |
| Interaction: WHR * Sex, Male | **37.30 (1.63, 73.10)** | **0.04** |
| **TG** |  |  |
| Main Effect: WHR | 164.00 (112.00, 216.00) | <0.001 |
| Main Effect: Sex, Male | -141.00 (-225.00, -55.90) | 0.00115 |
| Interaction: WHR * Sex, Male | **152.00 (62.00, 241.00)** | **0.001** |
| **LDL** |  |  |
| Main Effect: WHR | 28.50 (11.40, 45.50) | 0.00107 |
| Main Effect: Sex, Male | -28.70 (-56.50, -0.96) | 0.0426 |
| Interaction: WHR * Sex, Male | 14.50 (-14.80, 43.80) | 0.332 |
| **HDL** |  |  |
| Main Effect: WHR | -4.20 (-9.31, 0.91) | 0.107 |
| Main Effect: Sex, Male | -0.51 (-8.83, 7.81) | 0.905 |
| Interaction: WHR * Sex, Male | -4.64 (-13.40, 4.16) | 0.301 |
| **Non-HDL Cholesterol** |  |  |
| Main Effect: WHR | 46.2 (26.8, 65.6) | <0.001 |
| Main Effect: Sex, Male | -53.9 (-85.5, -22.2) | <0.001 |
| Interaction: WHR * Sex, Male | **42.0 (8.55, 75.4)** | **0.014** |
| **Remnant Cholesterol** |  |  |
| Main Effect: WHR | 17.7 (7.94, 27.6) | <0.001 |
| Main Effect: Sex, Male | -25.1 (-41.1, -9.16) | 0.002 |
| Interaction: WHR * Sex, Male | **27.5 (10.6, 44.3)** | **0.001** |
| *Note. SBP Systolic Blood Pressure, DBP Diastolic Blood Pressure, FBG Fasting Blood Glucose, HbA1c Glycosylated Haemoglobin, HOMA-IR Homeostatic Model Assessment for Insulin Resistance, TC Total Cholesterol, TG Triglyceride, LD Low Density Lipoprotein, HDL High Density Lipoprotein, WHR Wasit Hip Ratio* | | |

| **Table S17** |  |  |
| --- | --- | --- |
| *TS17. Interaction Effect of Sex on the Association of Waist-Height Ratio and Cardiometabolic Markers* | | |
| **Predictor Variables** | **β (95% CI)** | **p-value** |
| **SBP** |  |  |
| Main Effect: WHtR | 56.7 (44.7, 68.6) | <0.001 |
| Main Effect: Sex, Male | 9.53 (-0.77, 19.8) | 0.07 |
| Interaction: WHtR * Sex, Male | -10.8 (-30.1, 8.45) | 0.271 |
| **DBP** |  |  |
| Main Effect: WHtR | 34.9 (27.7, 42.1) | <0.001 |
| Main Effect: Sex, Male | 3.30 (-2.92, 9.53) | 0.298 |
| Interaction: WHtR * Sex, Male | -1.61 (-13.3, 10.0) | 0.786 |
| **FBG** |  |  |
| Main Effect: WHtR | 92.1 (58.4, 126) | <0.001 |
| Main Effect: Sex, Male | 3.79 (-25.2, 32.8) | 0.798 |
| Interaction: WHtR * Sex, Male | 12.9 (-41.3, 67.0) | 0.642 |
| **HBA1c** |  |  |
| Main Effect: WHtR | 4.69 (3.49, 5.89) | <0.001 |
| Main Effect: Sex, Male | -0.152 (-1.18, 0.881) | 0.773 |
| Interaction: WHtR * Sex, Male | 1.04 (-0.889, 2.97) | 0.29 |
| **Fasting Insulin** |  |  |
| Main Effect: WHtR | 32.9 (27.8, 37.9) | <0.001 |
| Main Effect: Sex, Male | -15.5 (-19.9, -11.1) | <0.001 |
| Interaction: WHtR * Sex, Male | **29.7 (21.5, 37.9)** | **<0.001** |
| **HOMAIR** |  |  |
| Main Effect: WHtR | 11.5 (9.53, 13.4) | <0.001 |
| Main Effect: Sex, Male | -4.46 (-6.13, -2.79) | <0.001 |
| Interaction: WHtR * Sex, Male | **9.01 (5.88, 12.1)** | **<0.001** |
| **TC** |  |  |
| Main Effect: WHtR | 80.9 (51.6, 110) | <0.001 |
| Main Effect: Sex, Male | -12.0 (-37.1, 13.2) | 0.352 |
| Interaction: WHtR * Sex, Male | -5.17 (-52.2, 41.9) | 0.829 |
| **TG** |  |  |
| Main Effect: WHtR | 290 (217, 363) | <0.001 |
| Main Effect: Sex, Male | -56.6 (-119, 6.17) | 0.077 |
| Interaction: WHtR * Sex, Male | **144 (26.8, 262)** | **0.016** |
| **LDL** |  |  |
| Main Effect: WHtR | 55.7 (31.7, 79.8) | <0.001 |
| Main Effect: Sex, Male | 0.795 (-19.9, 21.5) | 0.94 |
| Interaction: WHtR * Sex, Male | -24.7 (-63.3, 14.0) | 0.211 |
| **HDL** |  |  |
| Main Effect: WHtR | -16.0 (-23.1, -8.82) | <0.001 |
| Main Effect: Sex, Male | 0.872 (-5.28, 7.03) | 0.781 |
| Interaction: WHtR * Sex, Male | **-11.9 (-23.4, -0.39)** | **0.043** |
| **Non-HDL Cholesterol** |  |  |
| Main Effect: WHtR | 96.8 (69.5, 124) | <0.001 |
| Main Effect: Sex, Male | -12.8 (-36.3, 10.7) | 0.284 |
| Interaction: WHtR * Sex, Male | 6.72 (-37.2, 50.6) | 0.764 |
| **Remnant Cholesterol** |  |  |
| Main Effect: WHtR | 41.1 (27.4, 54.8) | <0.001 |
| Main Effect: Sex, Male | -13.6 (-25.4, -1.81) | 0.024 |
| Interaction: WHtR * Sex, Male | **31.4 (9.32, 53.5)** | **0.005** |
| *Note. SBP Systolic Blood Pressure, DBP Diastolic Blood Pressure, FBG Fasting Blood Glucose, HbA1c Glycosylated Haemoglobin, HOMA-IR Homeostatic Model Assessment for Insulin Resistance, TC Total Cholesterol, TG Triglyceride, LD Low Density Lipoprotein, HDL High Density Lipoprotein, WHtR Waist-Height Ratio* | | |

**Figure S8**

##### *FS8. Interaction Effect of Sex on the Association of Waist-Height Ratio and Cardiometabolic Markers*

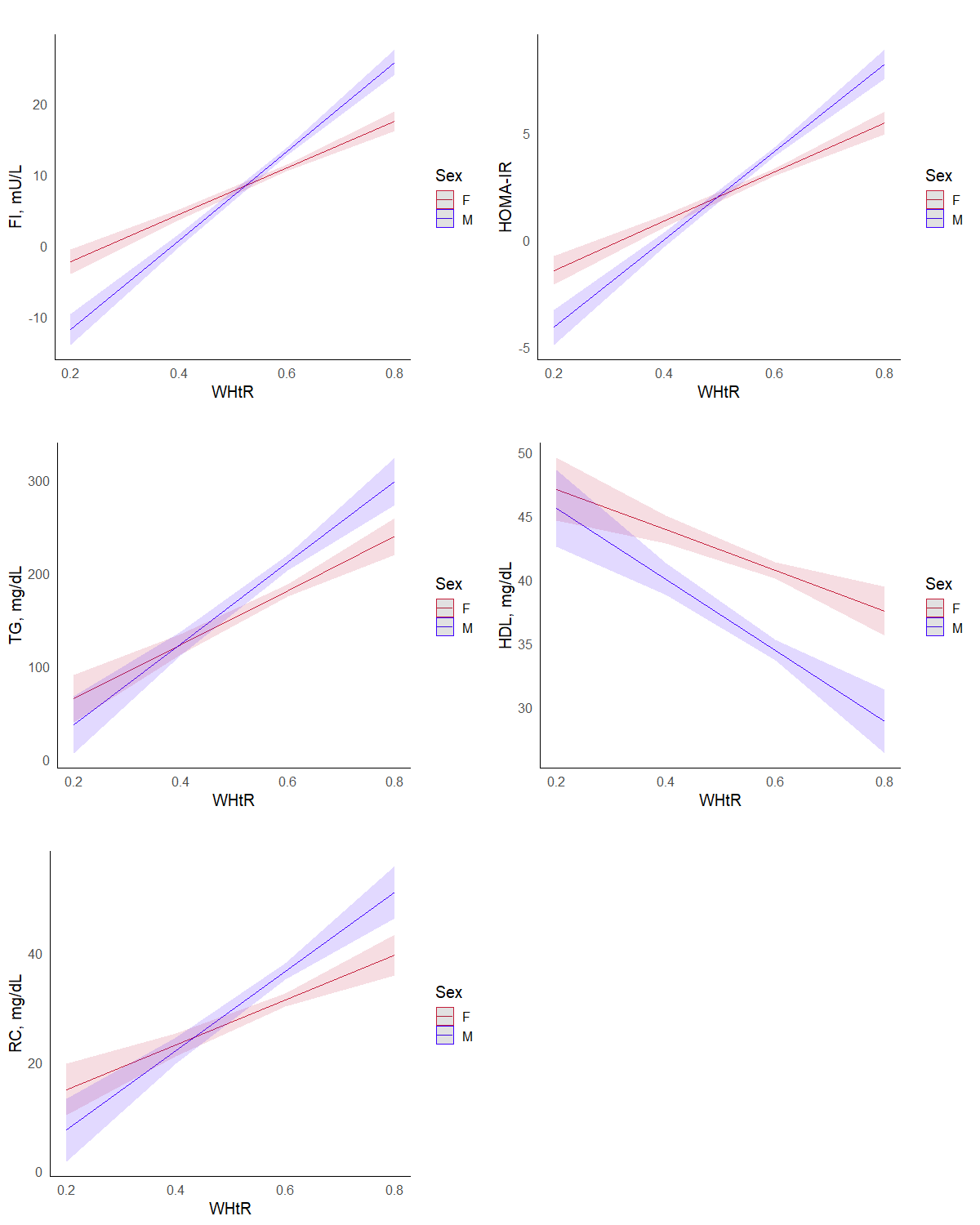

*Note. SBP Systolic Blood Pressure, DBP Diastolic Blood Pressure, FI Fasting Insulin, HOMA-IR Homeostatic Model Assessment for Insulin Resistance, TC Total Cholesterol, TG Triglyceride, LDL Low Density Lipoprotein, WHtR Waist-Height Ratio.*

| **Table S18** |  |  |
| --- | --- | --- |
| *TS18. Interaction Effect of Sex on the Association of Waist Circumference and Cardiometabolic Markers* | | |
| **Predictor Variables** | **β (95% CI)** | **p-value** |
| **SBP** |  |  |
| Main Effect: WC | 0.32 (0.24, 0.40) | <0.001 |
| Main Effect: Sex, Male | 8.08 (-2.14, 18.3) | 0.121 |
| Interaction: WC * Sex, Male | -0.08 (-0.20, 0.05) | 0.221 |
| **DBP** |  |  |
| Main Effect: WC | 0.21 (0.16, 0.26) | <0.001 |
| Main Effect: Sex, Male | 1.85 (-4.31, 8.02) | 0.555 |
| Interaction: WC * Sex, Male | -0.01 (-0.08, 0.06) | 0.781 |
| **FBG** |  |  |
| Main Effect: WC | 0.62 (0.40, 0.84) | <0.001 |
| Main Effect: Sex, Male | -8.34 (-36.9, 20.2) | 0.567 |
| Interaction: WC * Sex, Male | 0.17 (-0.17, 0.51) | 0.325 |
| **HBA1c** |  |  |
| Main Effect: WC | 0.03 (0.02, 0.04) | <0.001 |
| Main Effect: Sex, Male | -0.50 (-1.52, 0.52) | 0.335 |
| Interaction: WC * Sex, Male | 0.008 (-0.004, 0.02) | 0.196 |
| **Fasting Insulin** |  |  |
| Main Effect: WC | 0.22 (0.19, 0.26) | <0.001 |
| Main Effect: Sex, Male | -16.3 (-20.6, -12.0) | <0.001 |
| Interaction: WC * Sex, Male | **0.17 (0.12, 0.22)** | **<0.001** |
| **HOMAIR** |  |  |
| Main Effect: WC | 0.08 (0.06, 0.09) | <0.001 |
| Main Effect: Sex, Male | -5.11 (-6.74, -3.47) | <0.001 |
| Interaction: WC * Sex, Male | **0.06 (0.04, 0.08)** | **<0.001** |
| **TC** |  |  |
| Main Effect: WC | 0.51 (0.31, 0.70) | <0.001 |
| Main Effect: Sex, Male | -18.5 (-43.4, 6.36) | 0.145 |
| Interaction: WC * Sex, Male | 0.003 (-0.29, 0.30) | 0.982 |
| **TG** |  |  |
| Main Effect: WC | 1.91 (1.43, 2.39) | <0.001 |
| Main Effect: Sex, Male | -70.5 (-132, -8.60) | 0.026 |
| Interaction: WC * Sex, Male | **0.89 (0.16, 1.61)** | **0.017** |
| **LDL** |  |  |
| Main Effect: WC | 0.35 (0.19, 0.50) | <0.001 |
| Main Effect: Sex, Male | -5.83 (-26.3, 14.6) | 0.575 |
| Interaction: WC * Sex, Male | -0.10 (-0.34, 0.14) | 0.407 |
| **HDL** |  |  |
| Main Effect: WC | -0.11 (-0.16, -0.06) | <0.001 |
| Main Effect: Sex, Male | 2.39 (-3.68, 8.46) | 0.44 |
| Interaction: WC * Sex, Male | **-0.08 (-0.15, -0.01)** | **0.026** |
| **Non-HDL Cholesterol** |  |  |
| Main Effect: WC | 0.62 (0.44, 0.80) | <0.001 |
| Main Effect: Sex, Male | -20.9 (-44.1, 2.29) | 0.077 |
| Interaction: WC * Sex, Male | 0.08 (-0.19, 0.36) | 0.544 |
| **Remnant Cholesterol** |  |  |
| Main Effect: WC | 0.27 (0.18, 0.36) | <0.001 |
| Main Effect: Sex, Male | -15.1 (-26.7, -3.40) | 0.011 |
| Interaction: WC * Sex, Male | **0.19 (0.05, 0.32)** | **0.008** |
| *Note. SBP Systolic Blood Pressure, DBP Diastolic Blood Pressure, FBG Fasting Blood Glucose, HbA1c Glycosylated Haemoglobin, HOMA-IR Homeostatic Model Assessment for Insulin Resistance, TC Total Cholesterol, TG Triglyceride, LD Low Density Lipoprotein, HDL High Density Lipoprotein, WC Waist Circumference* | | |

**Figure S9**

##### *FS9. Interaction Effect of Sex on the Association of Waist Circumference and Cardiometabolic Markers*

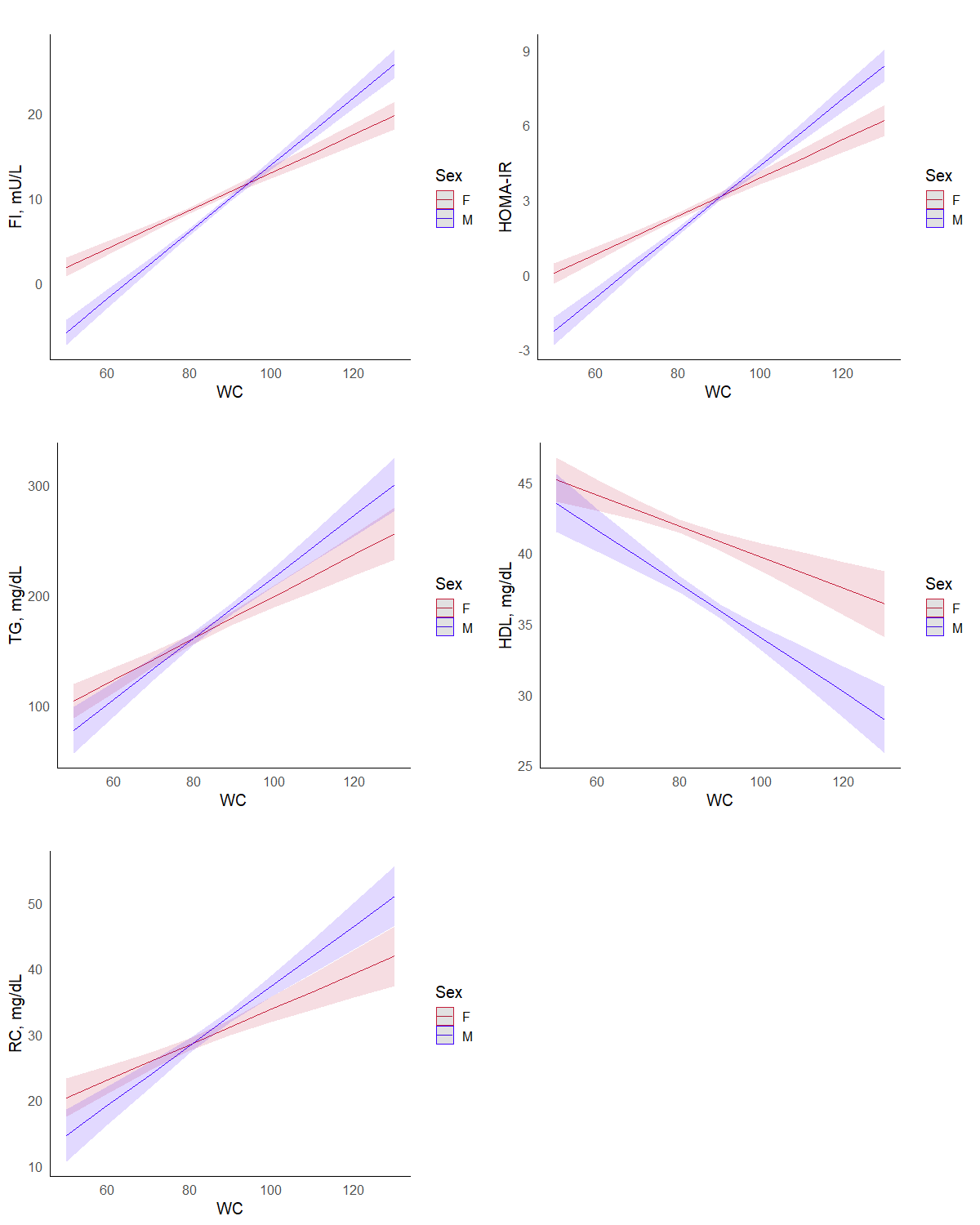

*Note. FI Fasting Insulin, HOMA-IR Homeostatic Model Assessment for Insulin Resistance, TG Triglyceride, HDL High Density Lipoprotein, RC Remnant Cholesterol, WC Waist Circumference*

| **Table S19** |  |  |
| --- | --- | --- |
| *TS19. Interaction Effect of Sex on the Association of Visceral Fat Percentage and Cardiometabolic Markers* | | |
| **Predictor Variables** | **β (95% CI)** | **p-value** |
| **SBP** |  |  |
| Main Effect: VFP | 1.73 (1.42, 2.04) | <0.001 |
| Main Effect: Sex, Male | 6.29 (2.70, 9.89) | 0.0006 |
| Interaction: VFP * Sex, Male | **-0.97 (-1.34, -0.59)** | **<0.001** |
| **DBP** |  |  |
| Main Effect: VFP | 1.03 (0.84, 1.22) | <0.001 |
| Main Effect: Sex, Male | 4.51 (2.33, 6.70) | <0.001 |
| Interaction: VFP * Sex, Male | **-0.63 (-0.85, -0.40)** | **<0.001** |
| **FBG** |  |  |
| Main Effect: VFP | 1.98 (1.10, 2.87) | <0.001 |
| Main Effect: Sex, Male | 11.5 (1.30, 21.7) | 0.027 |
| Interaction: VFP * Sex, Male | -0.97 (-2.04, 0.09) | 0.073 |
| **HBA1c** |  |  |
| Main Effect: VFP | 0.10 (0.07, 0.13) | <0.001 |
| Main Effect: Sex, Male | 0.34 (-0.02, 0.71) | 0.0676 |
| Interaction: VFP * Sex, Male | **-0.04 (-0.08, -0.003)** | **0.035** |
| **Fasting Insulin** |  |  |
| Main Effect: VFP | 0.83 (0.70, 0.97) | <0.001 |
| Main Effect: Sex, Male | -3.36 (-4.93, -1.80) | <0.001 |
| Interaction: VFP * Sex, Male | -0.09 (-0.25, 0.08) | 0.306 |
| **HOMAIR** |  |  |
| Main Effect: VFP | 0.27 (0.22, 0.32) | <0.001 |
| Main Effect: Sex, Male | -0.87 (-1.46, -0.27) | 0.0043 |
| Interaction: VFP * Sex, Male | -0.03 (-0.09, 0.04) | 0.399 |
| **TC** |  |  |
| Main Effect: VFP | 1.43 (0.66, 2.19) | <0.001 |
| Main Effect: Sex, Male | -18.2 (-27.0, -9.31) | <0.001 |
| Interaction: VFP * Sex, Male | -0.38 (-1.30, 0.55) | 0.425 |
| **TG** |  |  |
| Main Effect: VFP | 5.38 (3.45, 7.30) | <0.001 |
| Main Effect: Sex, Male | -9.66 (-31.9, 12.6) | 0.394 |
| Interaction: VFP * Sex, Male | -0.11 (-2.43, 2.21) | 0.929 |
| **LDL** |  |  |
| Main Effect: VFP | 1.03 (0.40, 1.66) | 0.0013 |
| Main Effect: Sex, Male | -13.1 (-20.4, -5.84) | 0.0004 |
| Interaction: VFP * Sex, Male | -0.41 (-1.16, 0.35) | 0.295 |
| **HDL** |  |  |
| Main Effect: VFP | -0.45 (-0.64, -0.26) | <0.001 |
| Main Effect: Sex, Male | -3.74 (-5.90, -1.58) | <0.001 |
| Interaction: VFP * Sex, Male | 0.07 (-0.16, 0.29) | 0.569 |
| **Non-HDL Cholesterol** |  |  |
| Main Effect: VFP | 1.88 (1.16, 2.59) | <0.001 |
| Main Effect: Sex, Male | -14.4 (-22.7, -6.16) | <0.001 |
| Interaction: VFP * Sex, Male | -0.44 (-1.31, 0.42) | 0.316 |
| **Remnant Cholesterol** |  |  |
| Main Effect: VFP | 0.84 (0.48, 1.20) | <0.001 |
| Main Effect: Sex, Male | -1.32 (-5.49, 2.85) | 0.534 |
| Interaction: VFP * Sex, Male | -0.04 (-0.47, 0.40) | 0.868 |
| *Note. SBP Systolic Blood Pressure, DBP Diastolic Blood Pressure, FBG Fasting Blood Glucose, HbA1c Glycosylated Haemoglobin, HOMA-IR Homeostatic Model Assessment for Insulin Resistance, TC Total Cholesterol, TG Triglyceride, LD Low Density Lipoprotein, HDL High Density Lipoprotein, VFP Visceral Fat percentage* | | |

**Figure S10**

##### *FS10. Interaction Effect of Sex on the Association of Visceral Fat Percentage and Cardiometabolic Markers*

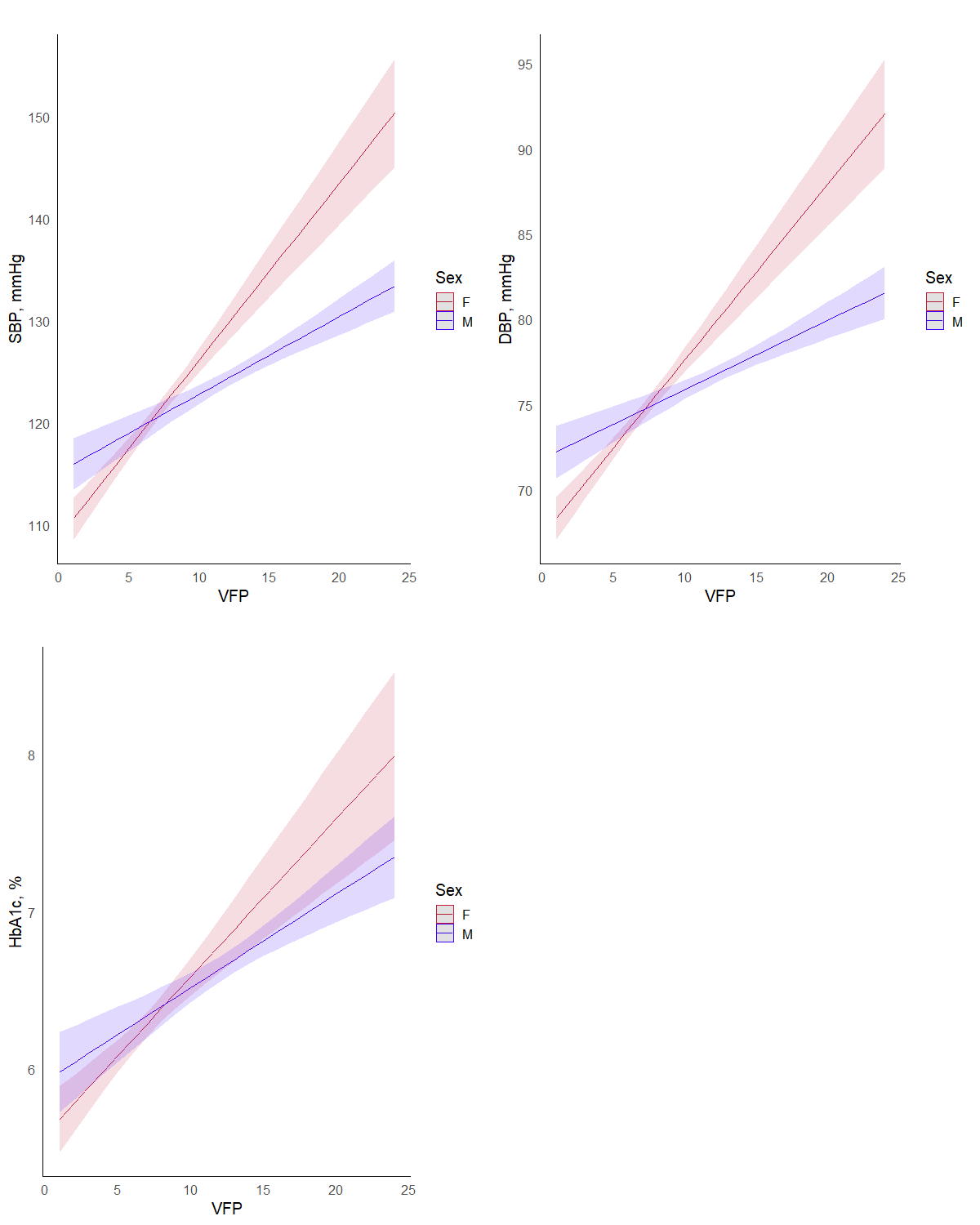

*Note. SBP Systolic Blood Pressure, DBP Diastolic Blood Pressure, HbA1c Glycosylated Haemoglobin, VFp Visceral fat percentage.*

#### **SS4.3 BMI Category**

| **Table S20** |  |  |
| --- | --- | --- |
| *TS20. Interaction Effect of BMI on the Association of Waist-Hip Ratio and Cardiometabolic Markers* | | |
| **Predictor Variables** | **β (95% CI)** | **p-value** |
| **SBP** |  |  |
| Main Effect: WHR | 16.7 (7.28, 26.2) | <0.001 |
| Main Effect: Abnormal BMI | 11.1 (-1.15, 23.4) | 0.08 |
| Interaction: WHR * Abnormal BMI | -7.50 (-20.7, 5.66) | 0.26 |
| **DBP** |  |  |
| Main Effect: WHR | 7.52 (1.80, 13.2) | 0.01 |
| Main Effect: Abnormal BMI | -2.81 (-10.2, 4.60) | 0.46 |
| Interaction: WHR * Abnormal BMI | 5.85 (-2.11, 13.8) | 0.15 |
| **FBG** |  |  |
| Main Effect: WHR | 74.1 (47.8, 100) | <0.001 |
| Main Effect: Abnormal BMI | -1.56 (-35.6, 32.5) | 0.93 |
| Interaction: WHR * Abnormal BMI | 7.81 (-28.8, 44.4) | 0.68 |
| **HBA1c** |  |  |
| Main Effect: WHR | 3.47 (2.54, 4.41) | <0.001 |
| Main Effect: Abnormal BMI | 0.0949 (-1.12, 1.31) | 0.88 |
| Interaction: WHR * Abnormal BMI | 0.220 (-1.08, 1.52) | 0.74 |
| **Fasting Insulin** |  |  |
| Main Effect: WHR | 4.49 (0.455, 8.52) | 0.03 |
| Main Effect: Abnormal BMI | -8.76 (-14.0, -3.54) | 0.00 |
| Interaction: WHR * Abnormal BMI | **13.8 (8.19, 19.4)** | **<0.001** |
| **HOMAIR** |  |  |
| Main Effect: WHR | 2.62 (1.09, 4.15) | 0.00 |
| Main Effect: Abnormal BMI | -3.42 (-5.41, -1.44) | 0.00 |
| Interaction: WHR * Abnormal BMI | **5.04 (2.91, 7.17)** | **<0.001** |
| **TC** |  |  |
| Main Effect: WHR | 5.91 (-17.4, 29.2) | 0.62 |
| Main Effect: Abnormal BMI | 14.5 (-15.8, 44.7) | 0.35 |
| Interaction: WHR * Abnormal BMI | -7.94 (-40.4, 24.5) | 0.63 |
| **TG** |  |  |
| Main Effect: WHR | 132 (75.2, 189) | <0.001 |
| Main Effect: Abnormal BMI | -57.5 (-131, 16.1) | 0.13 |
| Interaction: WHR * Abnormal BMI | **95.0 (16.0, 174)** | **0.02** |
| **LDL** |  |  |
| Main Effect: WHR | 5.19 (-14.0, 24.3) | 0.60 |
| Main Effect: Abnormal BMI | 25.7 (0.894, 50.5) | 0.04 |
| Interaction: WHR * Abnormal BMI | -22.9 (-49.5, 3.73) | 0.09 |
| **HDL** |  |  |
| Main Effect: WHR | -13.3 (-19.0, -7.54) | <0.001 |
| Main Effect: Abnormal BMI | -0.794 (-8.24, 6.65) | 0.83 |
| Interaction: WHR * Abnormal BMI | -1.54 (-9.53, 6.45) | 0.71 |
| **Non-HDL Cholesterol** |  |  |
| Main Effect: WHR | 19.2 (-2.39, 40.8) | 0.08 |
| Main Effect: Abnormal BMI | 15.3 (-12.7, 43.3) | 0.28 |
| Interaction: WHR * Abnormal BMI | -6.40 (-36.4, 23.6) | 0.68 |
| **Remnant Cholesterol** |  |  |
| Main Effect: WHR | 14.0 (3.27, 24.7) | 0.01 |
| Main Effect: Abnormal BMI | -10.4 (-24.3, 3.48) | 0.14 |
| Interaction: WHR * Abnormal BMI | **16.5 (1.58, 31.4)** | **0.03** |
| *Note. SBP Systolic Blood Pressure, DBP Diastolic Blood Pressure, FBG Fasting Blood Glucose, HbA1c Glycosylated Haemoglobin, HOMA-IR Homeostatic Model Assessment for Insulin Resistance, TC Total Cholesterol, TG Triglyceride, LD Low Density Lipoprotein, HDL High Density Lipoprotein, WHR Waist-Hip Ratio* | | |

**Figure S11**

##### *FS11. Interaction Effect of BMI on the Association of Waist-Hip Ratio and Cardiometabolic Markers*

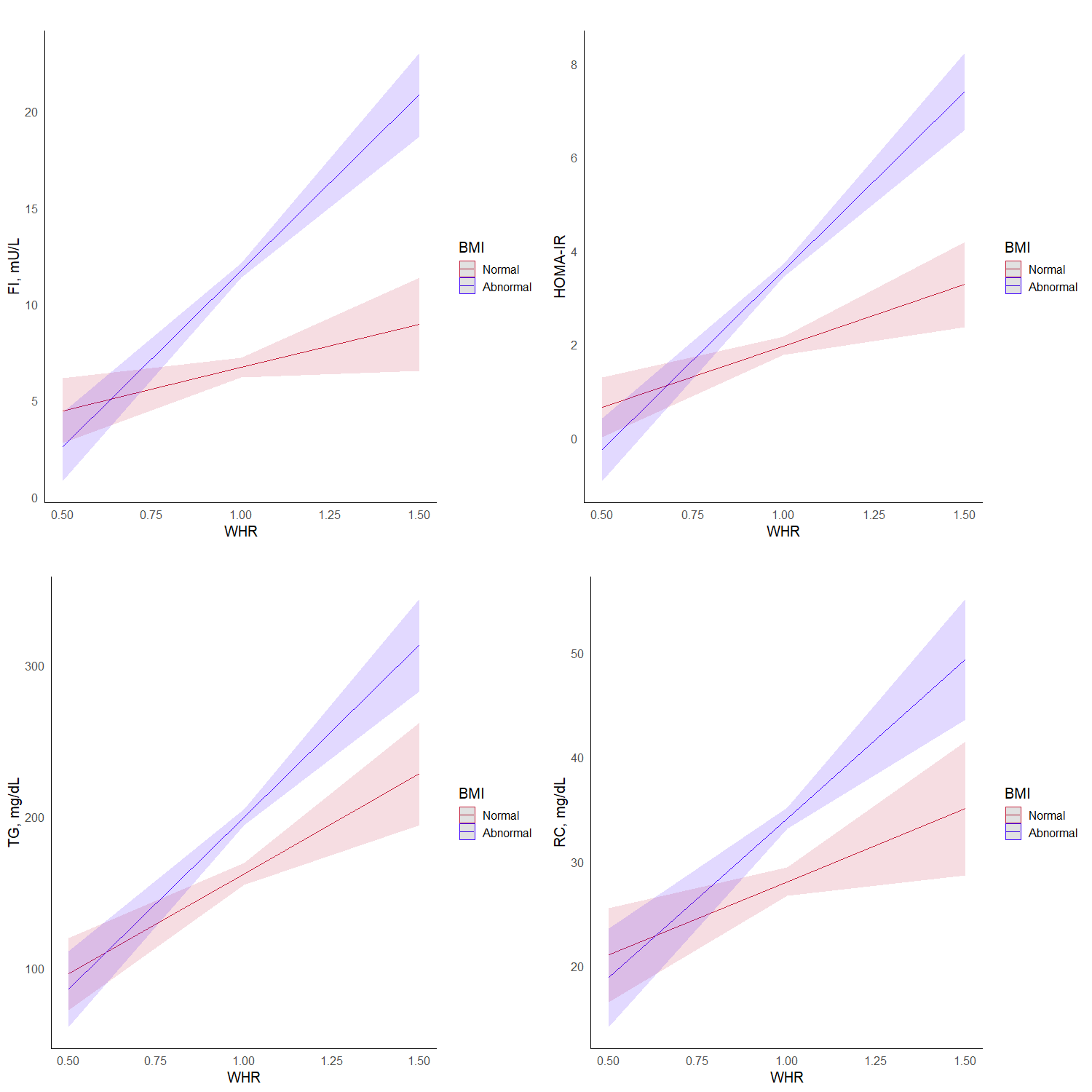

*Note. FI Fasting Insulin, HOMA-IR Homeostatic Model Assessment for Insulin Resistance, TG Triglyceride, RC Remnant Cholesterol, WHR Waist-Hip Ratio*

| **Table S21** |  |  |
| --- | --- | --- |
| *TS21. Interaction Effect of BMI on the Association of Waist-Height Ratio and Cardiometabolic Markers* | | |
| **Predictor Variables** | **β (95% CI)** | **p-value** |
| **SBP** |  |  |
| Main Effect: WHtR | 38.9 (19.6, 58.1) | <0.001 |
| Main Effect: Abnormal BMI | -0.47 (-13.1, 12.1) | 0.94 |
| Interaction: WHtR * Abnormal BMI | 4.28 (-19.9, 28.4) | 0.73 |
| **DBP** |  |  |
| Main Effect: WHtR | 13.3 (1.72, 25.0) | 0.02 |
| Main Effect: Abnormal BMI | -10.3 (-18.0, -2.72) | 0.01 |
| Interaction: WHtR * Abnormal BMI | **22.1 (7.46, 36.7)** | **<0.001** |
| **FBG** |  |  |
| Main Effect: WHtR | 99.7 (45.5, 154) | <0.001 |
| Main Effect: Abnormal BMI | 25.9 (-9.61, 61.4) | 0.15 |
| Interaction: WHtR * Abnormal BMI | -42.8 (-111, 25.2) | 0.22 |
| **HBA1c** |  |  |
| Main Effect: WHtR | 4.84 (2.91, 6.77) | <0.001 |
| Main Effect: Abnormal BMI | 0.76 (-0.50, 2.03) | 0.24 |
| Interaction: WHtR * Abnormal BMI | -1.18 (-3.60, 1.24) | 0.34 |
| **Fasting Insulin** |  |  |
| Main Effect: WHtR | 19.8 (11.7, 28.0) | <0.001 |
| Main Effect: Abnormal BMI | -9.16 (-14.5, -3.84) | 0.00 |
| Interaction: WHtR * Abnormal BMI | **21.8 (11.6, 31.9)** | **<0.001** |
| **HOMAIR** |  |  |
| Main Effect: WHtR | 8.20 (5.10, 11.3) | <0.001 |
| Main Effect: Abnormal BMI | -2.12 (-4.16, -0.086) | 0.04 |
| Interaction: WHtR * Abnormal BMI | **5.33 (1.43, 9.23)** | **0.01** |
| **TC** |  |  |
| Main Effect: WHtR | 106 (58.2, 153) | <0.001 |
| Main Effect: Abnormal BMI | 22.6 (-8.51, 53.7) | 0.15 |
| Interaction: WHtR * Abnormal BMI | -40.5 (-100, 19.1) | 0.18 |
| **TG** |  |  |
| Main Effect: WHtR | 317 (200, 434) | <0.001 |
| Main Effect: Abnormal BMI | 96.3 (19.8, 173) | 0.01 |
| Interaction: WHtR * Abnormal BMI | -144 (-290, 3.12) | 0.06 |
| **LDL** |  |  |
| Main Effect: WHtR | 71.8 (32.8, 111) | <0.001 |
| Main Effect: Abnormal BMI | 18.2 (-7.41, 43.8) | 0.16 |
| Interaction: WHtR * Abnormal BMI | -33.8 (-82.8, 15.3) | 0.18 |
| **HDL** |  |  |
| Main Effect: WHtR | -10.2 (-22.0, 1.59) | 0.09 |
| Main Effect: Abnormal BMI | -6.31 (-14.0, 1.43) | 0.11 |
| Interaction: WHtR * Abnormal BMI | 7.54 (-7.29, 22.4) | 0.32 |
| **Non-HDL Cholesterol** |  |  |
| Main Effect: WHtR | 116 (72.0, 160) | <0.001 |
| Main Effect: Abnormal BMI | 28.9 (0.156, 57.7) | 0.05 |
| Interaction: WHtR * Abnormal BMI | -48.1 (-103, 7.03) | 0.09 |
| **Remnant Cholesterol** |  |  |
| Main Effect: WHtR | 44.1 (22.1, 66.1) | <0.001 |
| Main Effect: Abnormal BMI | 10.7 (-3.66, 25.1) | 0.14 |
| Interaction: WHtR * Abnormal BMI | -14.3 (-41.9, 13.3) | 0.31 |
| *Note. SBP Systolic Blood Pressure, DBP Diastolic Blood Pressure, FBG Fasting Blood Glucose, HbA1c Glycosylated Haemoglobin, HOMA-IR Homeostatic Model Assessment for Insulin Resistance, TC Total Cholesterol, TG Triglyceride, LD Low Density Lipoprotein, HDL High Density Lipoprotein, WHtR Waist-Height Ratio* | | |

**Figure S12**

##### *FS12. Interaction Effect of BMI on the Association of Waist-Height Ratio and Cardiometabolic Markers*

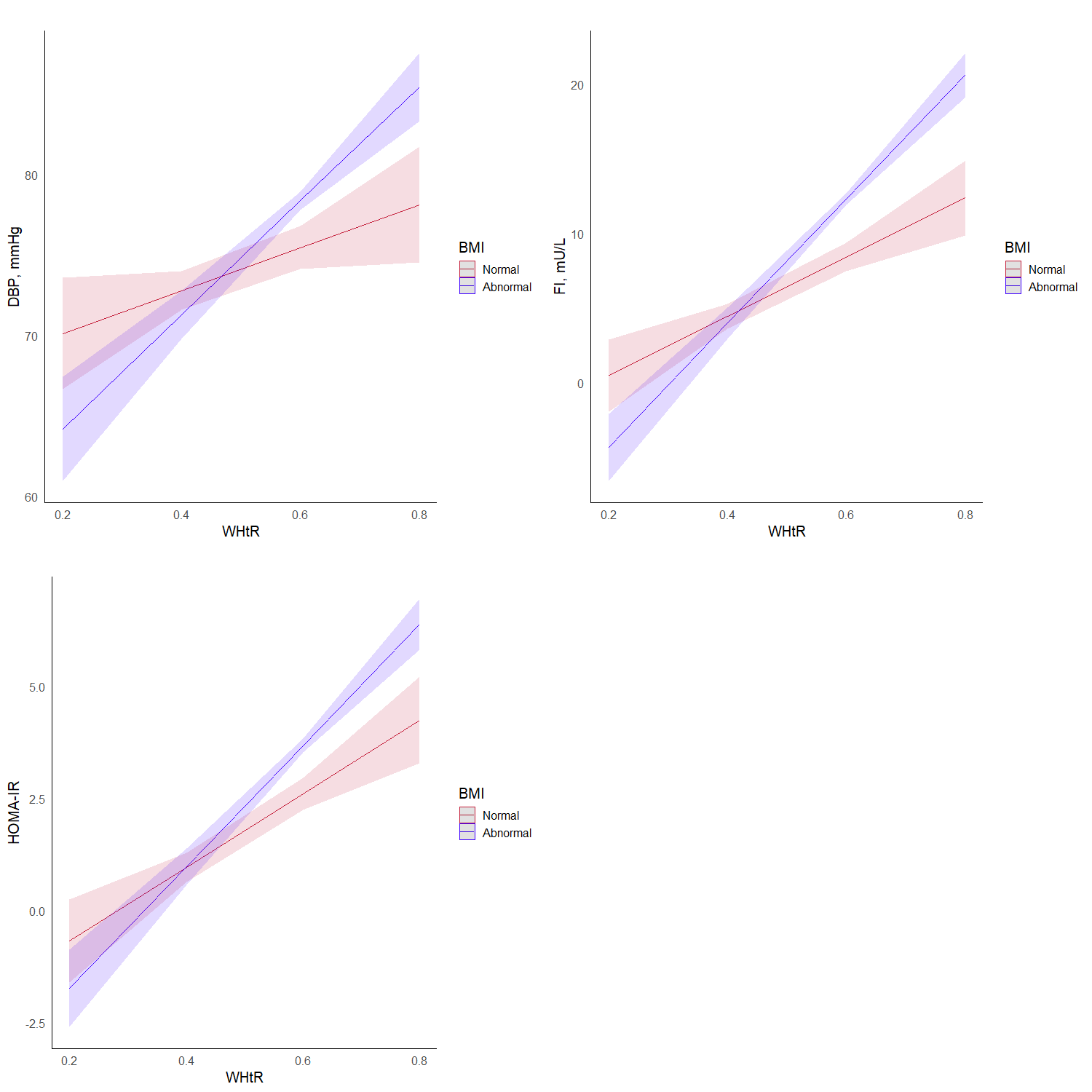

*Note. DBP Diastolic Blood Pressure, FI Fasting insulin, HOMA-IR Homeostatic Model Assessment for Insulin Resistance, WHtR Waist-Height Ratio*

| **Table S22** |  |  |
| --- | --- | --- |
| TS22. Interaction Effect of BMI on the Association of Waist Circumference and Cardiometabolic Markers | | |
| **Predictor Variables** | **β (95% CI)** | **p-value** |
| **SBP** |  |  |
| Main Effect: WC | 0.26 (0.14, 0.37) | <0.001 |
| Main Effect: Abnormal BMI | 1.18 (-10.80, 13.10) | 0.847 |
| Interaction: WC * Abnormal BMI | 0.01 (-0.14, 0.15) | 0.93 |
| **DBP** |  |  |
| Main Effect: WC | 0.14 (0.07, 0.21) | <0.001 |
| Main Effect: Abnormal BMI | -7.47 (-14.70, -0.27) | 0.042 |
| Interaction: WC * Abnormal BMI | **0.10 (0.01, 0.19)** | **0.024** |
| **FBG** |  |  |
| Main Effect: WC | 0.90 (0.57, 1.22) | <0.001 |
| Main Effect: Abnormal BMI | 14.70 (-18.80, 48.20) | 0.389 |
| Interaction: WC * Abnormal BMI | -0.18 (-0.59, 0.23) | 0.382 |
| **HBA1c** |  |  |
| Main Effect: WC | 0.04 (0.03, 0.05) | <0.001 |
| Main Effect: Abnormal BMI | 0.33 (-0.86, 1.52) | 0.588 |
| Interaction: WC * Abnormal BMI | -0.00 (-0.02, 0.01) | 0.623 |
| **Fasting Insulin** |  |  |
| Main Effect: WC | 0.08 (0.03, 0.13) | <0.001 |
| Main Effect: Abnormal BMI | -14.10 (-19.10, -9.05) | <0.001 |
| Interaction: WC * Abnormal BMI | **0.20 (0.14, 0.26)** | **<0.001** |
| **HOMAIR** |  |  |
| Main Effect: WC | 0.04 (0.02, 0.06) | <0.001 |
| Main Effect: Abnormal BMI | -4.40 (-6.31, -2.48) | <0.001 |
| Interaction: WC * Abnormal BMI | **0.06 (0.04, 0.08)** | **<0.001** |
| **TC** |  |  |
| Main Effect: WC | 0.12 (-0.17, 0.41) | 0.404 |
| Main Effect: Abnormal BMI | 17.80 (-11.90, 47.50) | 0.239 |
| Interaction: WC * Abnormal BMI | -0.14 (-0.50, 0.23) | 0.463 |
| **TG** |  |  |
| Main Effect: WC | 1.90 (1.20, 2.60) | <0.001 |
| Main Effect: Abnormal BMI | 20.40 (-52.00, 92.70) | 0.581 |
| Interaction: WC * Abnormal BMI | -0.05 (-0.93, 0.83) | 0.915 |
| **LDL** |  |  |
| Main Effect: WC | 0.08 (-0.16, 0.31) | 0.514 |
| Main Effect: Abnormal BMI | 21.80 (-2.55, 46.20) | 0.079 |
| Interaction: WC * Abnormal BMI | -0.21 (-0.50, 0.09) | 0.169 |
| **HDL** |  |  |
| Main Effect: WC | -0.22 (-0.29, -0.15) | <0.001 |
| Main Effect: Abnormal BMI | -3.28 (-10.50, 3.99) | 0.376 |
| Interaction: WC * Abnormal BMI | 0.03 (-0.06, 0.12) | 0.46 |
| **Non-HDL Cholesterol** |  |  |
| Main Effect: WC | 0.34 (0.07, 0.60) | 0.013 |
| Main Effect: Abnormal BMI | 21.10 (-6.34, 48.50) | 0.132 |
| Interaction: WC * Abnormal BMI | -0.17 (-0.50, 0.17) | 0.322 |
| **Remnant Cholesterol** |  |  |
| Main Effect: WC | 0.26 (0.13, 0.39) | <0.001 |
| Main Effect: Abnormal BMI | -0.70 (-14.30, 12.90) | 0.92 |
| Interaction: WC * Abnormal BMI | 0.04 (-0.13, 0.21) | 0.641 |
| *Note. SBP Systolic Blood Pressure, DBP Diastolic Blood Pressure, FBG Fasting Blood Glucose, HbA1c Glycosylated Haemoglobin, HOMA-IR Homeostatic Model Assessment for Insulin Resistance, TC Total Cholesterol, TG Triglyceride, LD Low Density Lipoprotein, HDL High Density Lipoprotein, WC Waist Circumference* | | |

**Figure S13**

##### *FS13. Interaction Effect of BMI on the Association of Waist Circumference and Cardiometabolic Markers*

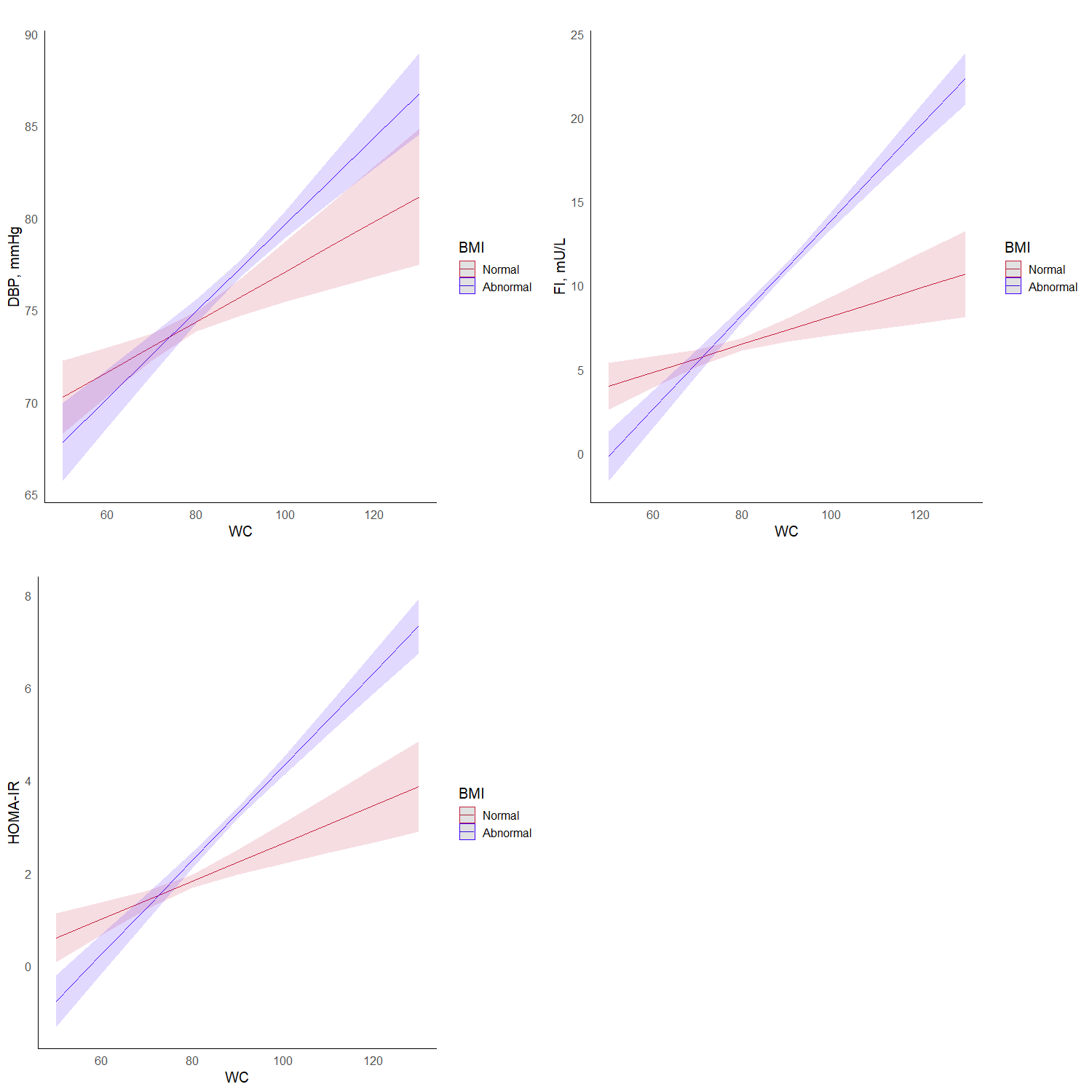

*Note. DBP Diastolic Blood Pressure, FI Fasting insulin, HOMA-IR Homeostatic Model Assessment for Insulin Resistance, WC Waist Circumference*

| **Table S23** |  |  |
| --- | --- | --- |
| *TS23. Interaction Effect of BMI on the Association of Visceral Fat Percentage and Cardiometabolic Markers* | | |
| **Predictor Variables** | **β (95% CI)** | **p-value** |
| **SBP** |  |  |
| Main Effect: VFP | 1.03 (0.77, 1.28) | <0.001 |
| Main Effect: Abnormal BMI | 4.50 (1.39, 7.62) | 0.005 |
| Interaction: VFP * Abnormal BMI | **-0.35 (-0.67, -0.03)** | **0.034** |
| **DBP** |  |  |
| Main Effect: VFP | 0.51 (0.36, 0.67) | <0.001 |
| Main Effect: Abnormal BMI | 2.03 (0.13, 3.92) | 0.036 |
| Interaction: VFP * Abnormal BMI | -0.09 (-0.29, 0.10) | 0.351 |
| **FBG** |  |  |
| Main Effect: VFP | 1.54 (0.82, 2.27) | <0.001 |
| Main Effect: Abnormal BMI | 6.14 (-2.69, 15.00) | 0.173 |
| Interaction: VFP * Abnormal BMI | -0.31 (-1.22, 0.60) | 0.501 |
| **HBA1c** |  |  |
| Main Effect: VFP | 0.07 (0.04, 0.09) | <0.001 |
| Main Effect: Abnormal BMI | 0.31 (-0.005, 0.63) | 0.054 |
| Interaction: VFP * Abnormal BMI | -0.01 (-0.04, 0.02) | 0.466 |
| **Fasting Insulin** |  |  |
| Main Effect: VFP | 0.06 (-0.05, 0.17) | 0.319 |
| Main Effect: Abnormal BMI | 0.79 (-0.56, 2.14) | 0.25 |
| Interaction: VFP * Abnormal BMI | **0.30 (0.16, 0.44)** | **<0.001** |
| **HOMAIR** |  |  |
| Main Effect: VFP | 0.04 (-0.00, 0.08) | 0.062 |
| Main Effect: Abnormal BMI | 0.16 (-0.36, 0.67) | 0.544 |
| Interaction: VFP * Abnormal BMI | **0.10 (0.05, 0.15)** | **<0.001** |
| **TC** |  |  |
| Main Effect: VFP | -1.04 (-1.67, -0.40) | 0.001 |
| Main Effect: Abnormal BMI | 14.50 (6.70, 22.20) | <0.001 |
| Interaction: VFP * Abnormal BMI | -0.27 (-1.07, 0.53) | 0.506 |
| **TG** |  |  |
| Main Effect: VFP | 2.64 (1.07, 4.21) | <0.001 |
| Main Effect: Abnormal BMI | 21.30 (2.14, 40.40) | 0.029 |
| Interaction: VFP * Abnormal BMI | 0.51 (-1.45, 2.47) | 0.609 |
| **LDL** |  |  |
| Main Effect: VFP | -0.77 (-1.29, -0.24) | 0.004 |
| Main Effect: Abnormal BMI | 12.00 (5.68, 18.40) | <0.001 |
| Interaction: VFP * Abnormal BMI | -0.42 (-1.07, 0.24) | 0.212 |
| **HDL** |  |  |
| Main Effect: VFP | -0.64 (-0.79, -0.49) | <0.001 |
| Main Effect: Abnormal BMI | -0.23 (-2.11, 1.65) | 0.809 |
| Interaction: VFP * Abnormal BMI | 0.00 (-0.19, 0.20) | 0.984 |
| **Non-HDL Cholesterol** |  |  |
| Main Effect: VFP | -0.40 (-0.99, 0.19) | 0.188 |
| Main Effect: Abnormal BMI | 14.70 (7.48, 21.90) | <0.001 |
| Interaction: VFP * Abnormal BMI | -0.27 (-1.01, 0.47) | 0.471 |
| **Remnant Cholesterol** |  |  |
| Main Effect: VFP | 0.37 (0.07, 0.66) | 0.015 |
| Main Effect: Abnormal BMI | 2.65 (-0.94, 6.24) | 0.148 |
| Interaction: VFP * Abnormal BMI | 0.14 (-0.23, 0.51) | 0.446 |
| *Note. SBP Systolic Blood Pressure, DBP Diastolic Blood Pressure, FBG Fasting Blood Glucose, HbA1c Glycosylated Haemoglobin, HOMA-IR Homeostatic Model Assessment for Insulin Resistance, TC Total Cholesterol, TG Triglyceride, LD Low Density Lipoprotein, HDL High Density Lipoprotein, VFP Visceral Fat percentage* | | |

**Figure S14**

##### *FS14. Interaction Effect of BMI on the Association of Visceral Fat Percentage and Cardiometabolic Markers*

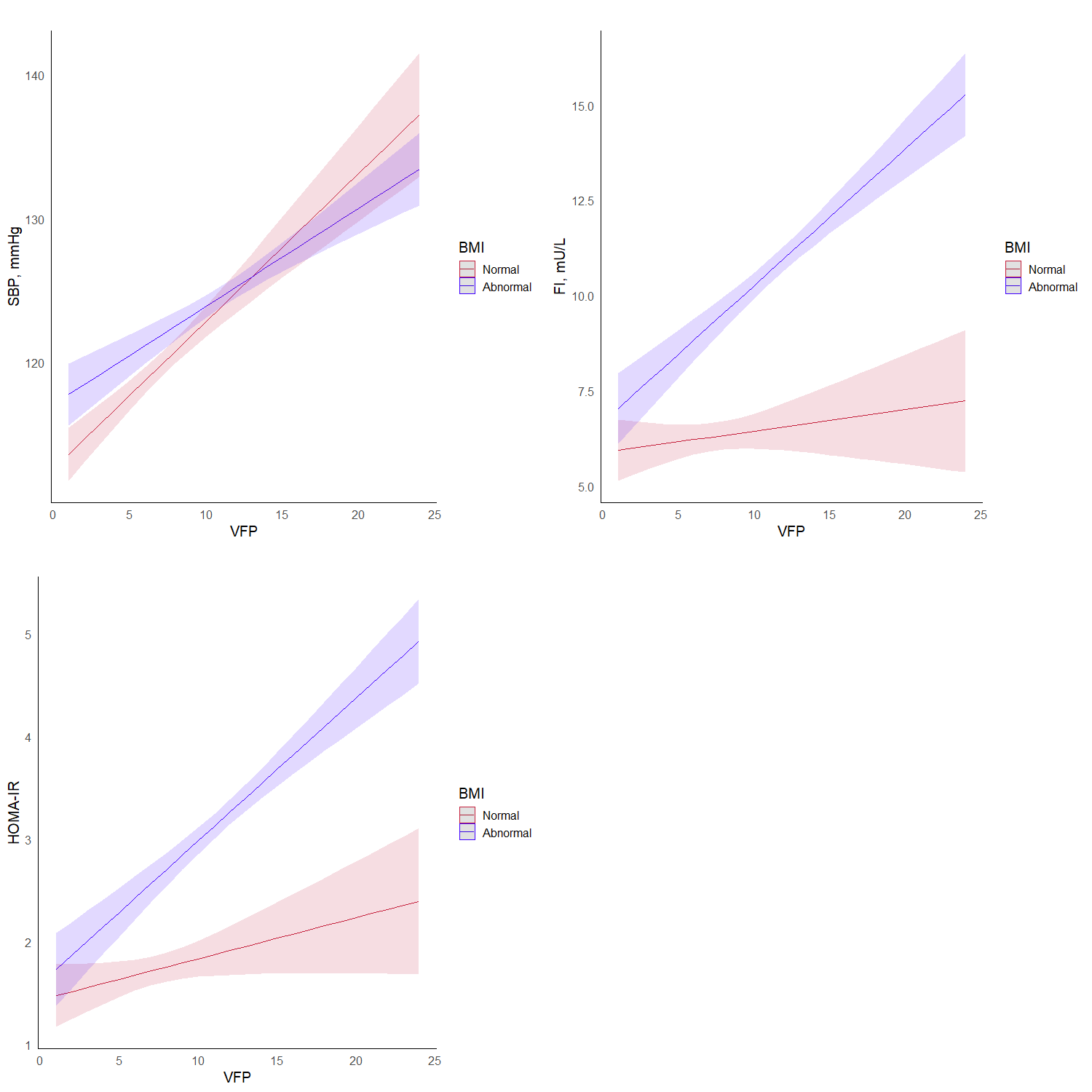

*Note. SBP Systolic Blood Pressure, FI Fasting insulin, HOMA-IR Homeostatic Model Assessment for Insulin Resistance, VFP Visceral Fat Percentage*

**Supplementary Section 5**

### *SS5. Mediation of the relationship of BMI with Cardiometabolic markers via central obesity indices*

| **Table S24** |  |  |  |
| --- | --- | --- | --- |
| *TS24. Adjusted direct and indirect associations of BMI with Cardiometabolic Markers via Waist-Hip Ratio* | | |  |
| **Association** | **β (95% CI)** | **P Value** | **ρ^a^** |
| **SBP** |  |  |  |
| Total association | 1.02 (0.88, 1.18) | <0.001 |  |
| Direct association | 1.03 (0.88, 1.19) | <0.001 |  |
| Indirect association | -0.01 (-0.05, 0.03) | 0.66 |  |
| Proportion mediated, % | - |  |  |
| **DBP** |  |  |  |
| Total association | 0.61 (0.52, 0.71) | <0.001 |  |
| Direct association | 0.59 (0.49, 0.69) | <0.001 | -0.6 |
| Indirect association | 0.03 (0.003, 0.05) | 0.02 | < 0.1 |
| Proportion mediated, % | **4.33 (0.64, 9.00)** |  |  |
| **FBG** |  |  |  |
| Total association | 1.08 (0.69, 1.49) | <0.001 |  |
| Direct association | 0.63 (0.20, 1.10) | 0.006 | -0.2 |
| Indirect association | 0.45 (0.31, 0.59) | <0.001 | 0.1 |
| Proportion mediated, % | **41.40 (25.30, 73.00)** |  |  |
| **HbA1C** |  |  |  |
| Total association | 0.06 (0.05, 0.08) | <0.001 |  |
| Direct association | 0.04 (0.03, 0.06) | <0.001 | -0.3 |
| Indirect association | 0.02 (0.01, 0.02) | <0.001 | 0.2 |
| Proportion mediated, % | **30.71 (21.30, 44.00)** |  |  |
| **Fasting Insulin** |  |  |  |
| Total association | 0.85 (0.76, 0.93) | <0.001 |  |
| Direct association | 0.79 (0.70, 0.87) | <0.001 | -0.8 |
| Indirect association | 0.06 (0.04, 0.08) | <0.001 | 0.1 |
| Proportion mediated, % | **7.03 (4.87, 10.00)** |  |  |
| **HOMA-IR** |  |  |  |
| Total association | 0.26 (0.24, 0.29) | <0.001 |  |
| Direct association | 0.24 (0.21, 0.26) | <0.001 | -0.8 |
| Indirect association | 0.03 (0.02, 0.04) | <0.001 | 0.1 |
| Proportion mediated, % | **10.39 (7.65, 14.00)** |  |  |
| **Total Cholesterol** |  |  |  |
| Total association | 1.50 (1.09, 1.91) | <0.001 |  |
| Direct association | 1.50 (1.10, 1.94) | <0.001 |  |
| Indirect association | -0.00 (-0.11, 0.10) | 0.89 |  |
| Proportion mediated, % | - |  |  |
| **Triglycerides** |  |  |  |
| Total association | 5.66 (4.82, 6.57) | <0.001 |  |
| Direct association | 4.61 (3.67, 5.57) | <0.001 | -0.5 |
| Indirect association | 1.06 (0.75, 1.36) | <0.001 | 0.1 |
| Proportion mediated, % | **18.70 (12.90, 25.00)** |  |  |
| **LDL Cholesterol** |  |  |  |
| Total association | 1.04 (0.71, 1.36) | <0.001 |  |
| Direct association | 1.09 (0.75, 1.43) | <0.001 |  |
| Indirect association | -0.06 (-0.13, 0.02) | 0.17 |  |
| Proportion mediated, % | - |  |  |
| **HDL Cholesterol** |  |  |  |
| Total association | -0.42 (-0.52, -0.32) | <0.001 |  |
| Direct association | -0.33 (-0.44, -0.23) | <0.001 | 0.4 |
| Indirect association | -0.08 (-0.11, -0.05) | <0.001 | -0.1 |
| Proportion mediated, % | **19.72 (12.37, 29.00)** |  |  |
| **Non-HDL Cholesterol** |  |  |  |
| Total association | 1.92 (1.54, 2,29) | <0.001 |  |
| Direct association | 1.84 (1.46, 2.23) | <0.001 |  |
| Indirect association | 0.08 (-0.02, 0.17) | 0.11 |  |
| Proportion mediated, % | - |  |  |
| **Remnant Cholesterol** |  |  |  |
| Total association | 0.88 (0.71, 1.05) | <0.001 |  |
| Direct association | 0.75 (0.56, 0.93) | <0.001 | -0.5 |
| Indirect association | 0.13 (0.08, 0.19) | <0.001 | 0.1 |
| Proportion mediated, % | **15.15 (9.37, 23.00)** |  |  |
| *Note. Adjusted for age, sex, education, income, smoking status, alcohol status.* | | | |
| *^a^ - Rho is the effect size of unknown confounder at which direct or indirect association becomes 0* | | | |
| *Note. SBP Systolic Blood Pressure, DBP Diastolic Blood Pressure, FBG Fasting Blood Glucose, HbA1c Glycosylated Haemoglobin, HOMA-IR Homeostatic Model Assessment for Insulin Resistance, LDL Low Density Lipoprotein, HDL High Density Lipoprotein* | | | |

| **Table S25** |  |  |  |
| --- | --- | --- | --- |
| *TS25. Adjusted direct and indirect associations of BMI with Cardiometabolic Markers via Waist-Height Ratio* | | |  |
| **Association** | **β (95% CI)** | **P Value** | **ρ^a^** |
| **SBP** |  |  |  |
| Total association | 1.02 (0.88, 1.18) | <0.001 |  |
| Direct association | 1.08 (0.81, 1.33) | <0.001 |  |
| Indirect association | -0.06 (-0.25, 0.13) | 0.51 |  |
| Proportion mediated, % | - |  |  |
| **DBP** |  |  |  |
| Total association | 0.61 (0.52, 0.71) | <0.001 |  |
| Direct association | 0.51 (0.36, 0.67) | <0.001 |  |
| Indirect association | 0.10 (-0.02, 0.22) | 0.078 |  |
| Proportion mediated, % | - |  |  |
| **FBG** |  |  |  |
| Total association | 1.08 (0.69, 1.49) | <0.001 |  |
| Direct association | 0.63 (0.20, 1.10) | 0.006 | <0.1 |
| Indirect association | 0.45 (0.31, 0.59) | <0.001 | 0.1 |
| Proportion mediated, % | **41.40 (25.30, 73.00)** |  |  |
| **HbA1C** |  |  |  |
| Total association | 0.06 (0.05, 0.08) | <0.001 |  |
| Direct association | 0.04 (0.03, 0.06) | <0.001 | <0.1 |
| Indirect association | 0.02 (0.01, 0.02) | <0.001 | 0.1 |
| Proportion mediated, % | **30.71 (21.30, 44.00)** |  |  |
| **Fasting Insulin** |  |  |  |
| Total association | 0.85 (0.76, 0.93) | <0.001 |  |
| Direct association | 0.79 (0.70, 0.87) | <0.001 | -0.2 |
| Indirect association | 0.06 (0.04, 0.08) | <0.001 | 0.1 |
| Proportion mediated, % | **7.03 (4.87, 10.00)** |  |  |
| **HOMA-IR** |  |  |  |
| Total association | 0.26 (0.24, 0.29) | <0.001 |  |
| Direct association | 0.24 (0.21, 0.26) | <0.001 | -0.2 |
| Indirect association | 0.03 (0.02, 0.04) | <0.001 | 0.1 |
| Proportion mediated, % | **10.39 (7.65, 14.00)** |  |  |
| **Total Cholesterol** |  |  |  |
| Total association | 1.50 (1.09, 1.91) | <0.001 |  |
| Direct association | 1.50 (1.10, 1.94) | <0.001 |  |
| Indirect association | -0.00 (-0.11, 0.10) | 0.89 |  |
| Proportion mediated, % | - | - |  |
| **Triglycerides** |  |  |  |
| Total association | 5.66 (4.82, 6.57) | <0.001 |  |
| Direct association | 4.61 (3.67, 5.57) | <0.001 | -0.1 |
| Indirect association | 1.06 (0.75, 1.36) | <0.001 | 0.1 |
| Proportion mediated, % | **18.70 (12.90, 25.00)** |  |  |
| **LDL Cholesterol** |  |  |  |
| Total association | 1.04 (0.71, 1.36) | <0.001 |  |
| Direct association | 1.09 (0.75, 1.43) | <0.001 |  |
| Indirect association | -0.06 (-0.13, 0.02) | 0.17 |  |
| Proportion mediated, % | - | - |  |
| **HDL Cholesterol** |  |  |  |
| Total association | -0.42 (-0.52, -0.32) | <0.001 |  |
| Direct association | -0.33 (-0.44, -0.23) | <0.001 | 0.1 |
| Indirect association | -0.08 (-0.11, -0.05) | <0.001 | <0.1 |
| Proportion mediated, % | **19.72 (12.37, 29.00)** |  |  |
| **Non-HDL Cholesterol** |  |  |  |
| Total association | 1.92 (1.54, 2.29) | <0.001 |  |
| Direct association | 1.84 (1.46, 2.23) | <0.001 |  |
| Indirect association | 0.08 (-0.02, 0.17) | 0.11 |  |
| Proportion mediated, % | - | - |  |
| **Remnant Cholesterol** |  |  |  |
| Total association | 0.88 (0.71, 1.05) | <0.001 |  |
| Direct association | 0.75 (0.56, 0.93) | <0.001 | -0.1 |
| Indirect association | 0.13 (0.08, 0.19) | <0.001 | 0.1 |
| Proportion mediated, % | **15.15 (9.37, 23.00)** |  |  |
| *Note. Adjusted for age, sex, education, income, smoking status, alcohol status.* | | | |
| *^a^ - Rho is the effect size of unknown confounder at which direct or indirect association becomes 0* | | | |
| *Note. SBP Systolic Blood Pressure, DBP Diastolic Blood Pressure, FBG Fasting Blood Glucose, HbA1c Glycosylated Haemoglobin, HOMA-IR Homeostatic Model Assessment for Insulin Resistance, LDL Low Density Lipoprotein, HDL High Density Lipoprotein* | | | |

| **Table S26** |  |  |  |
| --- | --- | --- | --- |
| *TS26. Adjusted direct and indirect associations of BMI with Cardiometabolic Markers via Waist Circumference* | | |  |
| **Association** | **β (95% CI)** | **P Value** | **ρ^a^** |
| **SBP** |  |  |  |
| Total association | 1.02 (0.87, 1.18) | <0.001 |  |
| Direct association | 1.00 (0.77, 1.24) | <0.001 |  |
| Indirect association | 0.02 (-0.14, 0.18) | 0.82 |  |
| Proportion mediated, % | - |  |  |
| **DBP** |  |  |  |
| Total association | 0.61 (0.51, 0.70) | <0.001 |  |
| Direct association | 0.43 (0.29, 0.57) | <0.001 | -0.1 |
| Indirect association | 0.18 (0.08, 0.28) | <0.001 | 0.1 |
| Proportion mediated, % | **29.07 (13.93, 48.00)** |  |  |
| **FBG** |  |  |  |
| Total association | 1.08 (0.67, 1.48) | <0.001 |  |
| Direct association | -0.63 (-1.29, -0.01) | 0.05 |  |
| Indirect association | 1.71 (1.23, 2.22) | <0.001 |  |
| Proportion mediated, % | - |  |  |
| **HbA1C** |  |  |  |
| Total association | 0.06 (0.05, 0.08) | <0.001 |  |
| Direct association | -0.01 (-0.03, 0.02) | 0.52 |  |
| Indirect association | 0.07 (0.05, 0.09) | <0.001 |  |
| Proportion mediated, % | - |  |  |
| **Fasting Insulin** |  |  |  |
| Total association | 0.85 (0.77, 0.93) | <0.001 |  |
| Direct association | 0.60 (0.49, 0.70) | <0.001 | -0.3 |
| Indirect association | 0.25 (0.18, 0.32) | <0.001 | 0.1 |
| Proportion mediated, % | **29.40 (20.50, 38.00)** |  |  |
| **HOMA-IR** |  |  |  |
| Total association | 0.26 (0.24, 0.29) | <0.001 |  |
| Direct association | 0.15 (0.11, 0.19) | <0.001 | -0.2 |
| Indirect association | 0.12 (0.09, 0.14) | <0.001 | 0.1 |
| Proportion mediated, % | **43.55 (32.41, 55.00)** |  |  |
| **Total Cholesterol** |  |  |  |
| Total association | 1.50 (1.09, 1.90) | <0.001 |  |
| Direct association | 1.84 (1.15, 2.51) | 0.034 |  |
| Indirect association | -0.34 (-0.77, 0.09) | 0.12 |  |
| Proportion mediated, % | - |  |  |
| **Triglycerides** |  |  |  |
| Total association | 5.66 (4.70, 6.59) | <0.001 |  |
| Direct association | 1.89 (0.48, 3.14) | 0.01 | -0.1 |
| Indirect association | 3.77 (2.79, 4.86) | <0.001 | 0.1 |
| Proportion mediated, % | **66.60 (48.60, 91.00)** |  |  |
| **LDL Cholesterol** |  |  |  |
| Total association | 1.04 (0.71, 1.37) | <0.001 |  |
| Direct association | 1.55 (1.02, 2.03) | 0.006 | -0.1 |
| Indirect association | -0.51 (-0.86, -0.17) | 0.002 | <0.1 |
| Proportion mediated, % | **-49.10 (93.00, 16.00)** |  |  |
| **HDL Cholesterol** |  |  |  |
| Total association | -0.42 (-0.52, -0.31) | <0.001 |  |
| Direct association | -0.01 (-0.14, 0.14) | 0.96 |  |
| Indirect association | -0.41 (-0.52, -0.30) | <0.001 |  |
| Proportion mediated, % | **-** |  |  |
| **Non-HDL Cholesterol** |  |  |  |
| Total association | 1.92 (1.52, 2.28) | <0.001 |  |
| Direct association | 1.84 (1.27, 2.35) | <0.001 |  |
| Indirect association | 0.07 (-0.30, 0.50) | 0.74 |  |
| Proportion mediated, % | - |  |  |
| **Remnant Cholesterol** |  |  |  |
| Total association | 0.88 (0.71, 1.07) | <0.001 |  |
| Direct association | 0.30 (0.05, 0.57) | 0.018 | -0.1 |
| Indirect association | 0.58 (0.39, 0.77) | <0.001 | 0.1 |
| Proportion mediated, % | **66.32 (43.26, 94.00)** |  |  |
| *Note. Adjusted for age, sex, education, income, smoking status, alcohol status.* | | | |
| *^a^ - Rho is the effect size of unknown confounder at which direct or indirect association becomes 0* | | | |
| *Note. SBP Systolic Blood Pressure, DBP Diastolic Blood Pressure, FBG Fasting Blood Glucose, HbA1c Glycosylated Haemoglobin, HOMA-IR Homeostatic Model Assessment for Insulin Resistance, LDL Low Density Lipoprotein, HDL High Density Lipoprotein* | | | |

| **Table S27** |  |  |  |
| --- | --- | --- | --- |
| *TS27. Adjusted direct and indirect associations of BMI with Cardiometabolic Markers via Visceral Fat* | | | |
| **Association** | **β (95% CI)** | **P Value** | **ρ^a^** |
| **SBP** |  |  |  |
| Total association | 1.02 (0.87, 1.18) | <0.001 |  |
| Direct association | 0.94 (0.74, 1.14) | <0.001 |  |
| Indirect association | 0.08 (-0.04, 0.22) | 0.18 |  |
| Proportion mediated, % | - |  |  |
| **DBP** |  |  |  |
| Total association | 0.61 (0.52, 0.71) | <0.001 |  |
| Direct association | 0.51 (0.37, 0.63) | <0.001 | -0.2 |
| Indirect association | 0.10 (0.02, 0.19) | 0.006 | <0.1 |
| Proportion mediated, % | **17.11 (3.58, 32.00)** |  |  |
| **FBG** |  |  |  |
| Total association | 1.08 (0.65, 1.48) | <0.001 |  |
| Direct association | 0.35 (-0.24, 0.89) | 0.22 |  |
| Indirect association | 0.73 (0.36, 1.12) | <0.001 |  |
| Proportion mediated, % | - |  |  |
| **HbA1C** |  |  |  |
| Total association | 0.06 (0.05, 0.08) | <0.001 |  |
| Direct association | 0.04 (0.02, 0.06) | <0.001 | -0.1 |
| Indirect association | 0.02 (0.01, 0.04) | <0.001 | 0.1 |
| Proportion mediated, % | **39.03 (17.54, 66.00)** |  |  |
| **Fasting Insulin** |  |  |  |
| Total association | 0.85 (0.77, 0.94) | <0.001 |  |
| Direct association | 0.79 (0.69, 0.89) | <0.001 | -0.4 |
| Indirect association | 0.06 (0.004, 0.12) | 0.038 | <0.1 |
| Proportion mediated, % | **7.09 (0.43, 14.00)** |  |  |
| **HOMA-IR** |  |  |  |
| Total association | 0.26 (0.24, 0.29) | <0.001 |  |
| Direct association | 0.23 (0.20, 0.27) | <0.001 | -0.3 |
| Indirect association | 0.03 (0.01, 0.06) | 0.004 | 0.1 |
| Proportion mediated, % | **12.78 (3.86, 21.00)** |  |  |
| **Total Cholesterol** |  |  |  |
| Total association | 1.50 (1.09, 1.95) | <0.001 |  |
| Direct association | 2.90 (2.41, 3.41) | <0.001 | -0.3 |
| Indirect association | -1.40 (-1.73, -1.07) | <0.001 | -0.1 |
| Proportion mediated, % | **-93.50 (-139.80, -66.00)** |  |  |
| **Triglycerides** |  |  |  |
| Total association | 5.66 (4.77, 6.63) | <0.001 |  |
| Direct association | 3.87 (2.76, 5.06) | <0.001 | -0.2 |
| Indirect association | 1.80 (1.01, 2.62) | <0.001 | 0.1 |
| Proportion mediated, % | **31.70 (17.80, 48.00)** |  |  |
| **LDL Cholesterol** |  |  |  |
| Total association | 1.04 (0.70, 1.37) | <0.001 |  |
| Direct association | 2.19 (1.75, 2.63) | <0.001 | -0.3 |
| Indirect association | -1.15 (-1.43, -0.90) | <0.001 | -0.1 |
| Proportion mediated, % | **-125.27 (98.23, 170.00)** |  |  |
| **HDL Cholesterol** |  |  |  |
| Total association | -0.42 (-0.52, -0.31) | <0.001 |  |
| Direct association | 0.11 (-0.01, 0.23) | 0.084 |  |
| Indirect association | -0.52 (-0.61, -0.44) | <0.001 |  |
| Proportion mediated, % | - |  |  |
| **Non-HDL Cholesterol** |  |  |  |
| Total association | 1.92 (1.53, 2.32) | <0.001 |  |
| Direct association | 2.80 (2.31, 3.28) | <0.001 | -0.3 |
| Indirect association | -0.88 (-1.18, -0.58) | <0.001 | -0.1 |
| Proportion mediated, % | **-46.00 (-65.70, -29.00)** |  |  |
| **Remnant Cholesterol** |  |  |  |
| Total association | 0.88 (0.71, 1.04) | <0.001 |  |
| Direct association | 0.61 (0.38, 0.82) | <0.001 | -0.1 |
| Indirect association | 0.27 (0.12, 0.41) | <0.001 | 0.1 |
| Proportion mediated, % | **31.10 (13.60, 50.00)** |  |  |
| *Note. Adjusted for age, sex, education, income, smoking status, alcohol status.* | | | |
| *^a^ - Rho is the effect size of unknown confounder at which direct or indirect association becomes 0* | | | |
| *Note. SBP Systolic Blood Pressure, DBP Diastolic Blood Pressure, FBG Fasting Blood Glucose, HbA1c Glycosylated Haemoglobin, HOMA-IR Homeostatic Model Assessment for Insulin Resistance, LDL Low Density Lipoprotein, HDL High Density Lipoprotein* | | | |

**Supplementary Section 6**

### *SS6. Sensitivity and Specificity of central obesity indices*

| **Table S28** |  |  |  |  |  |  |
| --- | --- | --- | --- | --- | --- | --- |
| *TS28. Sensitivity and specificity of central obesity indices for cardiometabolic disease stratified by sex and BMI status* | | | | | | |
| **Central Obesity Marker** | **Hypertension** | | **Diabetes** | | **Dyslipidemia** | |
|  | **Sensitivity^a^** | **Specificity^b^** | **Sensitivity^a^** | **Specificity^b^** | **Sensitivity^a^** | **Specificity^b^** |
| **Female** | | | | | | |
| **Normal BMI** |  |  |  |  |  |  |
| **WC (IDF)** | 34.1% (27.2%-41.5%) | 75.2% (71.4%-78.6%) | 36.1% (27.1%-45.9%) | 74.5% (71.0%-77.8%) | 27.5% (24.3%-30.9%) | 83.3% (67.2%-93.6%) |
| **WC (WHO)** | 34.1% (27.2%-41.5%) | 75.2% (71.4%-78.6%) | 36.1% (27.1%-45.9%) | 74.5% (71.0%-77.8%) | 27.5% (24.3%-30.9%) | 83.3% (67.2%-93.6%) |
| **WC (ATP)** | 9.5% (5.6%-14.8%) | 94.0% (91.7%-95.8%) | 13.9% (8.0%-21.9%) | 94.3% (92.3%-96.0%) | 6.9% (5.2%-9.0%) | 94.4% (81.3%-99.3%) |
| **WHR** | 80.4% (73.9%-86.0%) | 22.9% (19.6%-26.6%) | 91.7% (84.8%-96.1%) | 24.4% (21.2%-27.9%) | 78.3% (75.1%-81.2%) | 30.6% (16.3%-48.1%) |
| **WHtR** | 54.7% (47.2%-62.2%) | 55.3% (51.2%-59.4%) | 47.2% (37.5%-57.1%) | 53.0% (49.1%-56.9%) | 47.4% (43.7%-51.2%) | 61.1% (43.5%-76.9%) |
| **VO** | 4.5% (1.9%-8.6%) | 97.1% (95.3%-98.3%) | 5.6% (2.1%-11.7%) | 97.1% (95.5%-98.2%) | 3.3% (2.1%-4.9%) | 97.2% (85.5%-99.9%) |
| **Abnormal BMI** |  |  |  |  |  |  |
| **WC (IDF)** | 83.8% (79.5%-87.4%) | 27.2% (23.8%-31.0%) | 85.3% (80.9%-89.0%) | 27.2% (23.9%-30.8%) | 77.7% (74.9%-80.3%) | 51.7% (32.5%-70.6%) |
| **WC (WHO)** | 83.8% (79.5%-87.4%) | 27.2% (23.8%-31.0%) | 85.3% (80.9%-89.0%) | 27.2% (23.9%-30.8%) | 77.7% (74.9%-80.3%) | 51.7% (32.5%-70.6%) |
| **WC (ATP)** | 52.9% (47.6%-58.2%) | 63.8% (59.8%-67.6%) | 56.5% (50.9%-62.1%) | 64.4% (60.6%-68.0%) | 42.8% (39.6%-46.1%) | 72.4% (52.8%-87.3%) |
| **WHR** | 93.3% (90.2%-95.6%) | 8.2% (6.1%-10.6%) | 94.9% (91.8%-97.1%) | 8.8% (6.8%-11.3%) | 92.7% (90.8%-94.3%) | 17.2% (5.8%-35.8%) |
| **WHtR** | 92.2% (88.9%-94.7%) | 11.9% (9.5%-14.7%) | 93.9% (90.7%-96.3%) | 12.5% (10.1%-15.3%) | 90.2% (88.1%-92.0%) | 31.0% (15.3%-50.8%) |
| **VO** | 55.5% (50.1%-60.7%) | 64.6% (60.7%-68.4%) | 57.2% (51.5%-62.7%) | 64.1% (60.3%-67.8%) | 43.3% (40.1%-46.5%) | 72.4% (52.8%-87.3%) |
| **Male** | | | | | | |
| **Normal BMI** |  |  |  |  |  |  |
| **WC (IDF)** | 7.6% (4.5%-11.8%) | 95.9% (93.7%-97.5%) | 10.2% (6.1%-15.9%) | 96.3% (94.4%-97.7%) | 5.0% (3.5%-7.1%) | 93.2% (84.9%-97.8%) |
| **WC (WHO)** | 3.6% (1.5%-6.9%) | 99.0% (97.6%-99.7%) | 5.4% (2.5%-10.0%) | 99.3% (98.1%-99.8%) | 2.1% (1.1%-3.5%) | 100.0% (95.1%-100.0%) |
| **WC (ATP)** | c | c | c | c | c | c |
| **WHR** | 76.4% (70.3%-81.8%) | 29.0% (25.0%-33.3%) | 88.6% (82.7%-93.0%) | 32.1% (28.2%-36.2%) | 74.1% (70.5%-77.5%) | 39.2% (28.0%-51.2%) |
| **WHtR** | 48.9% (42.2%-55.6%) | 63.8% (59.3%-68.1%) | 58.4% (50.5%-66.0%) | 65.3% (61.1%-69.3%) | 41.2% (37.3%-45.1%) | 67.6% (55.7%-78.0%) |
| **VO** | 76.4% (70.3%-81.8%) | 34.0% (29.7%-38.4%) | 80.7% (73.9%-86.4%) | 34.1% (30.1%-38.3%) | 70.7% (66.9%-74.2%) | 41.9% (30.5%-53.9%) |
| **Abnormal BMI** |  |  |  |  |  |  |
| **WC (IDF)** | 69.2% (64.5%-73.6%) | 44.5% (40.3%-48.8%) | 72.2% (67.6%-76.6%) | 46.4% (42.2%-50.7%) | 61.7% (58.5%-64.9%) | 46.5% (31.2%-62.3%) |
| **WC (WHO)** | 45.1% (40.3%-50.1%) | 69.3% (65.3%-73.2%) | 45.0% (40.1%-50.0%) | 68.9% (64.9%-72.7%) | 37.3% (34.2%-40.5%) | 72.1% (56.3%-84.7%) |
| **WC (ATP)** | 11.2% (8.3%-14.6%) | 93.8% (91.4%-95.7%) | 12.2% (9.2%-15.9%) | 94.5% (92.2%-96.2%) | 8.6% (6.9%-10.6%) | 97.7% (87.7%-99.9%) |
| **WHR** | 95.6% (93.2%-97.4%) | 5.7% (3.9%-7.9%) | 97.8% (95.8%-99.0%) | 7.1% (5.2%-9.6%) | 94.9% (93.2%-96.2%) | 4.7% (0.6%-15.8%) |
| **WHtR** | 96.6% (94.4%-98.1%) | 6.2% (4.3%-8.6%) | 97.2% (95.1%-98.6%) | 6.6% (4.7%-9.0%) | 95.0% (93.4%-96.3%) | 4.7% (0.6%-15.8%) |
| **VO** | 97.6% (95.6%-98.8%) | 2.6% (1.4%-4.2%) | 97.2% (95.1%-98.6%) | 2.3% (1.2%-3.9%) | 97.6% (96.4%-98.5%) | 4.7% (0.6%-15.8%) |
| *Note. BMI Body Mass Index, WC Waist Circumference, WHR Waist-Hip Ratio, WHtR Waist-Height Ratio, VO Visceral Obesity, IDF International Diabetes Federation, WHO World Health Organisation, NCEP ATP III National Cholesterol Education Plan Adult Treatment Panel III* | | | | | | |
| *^a^ - sensitivity (95% Confidence Intervals)* | |  |  |  |  |  |
| *^b^ - specificity (95% Confidence Intervals)* | |  |  |  |  |  |
| *c - No males with normal BMI and central obesity according to NCEP ATPIII criteria.* | | | |  |  |  |
