## Supplementary material for "Dual Metrics of Obesity: Evaluating BMI and Central Obesity as Indicators of Cardiometabolic Risk in Rural India": Graphical Abstract

### "Asian Indian Phenotype"

Greater visceral fat

Lesser skeletal muscle mass

Greater risk for cardiometabolic disease (CMD)

*At the same BMI as caucasians.*

### Dual Metric Obesity Criteria

**Normal Weight**
**No Central Obesity**

Healthy

**Normal Weight**
**Central Obesity**

Males at increased risk for CMD

**Abnormal Weight**
**No Central Obesity**

Females at increased risk for CMD

**Abnormal Weight**
**Central Obesity**

**Greatest CMD risk**

**Waist-Hip Ratio**
**Waist-Height Ratio**
Visceral Fat Percentage
Waist Circumference

**Cardiometabolic Risk Profile**

Stronger association of CMD and Central Obesity seen in :

Middle-Age (45-58 years) > Older-Age (58+ years)

Males > Females

**Conclusion:** Central obesity is an important risk factor for cardiometabolic diseases independently and in combination with abnormal BMI. These association are sex and age-specific. Waist-Hip Ratio and Waist-Height Ratio are accurate and practical measures of central obesity in rural Indian settings.
